# PathML: A unified framework for whole-slide image analysis with deep learning

**DOI:** 10.1101/2021.07.07.21260138

**Authors:** Adam G. Berman, William R. Orchard, Marcel Gehrung, Florian Markowetz

## Abstract

The inspection of stained tissue slides by pathologists is essential for the early detection, diagnosis and monitoring of disease. Recently, deep learning methods for the analysis of whole-slide images (WSIs) have shown excellent performance on these tasks, and have the potential to substantially reduce the workload of pathologists. However, successful implementation of deep learning for WSI analysis is complex and requires careful consideration of model hyperparameters, slide and image artefacts, and data augmentation. Here we introduce PathML, a Python library for performing preand post-processing of WSIs, which has been designed to interact with the most widely used deep learning libraries, PyTorch and TensorFlow, thus allowing seamless integration into deep learning workflows. We present the current best practices in deep learning for WSI analysis, and give a step-by-step guide using the PathML framework: from annotating and pre-processing of slides, to implementing neural network architectures, to training and post-processing. PathML provides a unified framework in which deep learning methods for WSI analysis can be developed and applied, thus increasing the accessibility of an important new application of deep learning.

## 1 Introduction

In histopathology, tissue biopsies are fixed, embedded, sectioned, stained, and placed on a glass slide before being examined under a microscope. Examination of tissue slides to identify pathologically relevant features has been an essential tool for early detection, diagnosis and disease monitoring in medical practice and research for decades. Pathological features can be anything from the presence or absence of certain cell types or populations, changes in cellular or nuclear morphology, changes in the arrangement of cells in a tissue, to changes in the intensity of certain tissue stains. Until recently only expert pathologists have been able to perform this task, requiring years of training, and with individual slides often having to be evaluated by multiple pathologists before a judgement can be made (*1*). However, with a shift towards digitisation in pathology, tissue-slides are now routinely scanned to produce high-resolution whole-slide images (WSIs). Such images are amenable to automated image analysis and in the last decade the field has undergone a revolution. Deep learning methods for image analysis have shown excellent performance on diagnostic tasks (*1–3*), rivalling that of pathologists and further stimulating efforts to digitise glass slides. Recent work has even shown the capability of deep learning approaches to find correlations between tissue appearance and genomic features (*4*).

Pathologists have high inter-observer concordance rates on some diagnostic tasks, but in others they frequently disagree (*5*). This is compounded by high workload, necessitating rapid screening of individual cases, increasing the risk of introducing diagnostic errors (*6*). Deep learning methods are fast, often requiring only a few minutes to evaluate a slide, and give consistent evaluations. Thus, deep learning has the potential to substantially reduce the workload of pathologists, improve the inter-observer concordance rates and accelerate the evaluation of tissue-slides.

Despite this potential, deep learning based approaches have not yet seen widespread uptake in medical practice. This is in part due to a lack of a unified framework in which WSI neural network implementations are developed and applied, meaning that individual researchers often must re-implement their own preand post-processing pipelines in-house for each new histopathology task. Furthermore, successful implementation of deep learning to WSI analysis requires careful consideration of model hyperparameters, slide and image artefacts and data augmentation beyond those encountered in standard image analysis, and thus application of the latest advances in computer vision to WSI analysis is hampered without a framework for streamlined WSI processing into which such advances can be incorporated.

Here we introduce PathML, a new Python library for performing preand post-processing of WSI data in a unified framework. PathML simplifies and streamlines many of the steps required to tackle the unique challenges posed by WSIs. In particular, WSIs are very large, typically multiple gigabytes in size (*7*), and thus it is necessary to break up WSIs into ‘tiles’ before they can be analysed by contemporary deep learning architectures (see **Tiling**). Tiling, however, introduces further difficulties as WSIs are often labelled (e.g. cancerous or non-cancerous) at the slide level, not at the tile level, and so a deep learning approach must be adopted which accounts for how tiles inherit labels from slides (see **Annotation**). Furthermore, WSIs can contain unique artefacts introduced during slide preparation and imaging, which are not found in other image analysis settings, such as pen marks left by the pathologists reviewing them, or cracks and bubbles in the slide. All of these artefacts must be removed or accounted for when training a deep learning model (see **Deep tissue detector**). In addition, the staining and imaging protocols can differ widely between institutions, which if not accounted for can have a profound impact on a deep learning model’s ability to generalise across institutions (*8*). If an approach is to be successfully implemented in medical practice, it is essential that it is able to perform reliably across institutions. As such, data augmentation approaches aimed at accounting for WSI-specific issues have been developed, however, these introduce further important considerations (see **Data augmentation**). Finally, WSI datasets frequently encompass only a small number of slides, and occurrences of pathological features may be rare. To avoid overfitting in this setting, steps must be taken during data augmentation and training (see **Data augmentation**). PathML has been designed to interact with current popular deep learning libraries, PyTorch and TensorFlow (*9, 10*), allowing it to be seamlessly incorporated into deep learning workflows. By tackling the unique challenges posed by WSIs, PathML can help to translate deep learning methods into the clinic more easily, providing a broad method to replace *ad hoc* solutions. We made extensive use of PathML in our recent publication (*11*) applying deep learning to early detection of Barrett’s oesophagus in WSIs. PathML is available from https://github.com/markowetzlab/pathml.

In this article we explain key concepts and best practices in deep learning for image analysis, and give a step-by-step guide for performing WSI analysis with PathML: from annotating and pre-processing slides, to implementing neural network architectures, to training and post-processing (**fig. 1**). We do not discuss more general neural network theory, such as optimisers and choosing an appropriate learning rate, as these are very well covered elsewhere (*12–14*). We envisage this article to be useful both for users experienced with Python, but having only basic experience with deep learning and WSIs, as well as users who have extensive experience with deep learning and standard image analysis, but not with WSI analysis. The order of the following sections roughly follows the order in which each concept is encountered in a typical data analysis project.

**Figure 1:**
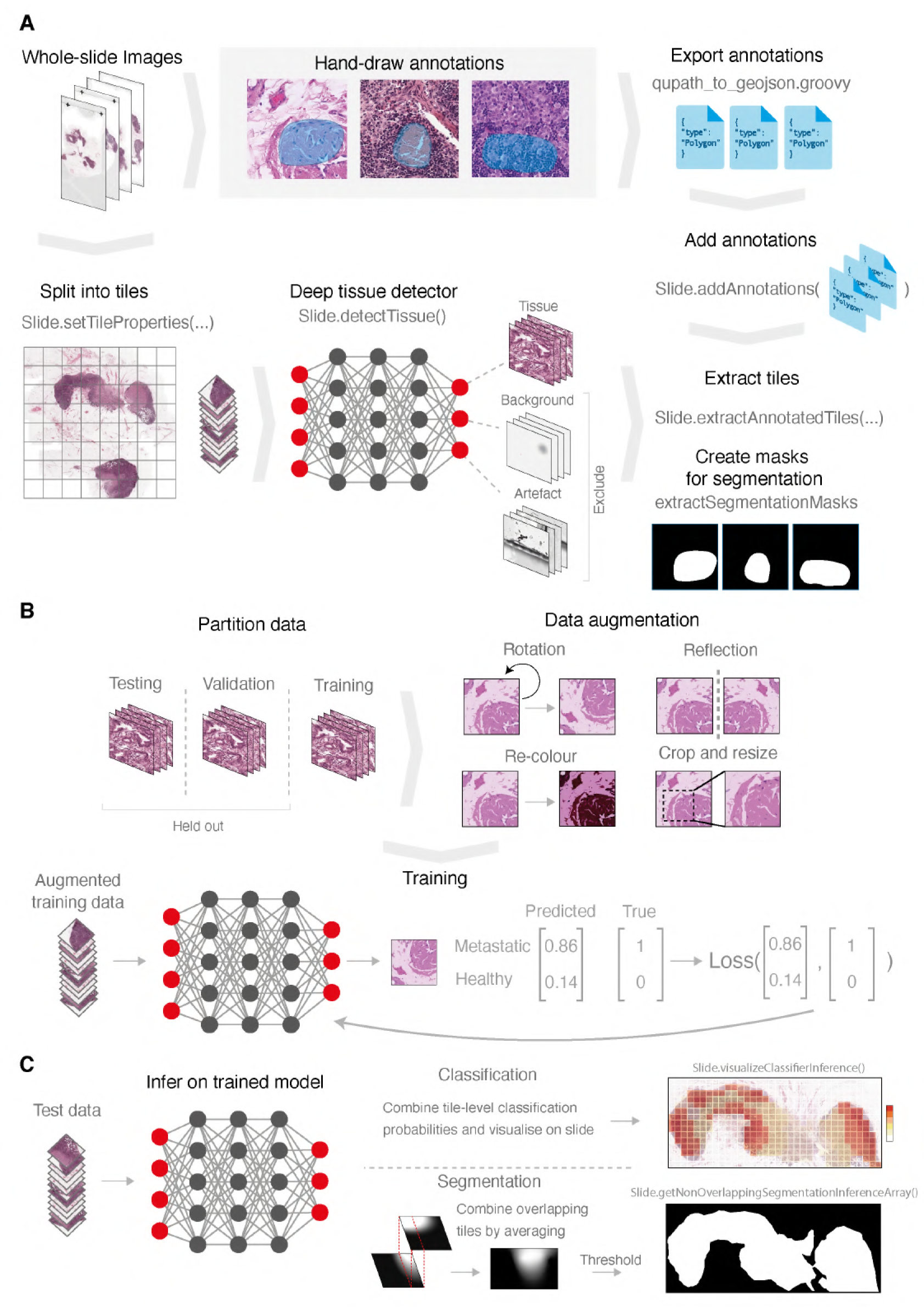
A deep learning pipeline with PathML. **(A)** Creating PathML Slide objects, exporting and adding annotations, and extracting tiles and segmentation masks. **(B)** Data partitioning, data augmentation, and model training. **(C)** Inferring on the trained model and stitching together overlapping segmentation results (if required).

### 1.1 Deep learning

In medical imaging, there are three sorts of tasks for which deep learning models are widely used: classification, segmentation or regression. Classification tasks involve training a model to be able to predict which label, out of a predefined set, should be assigned to a given image. For example, within a WSI, one may want to predict whether the tissue within a given tile should be labelled as ‘cancerous’ or ‘healthy’. Training is performed in a ‘supervised’ manner, meaning that the model is given examples for which a ground-truth label is already known and trained to correctly predict that label. During training, the parameters of the model, called ‘weights’, are adjusted to minimise the difference between the prediction the model makes and the groundtruth, with the discrepancy being computed using a ‘loss function’. The choice of loss function is dependent on the task being performed, and for classification tasks, the loss typically takes the form of a cross-entropy (see **table 1** for a short glossary of terms for deep learning on WSIs). In medical imaging, in the supervised setting, segmentation is a closely related task to classification. Segmentation tasks involve training a model to be able to divide an image up into a patchwork of regions, or ‘segments’, at the pixel-level, such that the pixels within a segment all belong to the same class. For example, a segmentation model might indicate which pixels of a WSI are tissue and which are background. In this case the model is trained with ground-truth ‘segmentation masks’: pixel-level bounded regions delineating segments from one another, and the loss function computes the degree of overlap between the predicted segmentation mask, and the ground-truth segmentation mask. A common choice is the ‘Dice loss’ (*15*). Finally, regression is similar to classification, however, instead of an image belonging to one of a finite set of classes, an image is instead assigned a continuous value. For example, one may train a model to predict the number of weeks survival from a WSI of a tumour tissue biopsy taken from a patient (*16*). Likewise, the ground-truth in this case is also a continuous value, and a different loss function used; typically the *L^2^*-distance between the predicted value and the ground-truth.

**Table 1:** A short glossary of important terms for deep learning on whole-slide images.

| **Term** | **Description** |
| --- | --- |
| Annotation | A digital line drawn around a region of interest in a WSI |
| Architecture | The specific structure of a neural network |
| CNN | A convolutional neural network, a type of neural network amenable to learning from image data |
| Data augmentation | A series of methods to reduce model overfitting by randomly altering tiles before each epoch |
| Epoch | One full pass of a model through all training examples, often split up into mini-batches |
| H&E | Hematoxylin and Eosin, a general histological stain to highlight cell nuclei blue and cytoplasm pink |
| Hyperparameter | Adjustable parameter that influences how training happens |
| Kernel | A pattern that is scanned across the image input into a convolutional neural network and at each point compared (by taking the dot product) to the underlying image. Also called a ‘filter’. |
| Label | A category or class into which a piece of data falls; what the neural network is trying to learn to predict. Also called the ‘ground truth’. |
| Loss function | The algorithm used to compute the difference between a model’s predicted output and the ground truth |
| Mini-batch | A subset of the dataset from which the model learns during training |
| Overfitting | A model learns a representation that corresponds too closely to the training dataset, such that the model generalises poorly to new examples |
| Optimisation function | The algorithm used to modify the weights in a model according to the loss accumulated during a mini-batch. |
| Tile | A small square subregion of a WSI with side length generally between 100 and 1000 pixels. Also called a ‘patch’. |
| Training | A series of passes a model makes through training examples to learn a generalisable correlation between examples and their labels |
| Transfer learning | Using knowledge acquired on one machine learning task to a different one. Often means: starting with an already trained model (‘pretrained’) to solve an unrelated problem. |
| WSI | Whole-slide image, a very high resolution image of a tissue slide taken with a specialised machine |

Which of these tasks is the goal of the deep learning model influences many of the decisions that the user needs to make. Each task requires different ground-truth annotations (see **Annotation**), and different neural network architectures are best suited for each task (see **Neural network architectures**), and how training and model evaluation is performed will differ too (see **Training, inference and validation**). Despite this, the general workflow in PathML will remain unchanged, and where there are differences, they will be explicitly highlighted and explained.

### 1.2 Whole-slide Image formats

When glass slides are digitised by digital whole-slide image scanners, high-resolution images are taken at multiple magnifications. WSIs therefore have a pyramidal data structure, with the images taken at each magnification each forming a ‘layer’ of the WSI. The maximum magnification of these images is frequently 200X (using a 20X objective lens at 10X magnification) or 400X (using a 40X objective lens at 10X magnification). Several different file formats have been developed to store pyramidal WSIs, some of which are proprietary and specific to the scanning machine, while others, such as the NDPI and pyramidal TIFF, are open source. Several open source software projects have been developed for reading and performing operations on pyramidal whole slide image files, including OpenSlide and Bio-Formats, which have libraries for many programming languages including C, C++, Ruby, and Python (*17, 18*).

PathML uses the pyvips library for reading WSIs (*19*), and so supports a wide range of formats, including NDPI and pyramidal TIFF. WSIs are instantiated as `Slide` objects by calling the `Slide` class on the file path to the WSI and specifying which layer you would like to access with the `level` argument. It is through `Slide` objects that the user interacts with their data and performs the pre-processing and post-processing steps described below and in detail in the **Procedure**.

### 1.3 Tiling

WSI images are very large; for example, an image scanned at 40X objective power of a 20 x 20 mm sample of tissue has 80,000 x 80,000 pixels; at standard 24-bit colour this would produce a flat image 19.2GB in size. Current neural network architectures are unable to process images of this size in one go. Thus, WSIs are broken up into ‘tiles’ or ‘patches’ upon which the model is trained: small square regions of the original image, typically 32 to 1000 pixels in height. Tiles can be chosen with or without overlap with neighbouring tiles. Choice of tile dimensions and overlap are some of the most important hyperparameters to choose when analysing WSIs (*7*). Tiles should be large enough that features relevant to the task are visible to the model to learn. For example, if nuclear morphology is a crucial feature for distinguishing healthy cells from cancerous ones, tiles should not be smaller than the width of a nucleus in the image. In the case of cytological images, a tile covers a single cell (5-10 µm). In histopathology, broader architectural features of the tissue need to be considered and so tiles need to be larger (10- 250 µm). A rule of thumb is that a pathologist should be able to readily classify or segment the ground-truth class by examining only each tile in isolation. Deep learning architectures have pre-defined input sizes to which a tile will need to be resized, and thus a tile should also not be too large as this may cause the relevant features to appear overly coarse after resizing to fit the input shape of the deep learning model. Furthermore, if tiles are being classified, then larger tiles will also decrease the resolution at which classification labels are assigned to the WSI, and the presence of multiple classes within a single tile may mean features are ‘diluted’ and missed. Choosing tiles which do not overlap has the advantage of reducing the numbers of tiles extracted from an image, decreasing both training and inference times. The risk, however, is that without overlap, relevant features will be split between adjacent tiles. During training, this will mean that the feature is not available in full for the model to learn from. During inference, a feature which would be identified by the model if visible in full may not be identifiable in any of the tiles individually when split. The size of tile overlap is therefore frequently chosen to encompass roughly half the length of a typical feature, for instance, half the length of an average cell of the tissue type being analysed.

In PathML, tile dimensions and overlap are chosen by calling the `Slide.setTileProperties()` method, and setting the `tileSize` and `tileOverlap` arguments, enabling users to easily experiment with different tiling strategies. Here one can also specify how tiles should be stored, and how they will be accessed during training. By calling `Slide.extractAnnotationTiles()`, one can extract and store each tile in individual images files in advance of training or inferring. PathML will automatically store tile image files according to their class (see **Annotation** below) and slide of origin in a directory structure appropriate for use with PyTorch (see **Procedure step 7** and **fig. 3**). Although functional, storing each individual tile image file may pose data storage issues. PathML stores the coordinates of each tile rather than the tile image itself, accessible with `Slide.getTile()` using the tile address as argument (all tile addresses can be iterated over with `Slide.iterateTiles()`), making it easy to build a dataset such that tiles are accessed on-the-fly, saving the need to extract each tile to an image file in situations where this would be too memory intensive. Apart from being substantially less disk memory intensive, this approach also makes it easier to experiment with different tiling strategies without having to re-extract tiles for each combination of tile size and overlap. The trade off is that on-the-fly tile accession approaches are typically much slower to train with, so are not recommended except in datasets where tiles number in the hundreds of thousands or millions and disk memory for these tile images is not available.

### 1.4 Deep tissue detector

WSIs can contain unique artefacts introduced during slide preparation and imaging which are not found in other image analysis settings. Tissue may tear and fold during slide preparation, the image may be unevenly illuminated or stained, and parts of the image may be out of focus. Tissue slides also often contain pen marks left by the pathologists reviewing them, and may contain cracks and bubbles. Left unaccounted for, such artefacts can have severe detrimental effects on a deep learning model. For instance, pen marks are often left by pathologists to indicate the presence of a pathological feature of interest, such as the presence of cancerous cells. If not removed, a deep learning model may simply learn to recognise the presence of a pen mark in slides containing cancer, and thereby be completely inapplicable in medical practice where no such annotation will be available. Beyond artefacts, WSIs will typically contain large portions of background, i.e. regions without any tissue, which do not contain any pathologically relevant information. After tiling your WSI, tiles which contain artefacts or which simply display background should therefore be removed speed up the training process and potentially improve performance.

PathML provides a built-in deep tissue detector: a DenseNet neural network architecture trained to classify tiles as either ‘artefact’, ‘background’, or ‘tissue’. PathML’s deep tissue detector was trained using 9,071 tiles extracted from 393 individual annotations from 61 WSIs scanned across a variety of machines, time periods, and tissue types, and two different species to account for a broad range of the variation of WSI artefacts, background, and tissue appearances (see **fig. 2A**). The deep tissue detector is applied by calling the `Slide.detectTissue()` method (see **Procedure step 5** for details), enabling robust detection of tissue tiles. The outputs are the probabilities, for each tile, of belonging to each of the three classes. In addition to the deep tissue detector included in PathML, a number of other approaches have been developed to detect and remove artefacts and background tiles which we briefly review in **Related methods** below.

**Figure 2:**
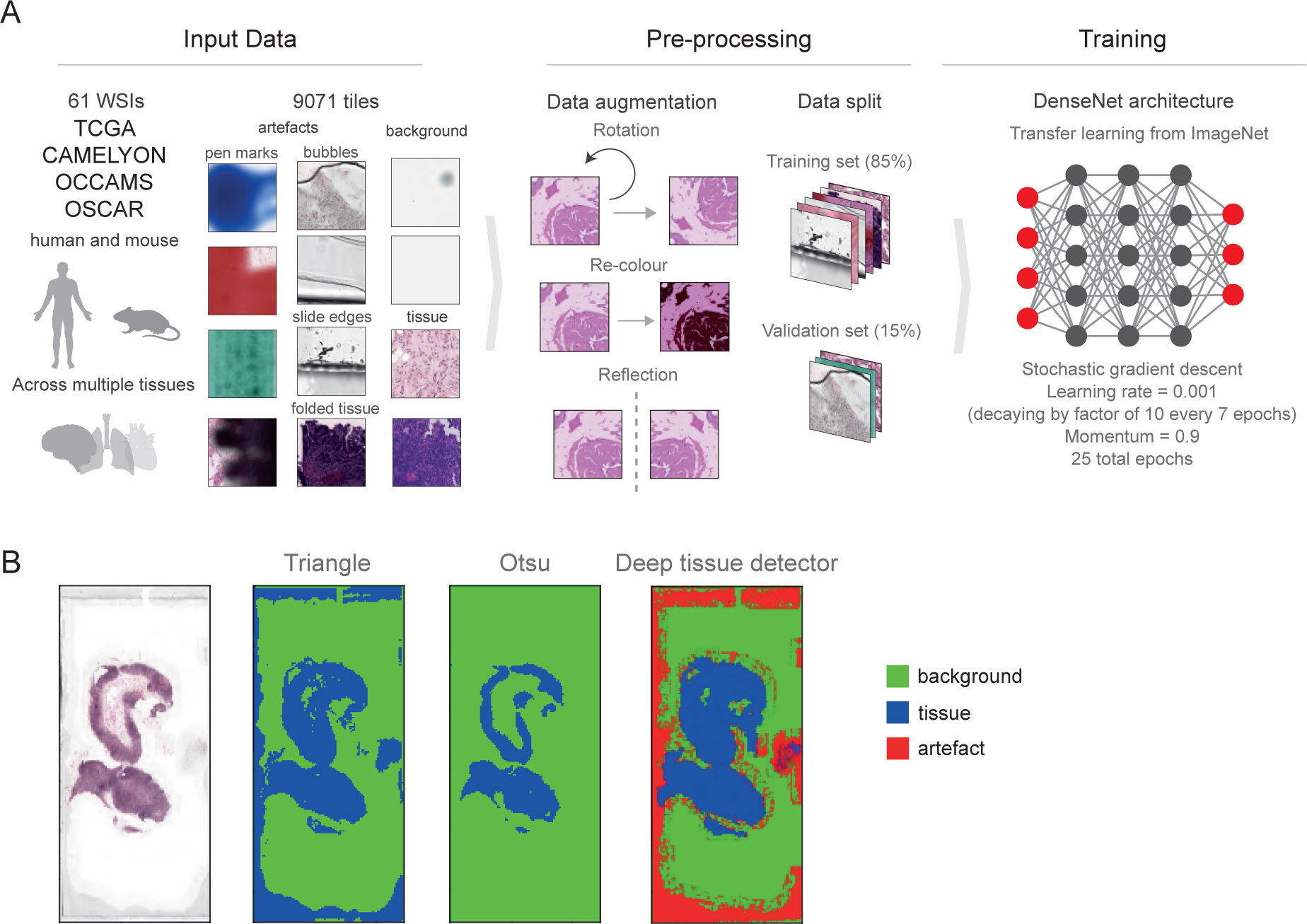
Tissue, artefact, and background detection with PathML. **(A)** Tiling, augmenting, and training a DenseNet CNN to classify tissue, artefact, and background regions on WSIs from a robust dataset representing multiple tissue and species types. This already-trained deep tissue detector can be applied to any PathML Slide object with PathML’s Slide.detectTissue() function. **(B)** The results for the task of background filtering on a CAMELYON16 WSI (tumor 021.tif) using the triangle algorithm, Otsu’s method, and PathML’s deep tissue detector. The triangle algorithm incorrectly calls slide artefacts tissue, Otsu’s method excludes lighter-coloured tissue regions (calling them background), and PathML’s deep tissue detector makes neither mistake while identifying artefact regions separately from background. The triangle algorithm and Otsu’s method can be applied with PathML’s Slide.detectForeground() function. Visualisations of filtering performance like these can be created with PathML’s Slide.visualizeForeground() and Slide.visualizeTissueDetection() functions.

**Figure 3:**
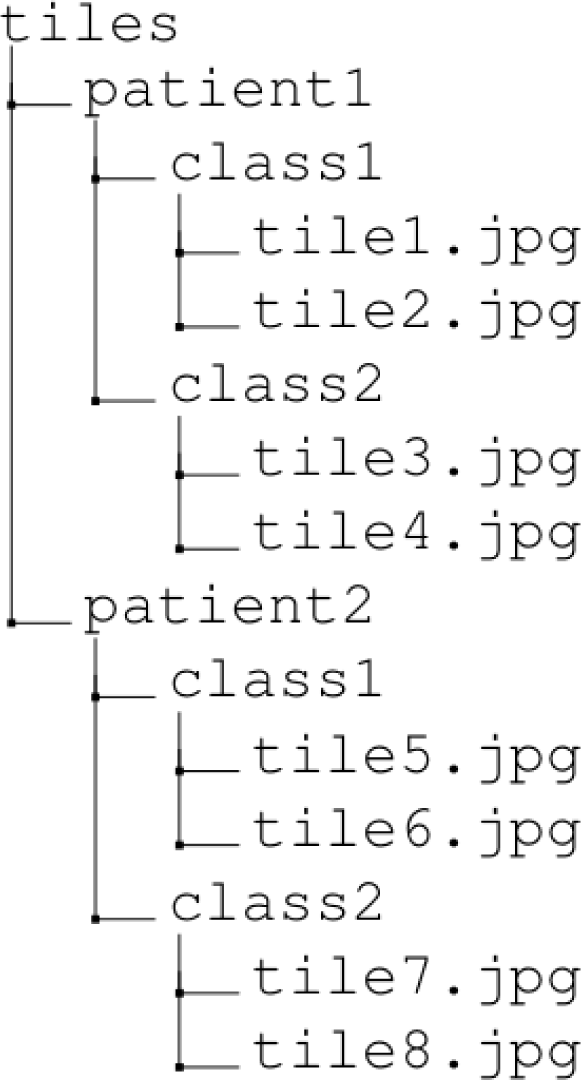
The directory structure output by Slide.extractAnnotationTiles() and Slide.extractRandomUnannotatedTiles().

There may be applications where the artefacts encountered are not well covered by the deep tissue detector in PathML, and thus one should always review examples of its output to verify that the detector is behaving as expected (see **Procedure step 6** and **fig. 4**). Where the user wishes to make use of the curated catalogue of WSIs and corresponding annotations used for training the Deep tissue detector, see **Data availability**. Furthermore, PathML makes it easy to apply a user-provided tissue detection model trained on additional, or alternative, annotated images by setting the `modelStateDictPath` and `architecture` parameters when calling `Slide.detectTissue()`, to the path to the custom model and its neural network architecture, respectively. In certain cases, users may want to make use of classical foreground filtering approaches in place of or in addition to the deep tissue detector. In PathML, this can be achieved by calling the `Slide.detectForeground()` method; specifying the desired approach with the `threshold` argument. PathML currently supports Otsu’s method (*20*), the triangle algorithm (*21*), as well as simple intensity thresholding (see **Procedure step 5**). In general, however, these approaches are far less robust to the diversity of artefacts observed in WSIs, as well as appearances of background and tissue, often requiring careful supervision for each application (*7*). **Figure 2B** compares the performance of Otsu’s method, the triangle algorithm, and PathML’s deep tissue detector on an example slide: whereas Otsu’s method is too strict (excluding lighter-coloured tissue regions as background) and the triangle algorithm is too permissive (including artefacts around the edge of the slide as tissue), the deep tissue detector makes neither of these mistakes, correctly identifying all true tissue while demarcating artefact and background appropriately.

**Figure 4:**
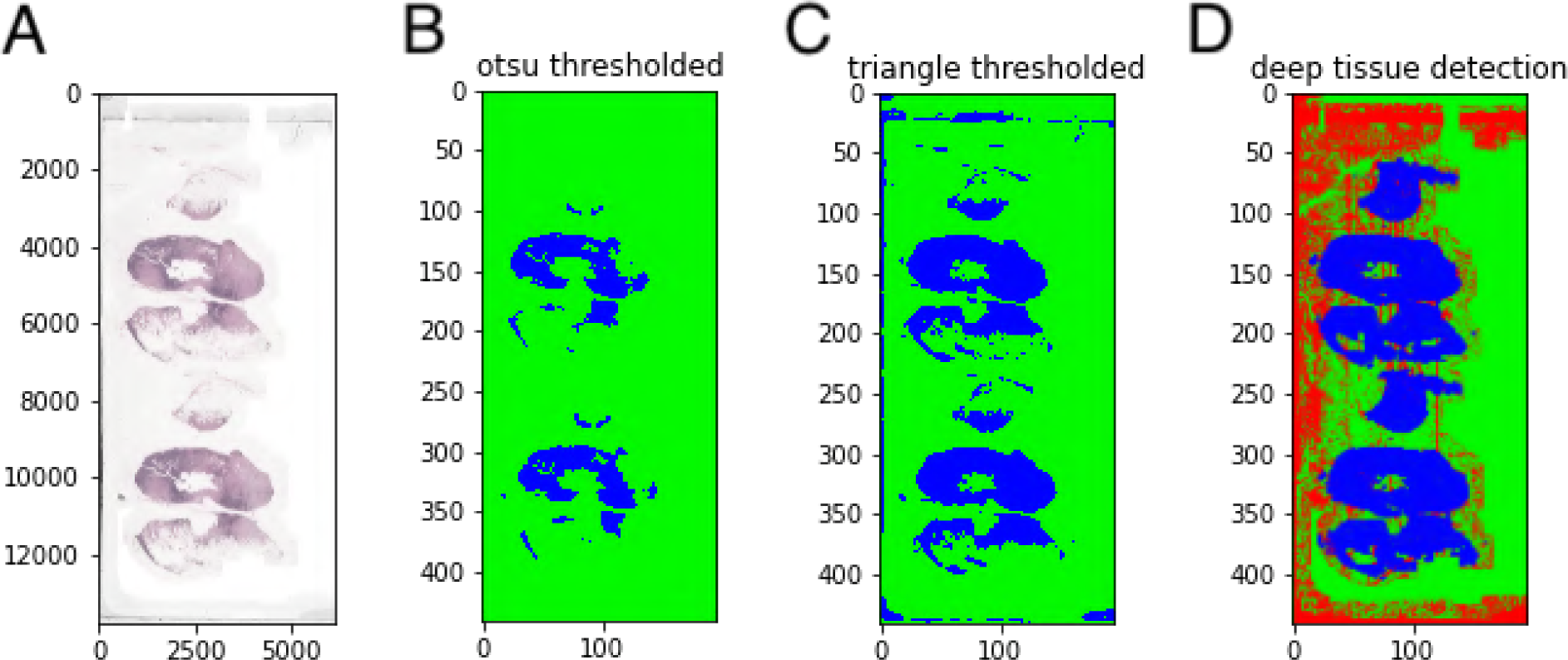
Foreground and deep tissue detection results. Blue is foreground/tissue, green is background, and red is artefact. **(A)** A low-resolution image of the WSI tumor 046.tif generated with Slide.visualizeThumbnail(). **(B)** Foreground filtering with Otsu’s method generated by PathML’s Slide.visualizeForegroundDetection(’otsu’). **(C)** Foreground filtering with the triangle method generated by PathML’s Slide.visualizeForegroundDetection(’triangle’). **(D)** Tissue detection with PathML’s built-in deep tissue detector generated by PathML’s Slide.visualizeTissueDetection().

### 1.5 Annotation

Ground-truth labels for a WSI may exist either at the region-level, wherein they are local to particular regions within the WSI, or at the slide-level, wherein a label applies to the WSI as a whole. Region-level labels typically take the form of digital annotations on the WSI, delineating the regions belonging to certain classes. Specialised software, such as QuPath and Automated Slide Analysis Platform (ASAP), allow users to draw digital annotations onto WSIs and then export them for use in image analysis workflows (*22, 23*). PathML supports the use of annotation files in the GeoJSON format as well as the XML format output by ASAP (*24*). QuPath annotations can be exported as GeoJSON files using the Groovy script, `qupath_to_geojson.groovy` provided in https://github.com/markowetzla `b/pathml-tutorial`.

Whether classifying or segmenting or segmentation labels, it is important to consider how to derive the ground truth labels of individual tiles from slide-wide label data. For segmentation tasks the principle is straightforward, because the ground-truth is at the pixel-level and thus whole-slide segmentation masks can also be tiled and directly inherited by the individual image tiles. PathML allows the user to easily create tile-level binary segmentation masks given an annotation file. An annotation file is added to a corresponding `Slide` object by calling the `Slide.addAnnotations()` method, providing the file path to the `annotationFilePath` argument. The annotations do not need to cover the entire WSI, instead PathML creates a binary segmentation mask by designating all pixels bounded by an annotation for a given class as being positive for that class, and all other pixels as negative. PathML provides a flexible framework for annotations: When applying the `Slide.extractAnnotationTiles()` method, binary segmentation masks are created by the `extractSegmentationMasks` argument set to `True`, and the `classesToExtract` argument set to the name of the class (or list of classes) for which the binary segmentation mask (or masks) should be created. By default, segmentation mask tiles for all classes found in the annotation file will be extracted if `extractSegmentationMasks` is utilised. The tile-level masks are then saved to a ‘masks’ directory in the location specified by the `outputDir` argument of `Slide.extractAnnotationTiles()`. Furthermore, the `numTilesToExtractPerClass` argument can be set to the maximum number of tiles to extract per class from the slide (default is 100 for each class) when there are more extractable tiles per class than the user desires. In addition, after calling the `Slide.detectTissue()` method on a given `Slide` object, each tile will be inferred on the deep tissue detector, and the resulting tissue probability will be saved at each tile. This will then allow the `tissueLevelThreshold` argument can to be used in subsequent functions such as the tile extraction functions (`Slide.extractAnnotationTiles()` and `Slide.extractRandomUnannotatedTiles()`) to set a minimum threshold for this probability for a tile to be used (recommended value of 0.995). Likewise, `foregroundLevelThreshold` can be used to only extract tiles with the desired simple foreground detection techniques such as (*20*) (set the argument to ‘otsu’) or the triangle algorithm (*21*) (set the argument to ‘triangle’). Simple average greyscale intensity filtering can be achieved by setting `foregroundLevelThreshold` to an integer between 0 and 100 (see Procedure step 7).

For classification tasks the situation is more complicated. For region-level labels, PathML applies a simple heuristic: a tile inherits the label of any annotations that cover more than a given threshold fraction of the area of the tile. Tiles which are not covered above this threshold for any annotations are ignored entirely. To achieve this in PathML, one begins as for segmentation, but when using the `Slide.extractAnnotationTiles()` method, the `tileAnnotationOverlapThreshold` argument (`default = 0.5`) should be set to the fraction of the area of a tile that must be covered by an annotation for it to inherit the corresponding label (see **Procedure step 7**). The appropriate threshold is determined by how precisely the annotation demarcates the region of interest, and how large a fraction of the area of a tile it is expected to occupy. If annotations are imprecise, encompassing not only the region of interest, but also surrounding tissue, then the threshold should be large enough that tiles which overlap an annotation, but which do not display an instance of the class, are not labelled as such. In contrast, if an entire region of interest only occupies a small fraction of the area of a tile, then the threshold should be small enough that the tile containing it is appropriately labelled. However, if the tile size has been properly chosen, this is unlikely to be a concern.

In histopathology, region-level ground-truth labels are frequently not available, and instead labels exist only at the slide-level. In the ‘strongly supervised’ setting, it is assumed that all the tiles in a slide (excluding background and artefact tiles) inherit the slide-level label. This may be appropriate in cases where all of the tiles in a slide, or at least a large majority, display representative features of the slide-level label. For example, if nearly all the tissue in a tumour biopsy slide is disease tissue, it may be appropriate that all tiles inherit a slide-level patient survival label, as the majority of the tiles likely display features predictive of prognosis. However, often the strongly supervised setting is not appropriate; for instance, if a tissue biopsy consists mostly of healthy tissue with only small regions of metastasis, a slide-level label of ‘metastasis’ should not be inherited by all tiles. In these cases a ‘weakly supervised’ method is preferable. The weakly supervised setting borrows from multiple instance learning, where it is instead assumed that a slide-level label implies the presence of at least *k* instances, i.e. tiles, within the WSI which correspond to the slide-level label. Particular neural network architectures have been designed to exploit this assumption in order to perform weakly supervised learning (see **Neural network architectures**) (*25–27*). Although weakly supervised learning can be a very powerful paradigm, and often the only option, it is less robust, performs less well, and requires more data than when performing strongly supervised learning with region-level labels (*28, 29*).

### 1.6 Data augmentation

Data augmentation describes a collection of techniques used to create additional training data by performing small transformations on existing data. Data augmentation serves to regularise deep learning models and reduce overfitting (see **table 1**) by artificially expanding the amount and diversity of the data they are trained on. For image data, common transformations include adding random noise, introducing random rotations and reflections, randomly adjusting bright-ness, contrast, saturation and hue, and randomly resizing and cropping the images.

Data augmentation is particularly important for WSI analysis due to two primary factors: Firstly, staining and imaging protocols can differ widely between institutions, producing WSIs which vary substantially in terms of their hue, contrast and stain intensity. As such, deep learning models which have been trained only on WSIs produced at a single institution may generalise poorly to WSIs produced elsewhere. Secondly, WSI datasets tend to be small, often consisting of only a few hundred slides. The combination of having a wide diversity in the appearance of WSIs, and the fact that datasets typically only sample a small portion of that diversity, means that deep learning models are at particular risk of overfitting.

Many ‘standard’ image analysis data augmentation techniques are recommended for WSI analysis in most settings. Random rotations and reflections help to train deep learning models to ignore the orientation of pathological features, e.g. a metastasis should still be identified as such independent of its orientation. Random cropping involves cropping out a small area of an image, and resizing the remainder to fit the original size. This helps the model become robust to features being cut off (for instance, by the edge of the tile), and to changes in their scale caused by small variations in the magnification the slide was scanned at. Augmentations consisting of random adjustments of the brightness, contrast, saturation and hue are collectively called ‘colour augmentations’, and are crucial in WSI analysis for making the model robust to changes in imaging and staining protocols between institutions. All of these data augmentation techniques can be easily applied to WSI data through PyTorch (see **Procedure steps 10 and 25**). Colour augmentation approaches specific to WSI analysis have also been developed, such as those proposed by Tellez et al (*8*) and Bug et al (*30*) for hematoxylin and eosin (H&E) stained slides, which have been tailored to mimic true H&E stain variations. It is also common to apply ‘Gaussian blur’ augmentations to mimic small out-of-focus regions, an image artefact often encountered in WSIs. In contrast to excluding such artefacts from analysis altogether (see **Deep tissue detector**), data augmentation instead allows the user to train the model to become robust to their presence, potentially making more efficient use of the data.

It is important, however, that data augmentation is applied with care. In general, augmented data should not deviate significantly from a ‘realistic’ range, otherwise one may inadvertently train their model to ignore informative features, causing underfitting and poorer performance (*31*). For example, by applying heavy colour augmentations to WSIs, one can produce images which do not resemble images produced with any imaging or staining protocol. In this case, the deep learning model is likely to learn to discard colour and intensity information when making a prediction, as it appears uninformative. In settings where it is desired that colour information is discarded, for instance when a pathological feature is largely (or entirely) characterised by morphological characteristics, deliberate use of extreme colour augmentations can be beneficial. In settings where stain intensity information is highly informative, such as for Imaging Mass Cytometry (IMC) data, even mild colour jittering could therefore be highly detrimental. Likewise, morphological features are often important for distinguishing healthy tissue from diseased tissue, and so applying data augmentations which warp morphological features in the image, such as an elastic transform, would likely be detrimental for most histopathological tasks. It is therefore recommended that the user always visualises samples of augmented data to assess whether it appears to be within the expected range (see **Procedure steps 12 and 26**, **fig. 5**, and **fig. 6**).

**Figure 5:**
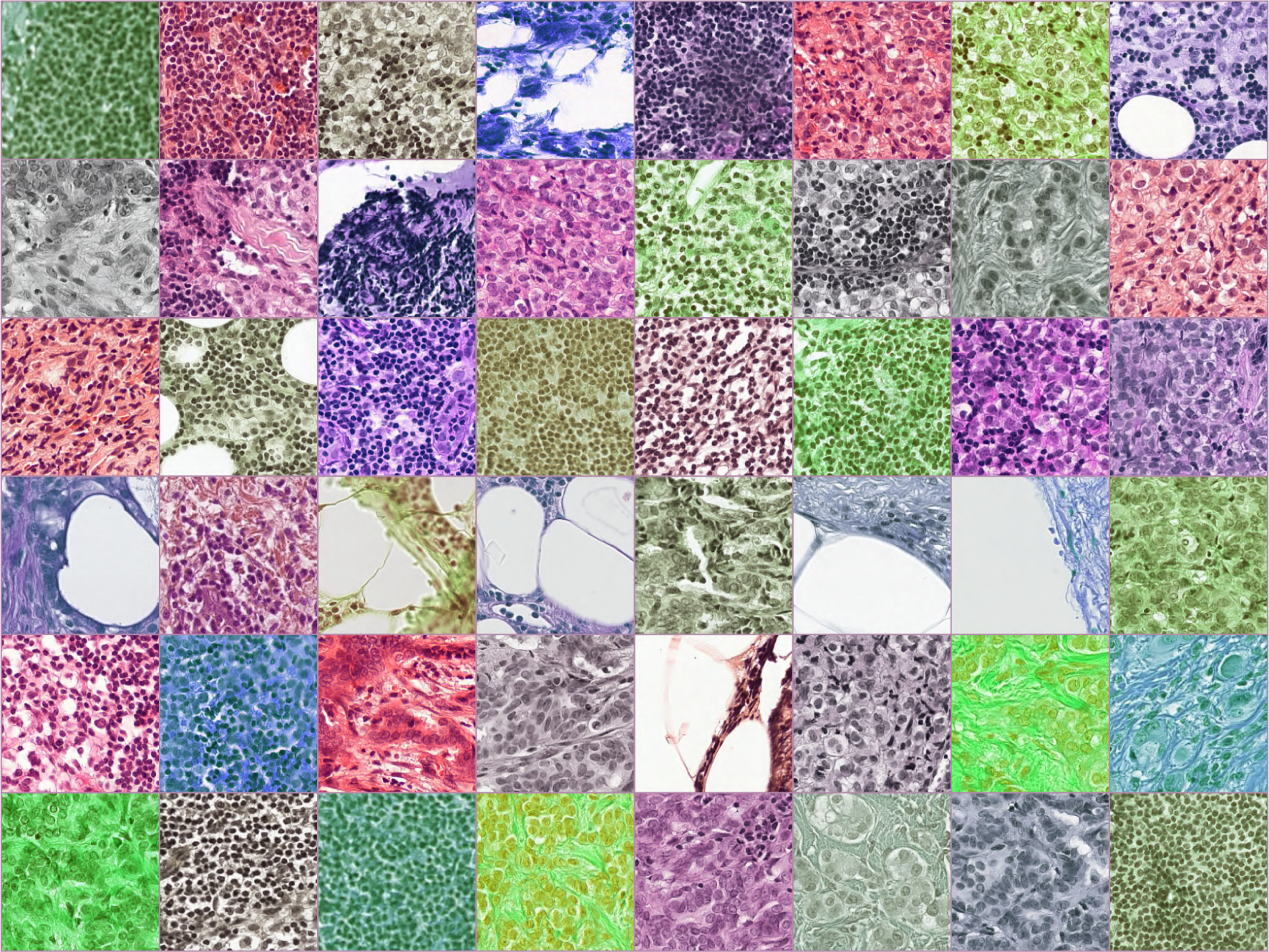
One batch of tiles with random data augmentations. Augmentations include random horizontal flip, random vertical flip, and random colour jitter including saturation and hue jittering (with jitter values of 1 and 0.5 respectively). Augmentations are performed with PyTorch’s torchvision library (*9*).

**Figure 6:**
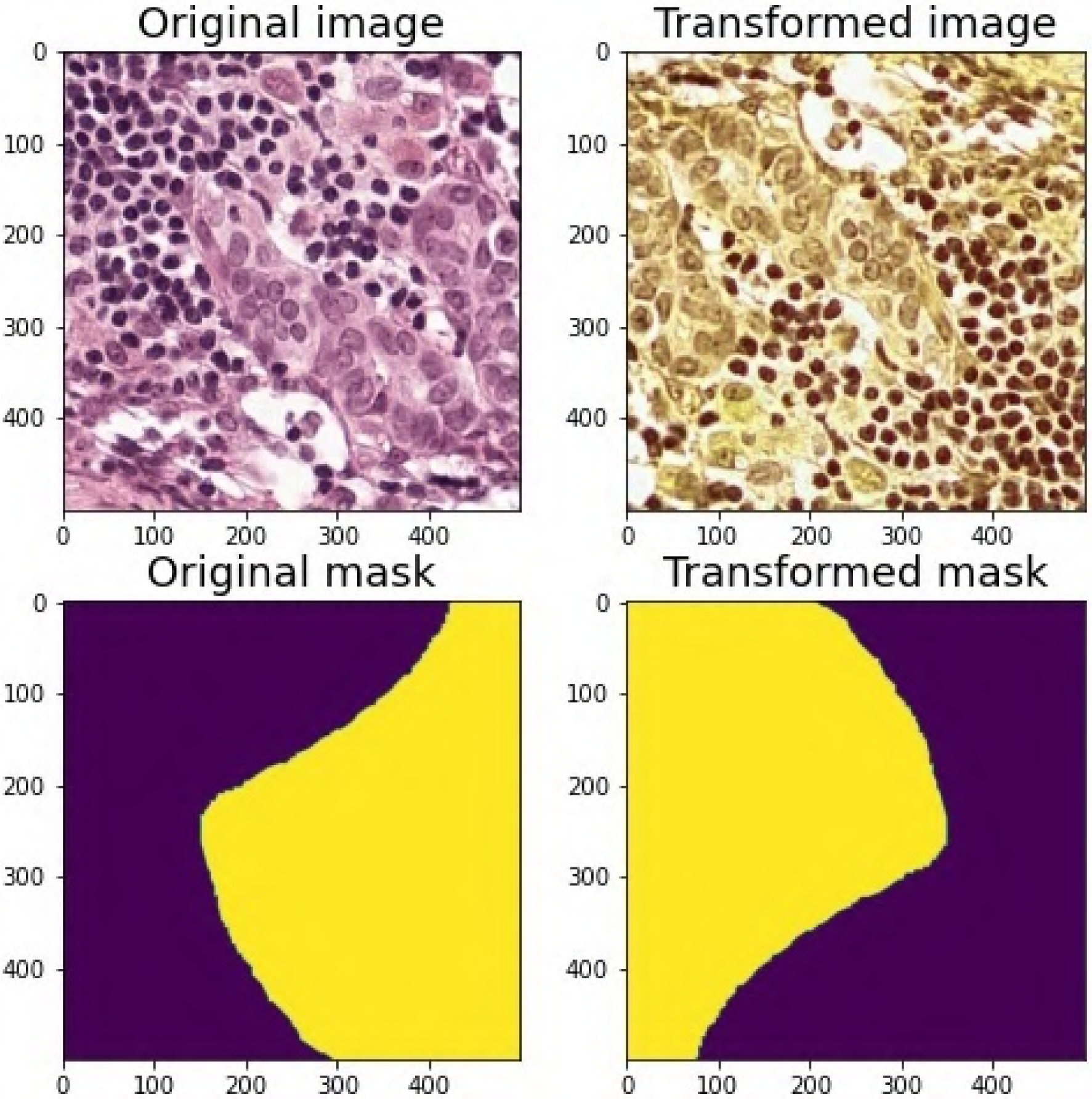
Visualising the effects of augmentation on a tile and matching segmentation mask. As desired, the spatial transformations (in this example horizontal and vertical flip) are preserved between the image and the mask, whereas the colour transformations (random saturation and hue jitter with values 1 and 0.5 respectively) are performed only on the image and not on the mask. Tandem image and mask augmentations like these can be performed with PyTorch’s Albumentations library (*58*).

Another data augmentation technique important in WSI analysis is ‘synthetic oversampling’. It is common in WSI analysis to have large class imbalances. For example, in early cancer detection screening, a pathologist may be looking for a single instance of an atypical cell within a tissue slide. In this case, most tiles extracted from a WSI will display only healthy cells, and thus a model trained on this data will be exposed to many more examples of healthy tissue than atypical tissue. As a result, the model will tend to overfit to the healthy tissue, and have a poor accuracy when classifying atypical tissue. To address this class imbalance, synthetic oversampling involves re-sampling from the data for a minority class such that the training data becomes balanced. The issue with synthetic oversampling is that because tiles from the minority class are simply randomly duplicated, the model does not learn more robust features, and may become overfit to the minority class instead. Consequently, this technique does not perform well with highly imbalanced data (*32*).

With its functions `Slide.extractAnnotationTiles()` and `Slide.extractRandomUnannotatedTiles()`, PathML generates tile images into a directory structure that can be used as direct input to PyTorch’s `torchvision.datasets.ImageFolder` dataset creator (*33*), enabling easy access to the data augmentation solutions provided by PyTorch. Furthermore, by allowing users to request exactly how many tiles they would like extracted from each class using the `numTilesToExtractPerClass` of the two tile extraction functions above, PathML makes it straightforward to avoid massive class imbalance issues.

### 1.7 Neural network architectures

Convolutional neural networks (CNNs) have revolutionised the field of computer vision, and are the class of neural networks at the centre of deep learning approaches in WSI analysis. CNNs have become the state-of-the-art for image analysis because they exploit three properties which are found in imaging data: locality, stationarity and compositionality (*34*). Locality describes the fact that in images, the pixels close to each other give more information about each other than pixels which are far apart. Stationarity describes the fact that the same ‘patterns’, for instance shapes and textures, often reoccur within and across images. Compositionality describes the fact that objects in images are composed of combinations of increasingly complex patterns. CNNs are able to exploit these properties by making use of ‘convolutional layers’. In a convolutional layer, a kernel, or filter, which describes a pattern, is scanned across the image and at each point compared (by taking the dot product) to the underlying image. The more the pattern and the underlying image match, the larger the score at that point. The size of the kernel is a tunable hyperparameter in a CNN, but exploits locality by being relatively small (e.g. 3x3 or 5x5), collecting information only from nearby pixels. Scanning the same kernel across the image exploits the stationarity property, as a reoccurring pattern will be detected wherever it is in the image. Finally, compositionality is exploited by having the neural network contain multiple convolution layers, the inputs of each being (transformed) outputs from the layers before. In this way, deeper convolution layers capture complex compositions of patterns detected in previous layers. In classical image analysis, similar kernel convolution operations are used, but in this case each kernel has been handcrafted by researchers. In CNNs, each kernel is learned during training, enabling highly complex and abstract information to be captured.

There is a large variety of CNN architectures to choose from which differ in the sizes of kernels they apply, and the number and types of layers they contain. Different architectures are more suitable for certain tasks than others, and so choosing the most appropriate for a particular application is a key consideration. For tile-level classification tasks, popular CNN architectures include ResNet (*35*), Inception (*36*), VGG (*37*), and DenseNet (*38*). Though these architectures have all shown excellent performance, their main drawback is their size: they contain large numbers of layers and millions of parameters. Without a sufficiently large dataset, such large architectures are susceptible to overfitting to the training data. One option in this case is to use a smaller architecture, and many have been designed to give competitive performance with fewer layers and parameters. For example, SqueezeNet (*39*) has proven to be effective for training on WSI tiles across a variety of studies. For regression tasks, the final layers of these architectures are typically adapted to output a continuous value and the loss function changed to one appropriate for regression. Although any of these architectures can also be adapted to perform segmentation tasks, the U-Net architecture (*40*) was specifically developed for segmentation of medical images, and is one of the most popular architectures for performing segmentation of WSIs.

For classification in the weakly supervised setting, certain specialised architectures have been developed, such as CHOWDER (*41*). However, simple approaches to weakly supervised learning can still make use of the aforementioned architectures when combined with a modified training procedure. For example, one approach makes use of two assumptions: first, that for a WSI with a negative slide-level label, all tiles inherit the negative label, and second, for a WSI with a positive slide-level label, only the top *k* tiles, as predicted by the model, inherit the positive label. This means: for a WSI with a slide-level label of ‘healthy’, it is assumed that all tiles extracted from the image display healthy tissue; while for a WSI with a slide-level label of ‘cancerous’, only the top *k* tiles predicted by the model to be cancerous inherit the label. These assumptions define an approach for slide-level label inheritance during training, enabling standard classification architectures to be applied, but do not generally make efficient use of the data. In WSIs where there are more than *k* tiles displaying a positive feature, these additional tiles are simply discarded. In contrast, specialised architectures like CHOWDER aim to make more efficient use of the data.

Once a model has been trained, PathML supports the use of any conventional CNN architecture in PyTorch for inference on `Slide` objects with its `Slide.inferClassifier()` and `Slide.inferSegmenter()` functions for classification and segmentation models, respectively. The trained model file is used as an argument, and PathML then infers each tile that passes the user’s requested tissue and foreground filtering standards through the model. The inference results of each tile are saved directly into the `Slide` object, enabling use of downstream analysis functions to evaluate inference performance, such as `Slide.classifierMetricAtThreshold()`, `Slide.segmenterMetricAtThreshold()` (to check an inference statistic at a threshold or thresholds), `Slide.visualizeClassifier()`, and `Slide.visualizeSegmenterInference()` (to generate a visual inference map at a class of inference). See **Training, inference and validation**, **Procedure steps 17, 19, 30, and 32**, and **Anticipated results** for more details of the these functions, their application, and their results.

### 1.8 Transfer learning

We have already discussed using data augmentation and small CNN architectures as strategies to reduce the risk of overfitting. Another very important approach, particularly widespread in computer vision, is transfer learning. Typically when training a neural network, one will first initialise its weights by sampling from a standard normal distribution. In transfer learning, instead of beginning training with random weight values, the weights are instead given their values by learning on a separate dataset first, typically from a different domain. The rationale for transfer learning in CNNs is that, whether the model was trained to distinguish cats from dogs, or healthy cells from cancerous ones, many of the kernels learnt during training will be similar, particularly in early convolutional layers. For example, it is likely that kernels for detecting edges would be learnt in both settings. By initialising the weights in this way, we can therefore transfer this knowledge from one domain to another, without having to relearn it. Most of the CNN architectures discussed in the previous section have been previously trained on the ImageNet dataset (*42*) to be able to detect the presence of thousands of different objects, including over a hundred different breeds of dog. Although ImageNet images are quite different from WSIs, many publications have found transfer learning improves training times and validation accuracy for tasks in other domains (*43*). Transfer learning also reduces the risk of overfitting as it, in effect, increases the size of the training dataset, and so is particularly useful when the training dataset is small. PyTorch and TensorFlow both provide methods to initialise weights for the most popular CNN architectures with those learnt on ImageNet (see **Procedure step 13**) (*9, 10*).

### 1.9 Training, inference and validation

In the previous sections, we have discussed the major pre-processing steps and most important considerations one has to make when performing deep learning on WSIs. In this subsection we give a brief introduction to some further considerations that need to be made when training a deep learning model, and how PathML can help, before discussing applying the model and assessing its performance.

#### 1.9.1 Data partitioning

As in any machine learning setting, one’s data should be split into three disjoint subsets: a training set, validation set and test set. The training set is used to train the deep learning model. The validation set is used to assess the performance of the trained model on an independent dataset, and enable the selection of optimum hyperparameter values. Once the model has been trained, and the hyperparameters chosen which give the best performance on the validation set, the final assessment of the model is performed on the test set. Applying the model to the test set gives an estimate for how it will likely perform when encountering new, unseen data, and thus how it is expected to perform when applied in the clinic. Once the model has been assessed on the test set, no further modifications can be made. A typical split might allocate 70% of the available data for training, 15% percent for validation and 15% for testing; which we will call a ‘70-15-15 split’. For smaller datasets, the validation and test sets are frequently made a larger fraction of the dataset, a 60-20-20 split for example, and for larger datasets more data are apportioned for training, such as a 90-5-5 split, but there are no final, agreed upon ratios (*44*).

For WSIs, although the model is trained at the tile-level, it is important that the partition between training, validation, and test sets is at the patient-level. If there are multiple WSIs for a single patient, all of them should be partitioned into the same set. Likewise, all tiles from the same slide should be part of the same set (*7*). As tiles taken from a single patient are more likely to be similar to each other than tiles from a different patient, having tiles from the same patient appear in both training and test sets could lead to artificially inflated estimates of model performance. It is good practice then to partition patients between the three sets as a first step before training.

#### 1.9.2 Training

Building a dataset with labels that can be interpreted by a deep learning library is an important training consideration. PathML’s tile extraction functions (`Slide.extractAnnotationTiles()` and `Slide.extractRandomUnannotatedTiles()`) output tiles and labels in a directory structures that are by default compliant for direct input into PyTorch’s `torchvision.datasets.ImageFolder` dataset constructor (see **fig. 3**), making it incredibly straightforward to load the tiles PathML has extracted into a format ready for training (see **Procedure step 11**). For classification tasks, per `torchvision.datasets.ImageFolder`, tile labels are stored as directory names within a parent directory containing the slide or case name (*33*). For segmentation tasks, we have included in PathML a custom dataset, `PathmlSegmentationDataset`, that takes as arguments the paths to the `tile` and `mask` directories output by PathML’s tile extraction functions, as well as the desired augmentation transformations, to make a PyTorch-ready segmentation dataset that correctly pairs and parallel-augments tiles and masks (see **Procedure step 27** for an example of its use).

#### 1.9.3 Inference

After a model has been trained, the next step is inferring the final model on the validation and test sets to verify that the model learned appropriately and compute some statistics. Commonly, a metric is measured on the validation set in order to subsequently apply it to the test set (see Validating the model). PathML has two functions that infer a trained model on all tissue-filter passing tiles in a `Slide` object, saving the results into the tile dictionary internal to it: `Slide.inferClassifier()` and `Slide.inferSegmenter()`, which take as input a trained PyTorch model file (**Procedure steps 17 and 30**).

To ensure that the model learned roughly the correct patterns during inference, it is important to visual inference results by plotting the inference predictions of tiles spatially as they appear in the WSI. Once inference has been performed on the `Slide` objects, PathML’s `Slide.visualizeClassifierInference()` and `Slide.visualizeClassifierInference()` functions create these plots for the user overlaid atop a low-resolution image of the WSI below it (**Procedure steps 18 and 31**), taking the class to visual as an argument. If the regions highlighted by the model seem not to correspond to what is expected given the ground truth, trying different training configurations, tile sizes, or other parameters is recommended to solve this issue before proceeding.

For segmentation tasks, it is often important to create a pixel-wise inference mask that is the same pixel dimensions as the WSI (at the pyramid level that was used for tile extraction). If tile overlap was used during inference, it is necessary to merge the overlapping pixel predictions into one prediction to create a single two-dimensional output matrix. PathML’s `Slide.getNonOverlappingSegmentationInferenceArray()` handles all of these tasks, returning an overlap-corrected inference array (see **Procedure step 36** for a usage example).

#### 1.9.4 Validating the model

In addition to visual verification, numeric methods are frequently used to interpret model performance. In particular, performance on the test set is often used as a final check of the model’s efficacy. There are two main scenarios that can occur when using WSIs as data depending on whether test set ground truth is present at the level of tiles or at the level of patients or slides. In the former case, determining test set performance is more straightforward, as tile-level results can be compared to tile-level ground truth, and then averaged across tiles to give patient- or slide-level results. Still, it is often first necessary to determine an operating threshold, both for classification and segmentation tasks. As most conventional CNN architectures output continuous prediction probabilities between 0 and 1, whereas most ground truth labels derived from WSI annotations are discrete class assignments, a method is required to discretise model predictions for comparison to the ground truth. Commonly, a range of prediction probability thresholds are used to discrete predictions, and then the prediction probability that is most performant on the validation set is applied to the test set to measure the model’s success. Once inference has been performed on the validation set, PathML’s `Slide.classifierMetricAtThreshold()` and `segmenterMetricAtThreshold()` functions are purpose-built to do this task; users simply input a list of probability thresholds to try as arguments to these functions, as well as which performance metric to measure (`accuracy`, `balanced_accuracy`, `f1`, `precision`, and `recall` are options for `Slide.classifierMetricAtThreshold()` and `dice_coeff` is available for `segmenterMetricAtThreshold()`) and which class (argument `classToThreshold`) to measure it on, and they return that metric computed at each inputted probability threshold performance metric (see **Procedure steps 19 and 32** for usage examples). The threshold that produced the best performance on the validation set can then be applied to the test set with the same two functions, just inputting the single best threshold in the `probablityThresholds` argument.

In the case where only slide-level labels are present for the test set (very common in medical contexts where manual annotation of WSIs is time-intensive) a method is needed to bridge gap between the tile-level predictions output by the model and the slide-level ground truth of the patient so that they can be compared to measure performance. For example, for classification tasks, a count of the number of tiles positive for the class of interest is compared with slide-level ground truth via AUC analysis to measure test set performance. PathML was made to accommodate such techniques. Its `Slide.numTilesAboveClassPredictionThreshold()` function returns the total number of tiles in the `Slide` whose inference prediction probabilities for a given class (`classToThreshold`) lie at or above the probability thresholds inputted into the `probablityThresholds` argument. These class-positive counts per slide can then be used to generate an AUC compared to slide-level ground truth. As above, often a range of probability thresholds are tried, and the one which yields the largest AUC on the validation set is then applied to the test set to give the final AUC. AUC-ROC can also be plotted as a curve. (see **Procedure steps 22–23**).

#### 1.9.5 Interpretability methods

Since deep convolutional neural networks take raw image data as an input, as with other deep models, they are frequently criticised for lacking transparency, since it can be difficult to interpret what local features of the images the model learned during training (*45, 46*). To assuage this lack of clarity, some methods have been developed to visualise what local aspects of a trained image a model uses to make its predictions. These so called ‘attention’ methods, or discriminative localisation methods, reveal what local aspects of images a model focuses its attention on to classify it into a certain class (*47*).

One of the most popular attention methods for interpreting trained deep CNNs is the class activation mapping (CAM) methods, which produce course localisation maps highlighting the regions of an image that were most important for the model’s prediction of that image (*48, 49*). This popularity extends to tile-based WSI deep learning studies, many of which from the last few years have included CAM analyses of their tiles after training to verify or discover which regions of a tile the model learned to be relevant for the classification task (*50–53*).

Among CAM methods, the Gradient-weighted CAM (Grad-CAM) method of Selvaraju et al is among the most frequently used in studies that train deep CNNs on WSI tiles. Grad-CAM uses the gradient of a target concept (for example, a class) as it passes to the last convolutional layer of an architecture to produce its localisation maps. Grad-CAM is unique among CAM methods in that it is generalisable to many different kinds of CNNs so is usable across a wide range of frequently used architectures (*49*). Due to their its architecture-agnostic design, GradCAM heatmaps can be implemented with relatively minor additions to PyTorch or TensorFlow model code.

### 1.10 Related methods

Several tools have been developed which also provide support for some of the functionalities available in PathML. Providing a complete description and comparison of these tools is beyond the scope of this protocol. Here we briefly describe these tools and direct the reader towards the articles presenting them for more details.

HistoQC (*54*) is a Python-based tool for performing quality control of WSIs, aiding users in the identification slides containing potential technical artefacts and affected by batch effects. By providing the user with modules for performing a wide range of classical image analysis techniques, HistoQC enables the construction of custom pipelines for performing foreground filtering, detection of slide artefacts such as pen marks, and identification of batch effects such as slides with darker staining compared to the rest. In HistoQC, this is achieved using a combination of approaches including inspection of colour distributions, application of edge and smoothness detectors, and classical filters such as Gabor and Frangi filters for texture analysis. For example, if the background of a WSI is uniformly white, by applying a threshold to the colour distribution which excludes white pixels can be used to perform foreground filtering. Similarly, a bright green pen mark may be clearly distinguishable from tissue by inspection of the green colour distribution of the WSI. In addition, HistoQC provides an interactive user interface for exploring one’s data. These approaches can achieve competitive results when carefully tuned by the user, but may struggle in more complex cases, such as uneven background, and pen marks with similar colour to the tissue. HistoQC is therefore a useful tool, complementing the wider functionality of robustness of PathML, and enabling rapid quality control processing of one’s data.

HistomicsTK (*55*) is a Python library for performing a number of image analysis tasks specific to WSIs including stain colour deconvolution, normalisation and augmentation, as well as cell/nuclei segmentation and even a user interface for manual annotation of WSIs. Like HistoQC, all image analysis techniques are performed using classical approaches. HistomicsTK is highly complementary to PathML, and in particular, we envisage that users may make use of HistomicsTK for performing WSI-specific colour augmentations within a PathML workflow.

Histolab (*56*) is a Python library combining features found both in HistoQC and HistomicsTK, including functions for performing classical image analysis techniques to facilitate tissue detection and artefact removal, cell/nuclei segmentation, and colour transformations such as colour deconvolution. In addition, Histolab, like PathML, supports the extracting of tiles from WSIs, and enables one to easily test alternative tiling strategies, including random extraction of tiles according to tissue detection score thresholds.

Unlike PathML, none of these tools implement deep tissue detectors, nor do they implement tools for importing annotations for WSIs to facilitate labelling of tiles or generation of tile-level segmentation masks. Furthermore, while all of these tools provide support for the preprocessing of WSIs, none provides tools for model evaluation and post-processing, such as the stitching together of tile-level segmentation masks to produce a slide-level mask.

### 1.11 Limitations

PathML is a library that was made to be as intuitive and user-friendly as possible, but certain functions can take a be time intensive or memory intensive, particularly if many tissue-dense slides are used for segmentation tasks. In particular, `Slide.inferSegmenter()` will add an array of pixel-level predictions to each suitable tile in the `Slide`. This can result in .pml files that are over 10 gigabytes for large WSIs. If float-level precision isn’t required for segmentation predictions, setting the `dtype` argument of `Slide.inferSegmenter()` to `int` to can mitigate this issue by scaling predictions to integers from 0 to 255 to produce a much smaller .pml file. Furthermore, computing the Dice coefficient for each tile in a large `Slide` with `Slide.segmenterMetricAtThreshold()` can take tens of minutes per slide. Generating a WSI-sized pixel-wise segmentation inference array with `Slide.getNonOverlappingSegmentationInferenceArray()` also required tens of minutes and up to 30 gigabytes of RAM for larger `Slide` objects. It should also be noted that the deep tissue detector built into PathML currently does not support IHC images.

### 1.12 Materials

#### 1.12.1 Equipment

##### Hardware

It is recommended that PathML users either work on a machine with at least 32 GB of RAM and enough disk space to hold the number of number of tiles they would like to extract (tiles are not large, but if thousands are extracted, a proportional amount of disk space is required. At least four cores are recommended. WSIs tend to be 0.5–5 GB in size, so if tens or hundreds are used in an analysis, disk space to store them is required (external hard drives work well). Users that wish to utilise the inference functions of `Slide`, `Slide.inferClassifier()` and `Slide.inferSegmenter()` are highly recommended to have a CUDA-compatible Graphics Processing Unit (GPU). PathML also works well in high performance computing environments that meet these conditions.

##### Software and data

- A whole-slide image viewer with support to extract annotations into either ASAP-compliant .xml format or GeoJSON format; we recommend ASAP (https://co mputationalpathologygroup.github.io/ASAP/) or QuPath (https://qupath.github.io/). Only necessary if you intend to generate or modify any annotations.
- Python version 3.7 or above ( https://www.python.org/downloads/)
- Anaconda (recommended, https://www.anaconda.com/products/indivi dual)
- The PathML Python library and associated dependencies ( https://github.com/m `arkowetzlab/pathml`)

#### 1.12.2 Equipment setup

##### PathML installation with its dependencies

- Install Anaconda (follow instructions at https://docs.anaconda.com/anacon `da/install`)
- Clone the PathML GitHub repository: 

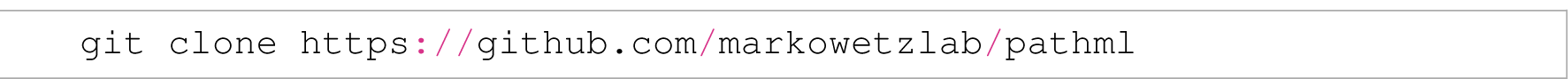
- Install PathML’s dependencies by creating an Anaconda environment using the `pathml-environment.yml` with the following command (including the path to the `pathml-environment.yml` file in the PathML repository that was just cloned) and then activate that environment: 

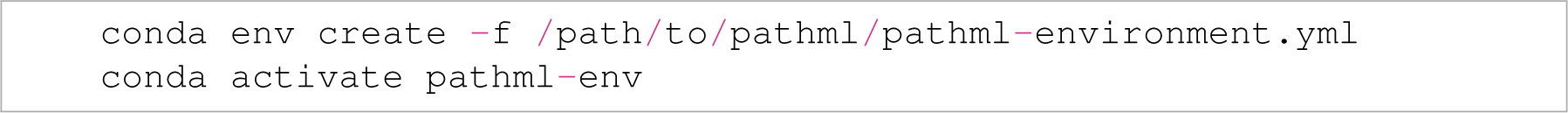 Please note that `pathml-environment.yml` installs Python version 3.7, PyTorch version 1.4, Torchvision version 0.5, and CUDA version 10.0. Stable versions above these should also work as long as the versions are cross-compatible. Be sure that the CUDA version matches the version installed on your GPU; if not, either update your GPU’s CUDA or change the `cudatoolkit` line of `pathml-environment.yml` to match your GPU’s version before creating your `pathml-env` environment.

##### (Optional)#Procedure tutorial preparation

- Clone the PathML tutorial GitHub repository: 

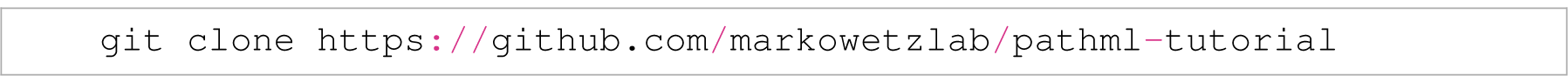
- Create a directory called `wsi_data` where there is at least 38 GB of disk space. Download the following 18 WSIs from the CAMELYON16 dataset into `wsi_data` (downloadable in the `tumor` and `normal` folders at the following link: https://drive.google.com/drive/folders/0BzsdkU4jWx9Ba2x1NTZhdz `Q5Zjg?resourcekey=0-g2TRih6YKi5P2O1SiBB1LA`): 

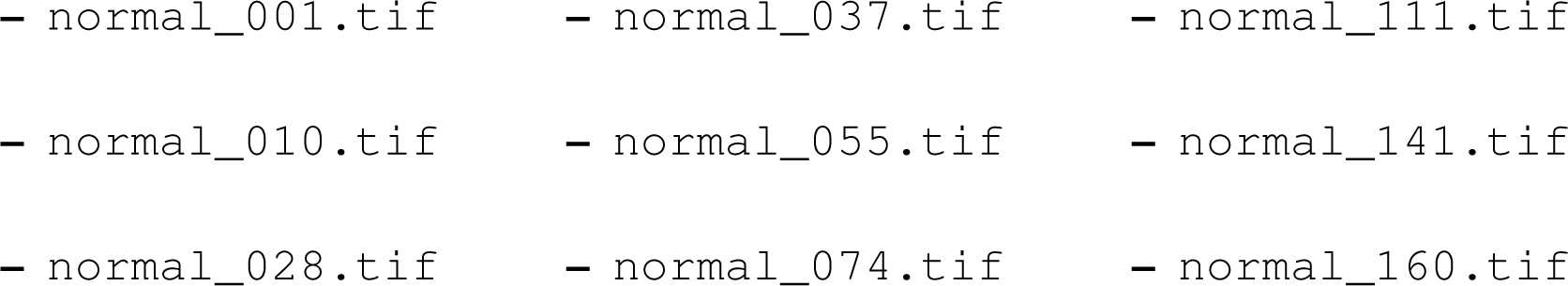
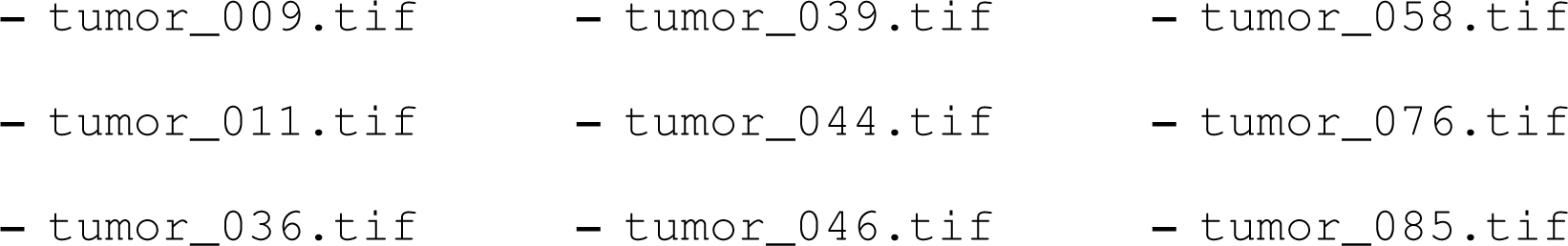
- If running the tutorial using the Jupyter notebook in the `pathml-tutorial` repository (`pathml-tutorial.ipynb`), install Jupyter notebook: 

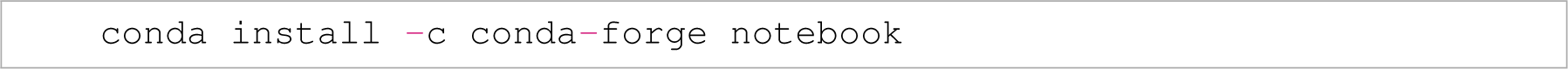
- If running the tutorial using the Jupyter notebook, start the notebook before running the code in `pathml-tutorial.ipynb` (for instructions on running Jupyter notebooks, see https://jupyter.org/documentation): 

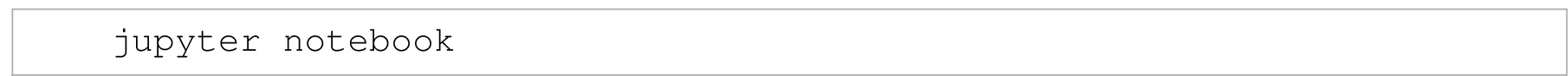

## 2 Procedure

This tutorial uses 18 WSIs and their corresponding annotations from the CAMELYON16 Grand Challenge (*57*). The WSIs show H&E stained lymph node tissue scanned at 40X. 9 of the 18 WSIs (the ones with `tumor` in the file name) contain metastatic breast cancer lesions among the lymph node tissue. The annotations, made by pathologists (*57*), demarcate these metastatic regions in a class in the .xml files called `metastasis`. The annotation .xml files also contain a class named `negative` which marks doughnut-hole regions of healthy tissue that appear inside metastatic regions. The tutorial below uses PathML to train and evaluate a model to classify metastatic from non-metastatic tiles and WSIs (**Procedure steps 1–9**) as well as to train a model and evaluate to segment metastatic from non-metastatic regions (**Procedure steps 10–36**).

Note that for the sake of the tutorial being relatively easy and quick to run as a proof of concept, only 18 of the 400 CAMELYON16 WSIs are used, with 6 WSIs in the training set, 6 in the validation set, and 6 in the test set (3 metastasis-positive slides and 3 metastasis negative WSIs were selected for each set). In practice, though, it is recommended that users make use of all of the data they have available to them to train more robust models.

The code below is reproduced in a Jupyter notebook, `pathml-tutorial.ipynb`, in the `pathml-tutorial` GitHub repository (https://github.com/markowetzlab/ `pathml-tutorial`). The complete results of a full run of the code below can be also be found in the repository.

### Preparation of programming environment and working directories and defining patient-level train-val-test split

1. Import PathML and its dependencies. 

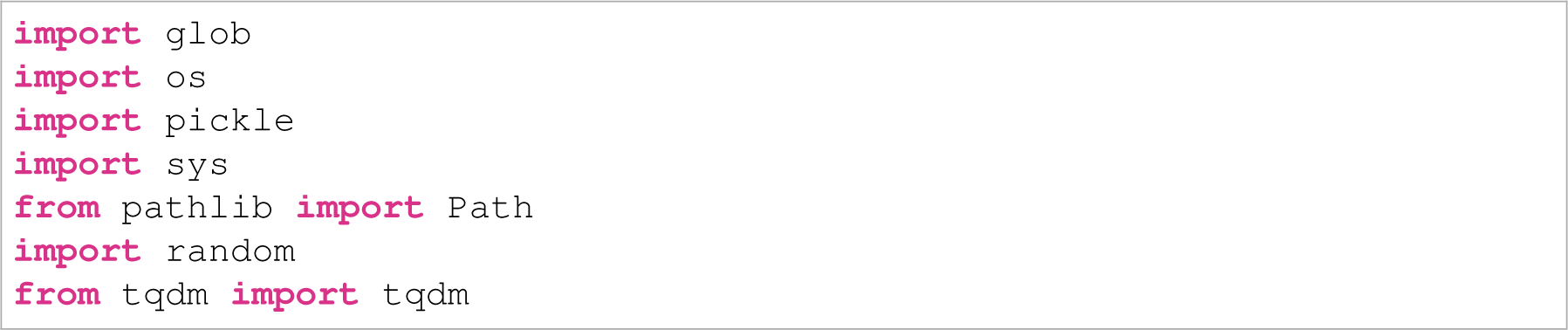 Next, append PathML’s directory path to the system path, then import PathML and essential libraries for training and evaluation of model performance. Make sure to substitute the path to your local PathML installation in place of ’/path/to/pathml’. 

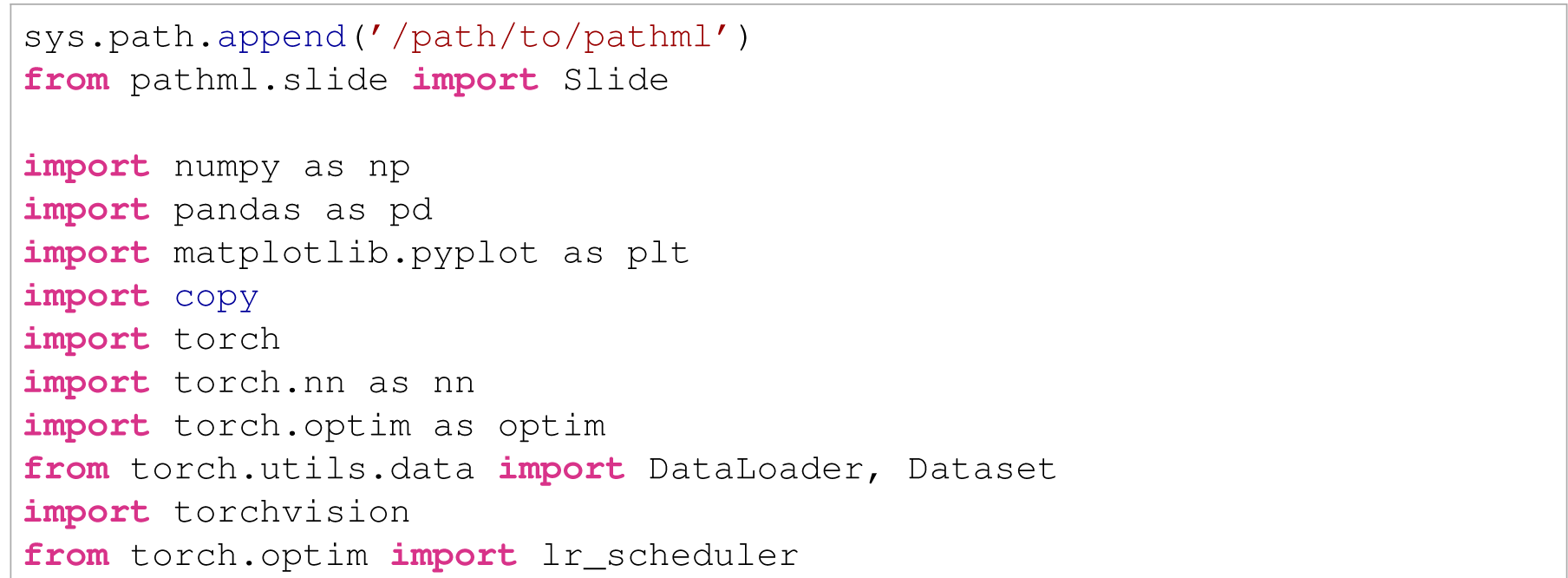
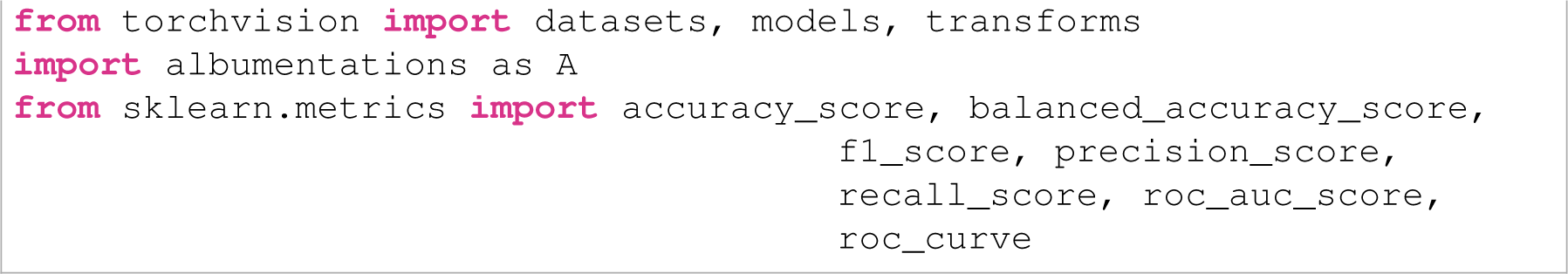
2. Prepare directory in which analysis will be performed and specify path to directory containing WSI data. The analysis directory should contain a `pathml_slides` subdirectory for saving of PathML slide objects. The analysis directory should also contain an `annotation` directory where annotation files (either .xml or .json) should be stored with the same names as the whole-slide images they belong to (e.g. `normal_001.xml` is the annotation file corresponding to the WSI file `normal_001.tif`). If following this tutorial, the `pathml-tutorial` directory cloned from GitHub (https://github.com/markowetzlab/pathml-tutorial) can serve as the analysis directory (`annotation` directory is included, but WSIs need to be moved into a `wsi_data` directory (see **section 1.12.2**). WSIs should all be stored in a single `wsi_data` directory, which does not need to be a subdirectory of the analysis directory (see **section 1.12.2**). 

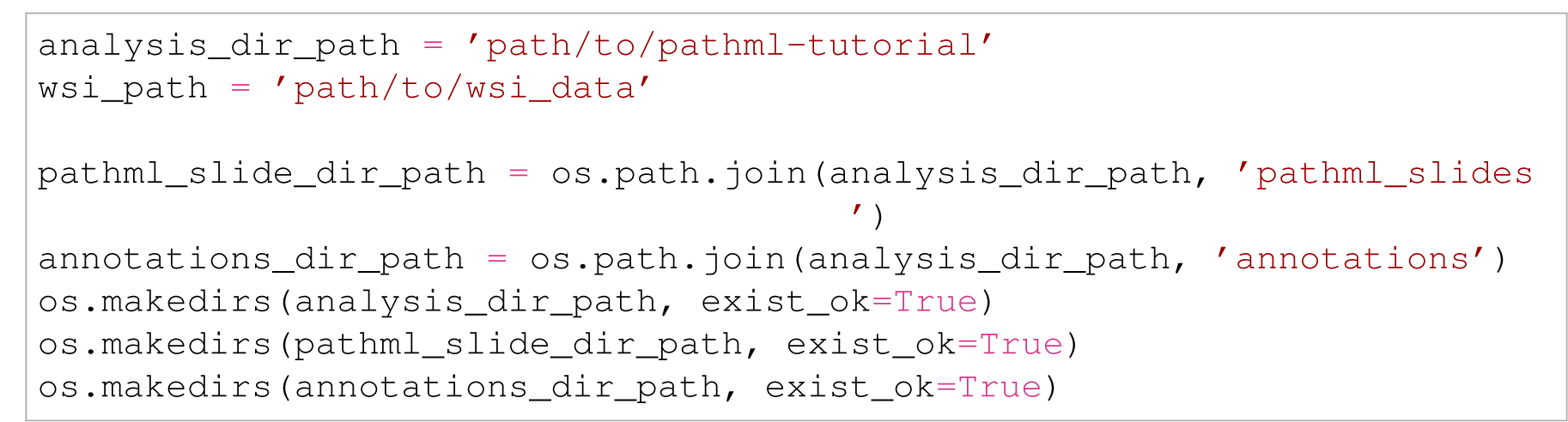
3. (Optional) If training models to perform more than one task, create results subdirectories within the analysis directory for outputs from each, e.g. separate classification and segmentation directories. 

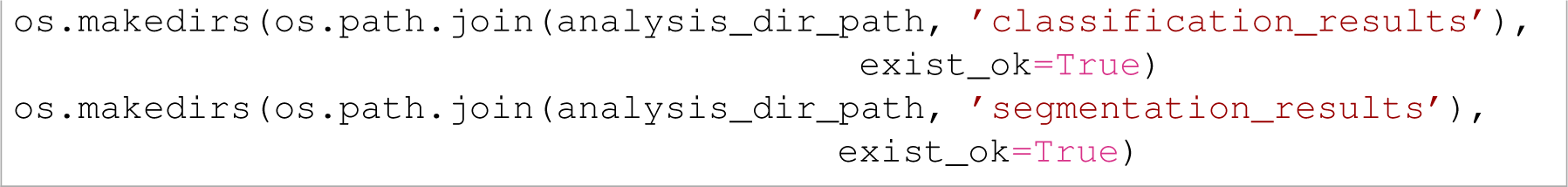
4. Define patient-level split for training, validation, and test sets. We create separate Python lists for the simple patient IDs (e.g. ‘normal 001’) and for the full paths to those wholeslide images. Please be aware that we use a fixed split for the purposes of this protocol, but users should create a random patient-level split for their own data.

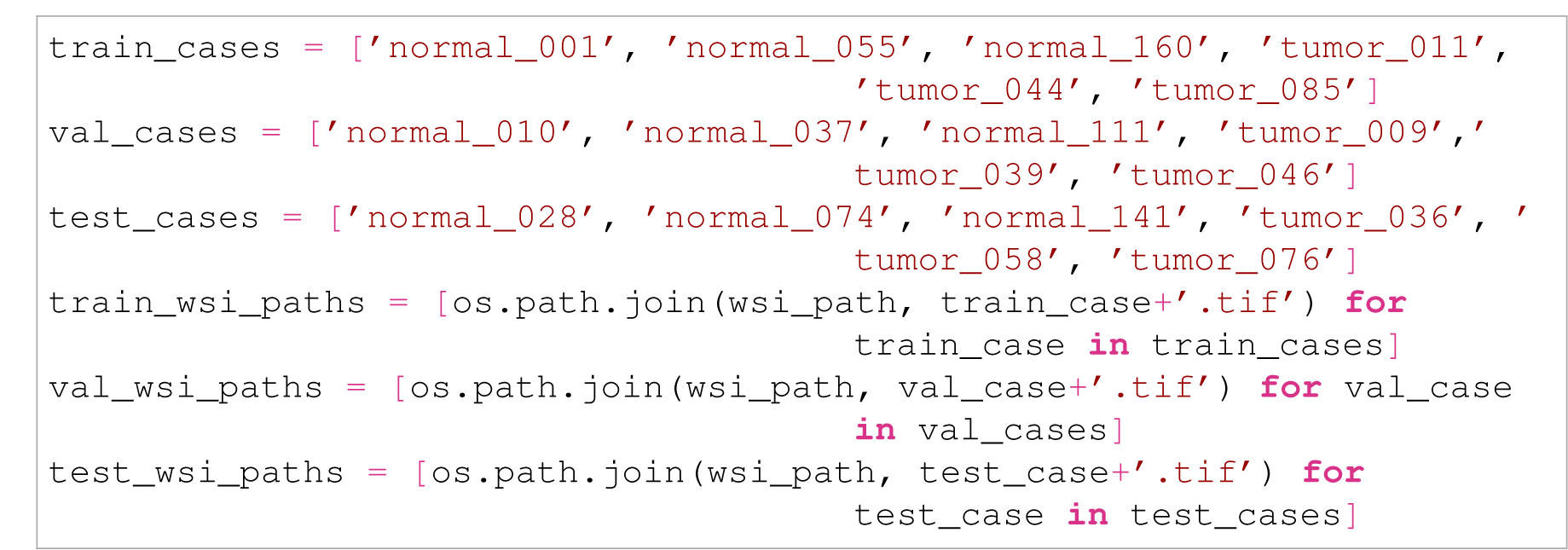

### Initialise PathML Slide objects, filter out non-tissue regions, and add annotations

5. Use a for loop to iterate through the file paths to each WSI. Inside the loop, get the patient case from the WSI path and initialise a PathML `Slide` with the WSI path, setting the `level` to 0 to work at the highest magnification of the pyramidal image, and using `Slide.setTileProperties()` to define our tile size edge length in pixels. Next, call `Slide.detectTissue()` to infer PathML’s deep tissue detector on the `Slide` and `Slide.detectForeground()` to apply simple foreground filtering methods. We can choose specific parameters for filtering using these methods later, after running these functions. If the case is a tumour case rather than a normal case, then find the matching annotation file in our `annotation` directory and use `Slide.addAnnotations()` to parse these annotations into the `Slide` object. If there are negative spaces in the geometry of the annotations (e.g. doughnut holes), set the `negativeClass` argument of `Slide.addAnnotations()` to be the name of that negative class in the annotation file. Finally, call `Slide.save()` to preserve our current `Slide` object as a .pml file in our `pathml_slides` directory for re-loading and re-use later. 

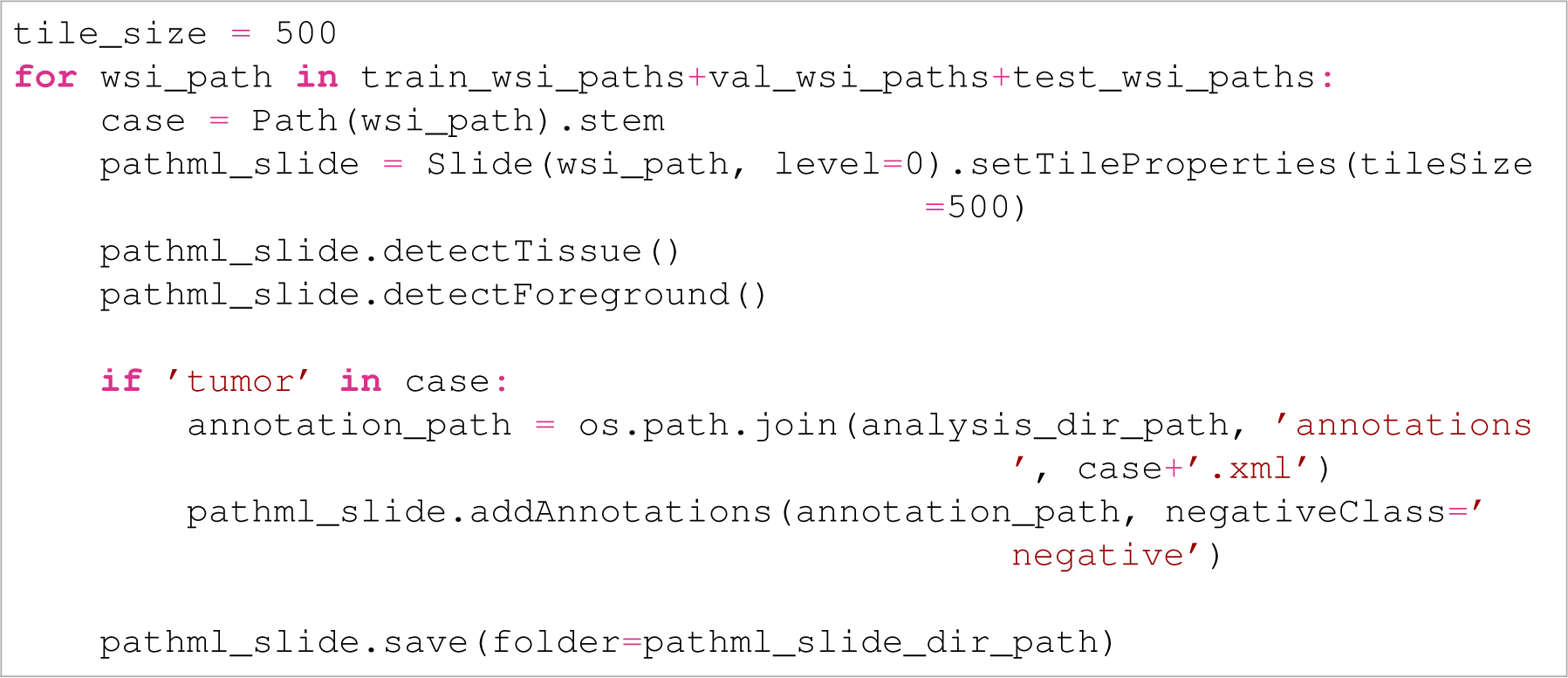
6. Load a `Slide` that was just created using `Slide()` (the initialiser) and including the path to a .pml file as argument. Then, use `Slide.visualizeThumbnail()` to portray a small, low-resolution image of the WSI. Then call `Slide.visualizeForeground()` twice, once while setting the argument to `otsu` to show the foreground as filtered using Otsu’s method, and once setting the argument to `triangle` to show the foreground as filtered using the triangle algorithm. Note that the argument can also be set to an integer from 0 (black) to 100 (white) to give a manual intensity cutoff which will consider all tiles below or equal to that integer to be part of the foreground. Finally, call `Slide.visualizeTissueDetection()` to show the background, artefact, and tissue regions detected by the deep tissue detector. Compare these visualisations to determine which method or methods of filtering works best for the application. It may be necessary to visualise many slides to get a good sense of which filters to use. Here, we visualise all six of our validation slides, saving the results to the `classification_results` directory:

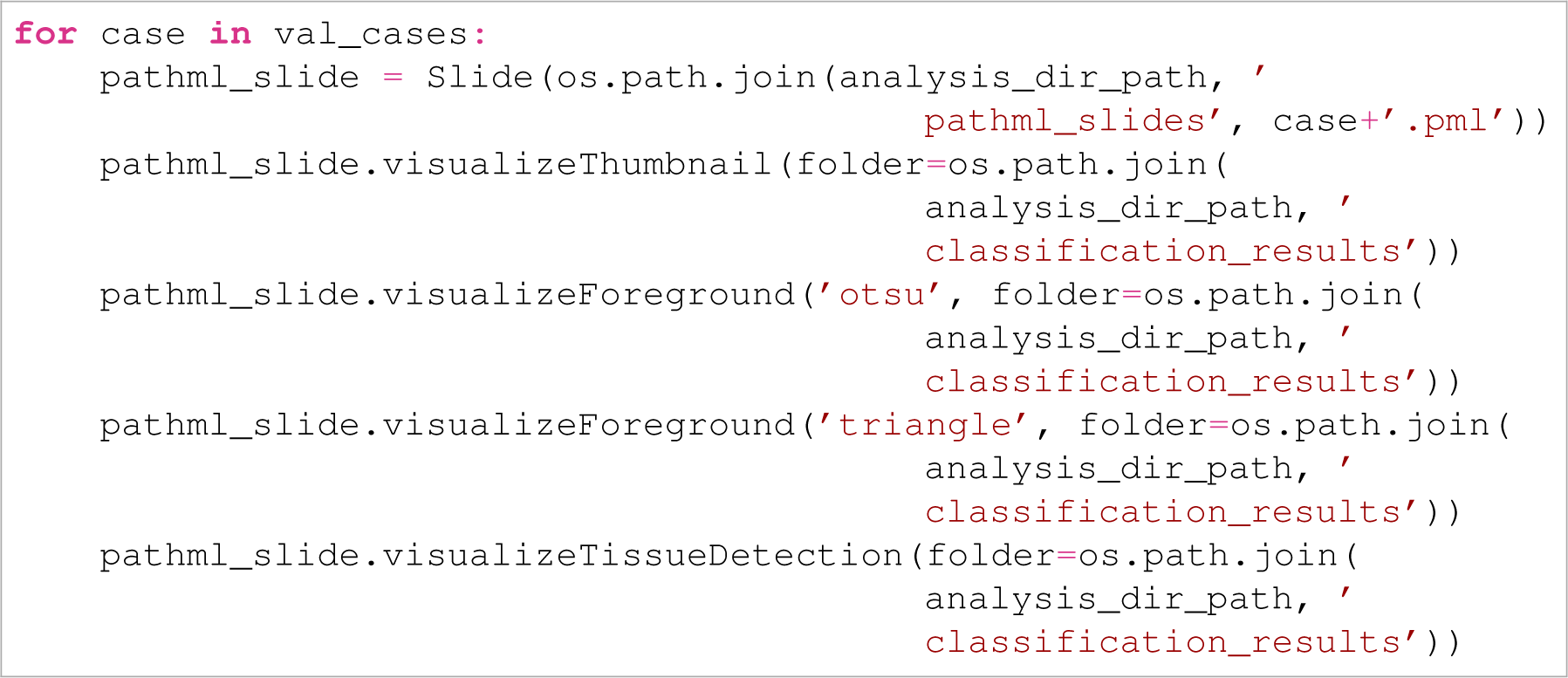

### Extracting tiles and masks from Slide objects

7. Iterate over all training and validation cases, extracting tiles that overlap the annotations added and their corresponding binary masks using `Slide.extractAnnotationTiles()` for `Slides` that have annotations added to them, and `Slide.extractRandomUnannotatedTiles()` for `Slides` that do not have any annotations. For both functions, set `tissueLevelThreshold` and `foregroundLevelThreshold` to appropriate thresholds so that only tiles considered part of the tissue region of the slides will be extracted, and set `numTilesToExtractPerClass` to determine the maximum number of tiles to extract from each class present in the annotation file. If this argument is not set, all suitable tiles from each desired class will be extracted. For specifically `Slide.extractRandomUnannotatedTiles()`, set `tileAnnotationOverlapThreshold` to be a float which will be used as the minimum fractional overlap with an annotation required for a tile to be considered part of an annotation class, and therefore extractable. Set `classesToExtract` to be a list of classes in the annotation file to extract from; if it is not defined, all classes in the file will be extracted from. In both functions, `otherClassNames` can be defined as a list that will create empty tile directories for classes desired, but not present in the annotation file. A `seed` argument can be set for both functions to ensure reproducibility in the tiles extracted. Tiles are extracted into a `tiles` directory, created in the `outputDir` specified as the first argument of both extraction functions, which has a subdirectory structure (visualised in **fig. 3**) that is amenable for use in the `ImageFolder` dataset creator of PyTorch’s `torchvision.datasets`.

Binary segmentation masks for each extracted tile can be extracted in an exactly parallel directory in `outputDir` called `masks` by setting the `extractSegmentationMasks` argument in either tile extraction function to `True`.

Pixel values of 255 appear as white in these masks indicate positivity for the class, whereas pixel values of 0 appear as black and indicate negativity for the class.

Unless the `returnTileStats` argument (of either tile extraction function) is set to `False`, the function will return a `dict` containing the 0 to 1 normalised sum of channel values across all extracted tiles, the sum of the squares of channel values for all extracted tiles, and the total number of tiles extracted. These values can be used to compute the channel-wise mean and variance across all tiles in a dataset, which can be useful when performing data augmentation. Collect this dictionary for each call to `Slide.extractAnnotationTiles()` and `Slide.extractRandomUnannotatedTiles()`, cumulatively summing the channel values, squared channel values, and total tile count across all extracted tiles.

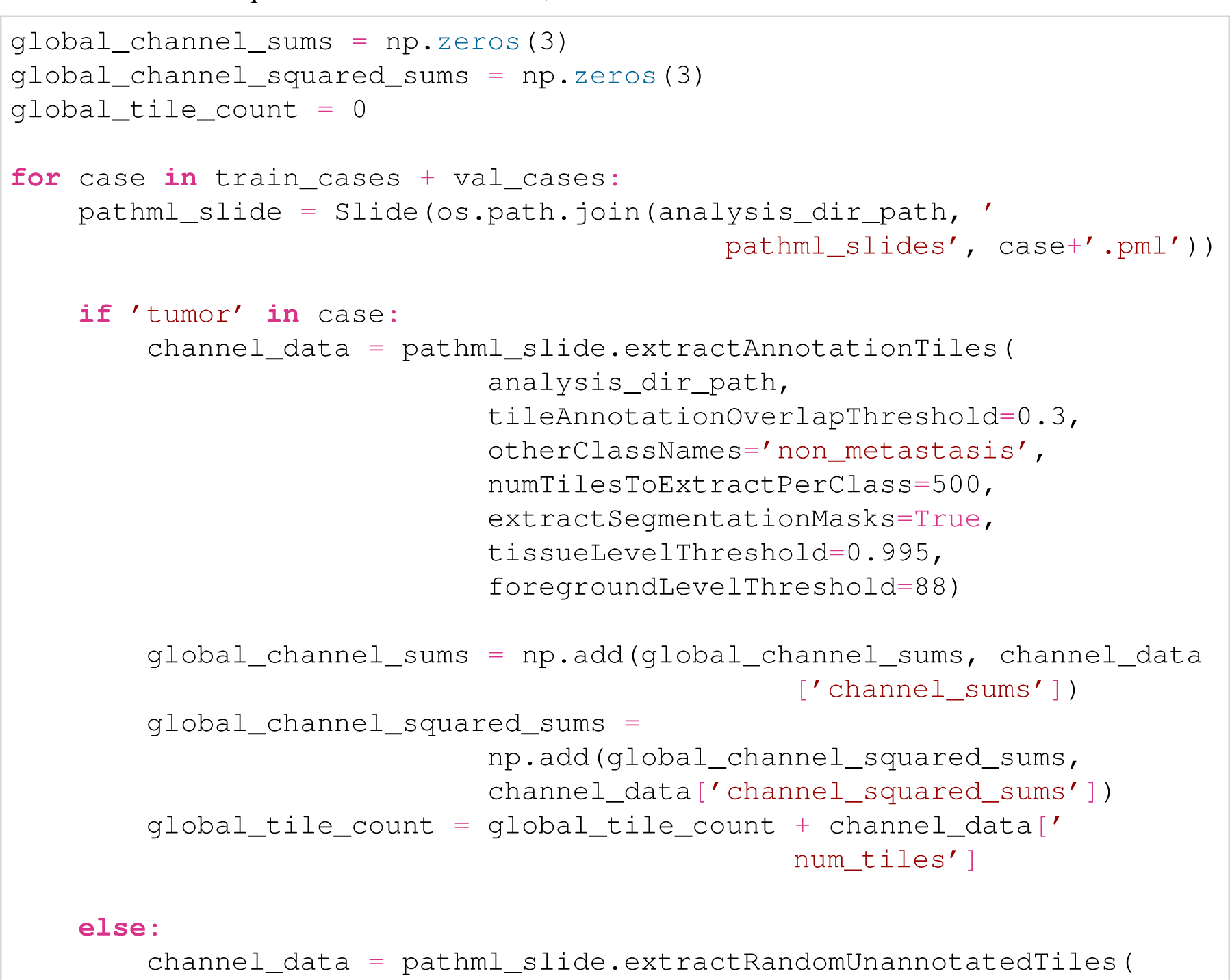
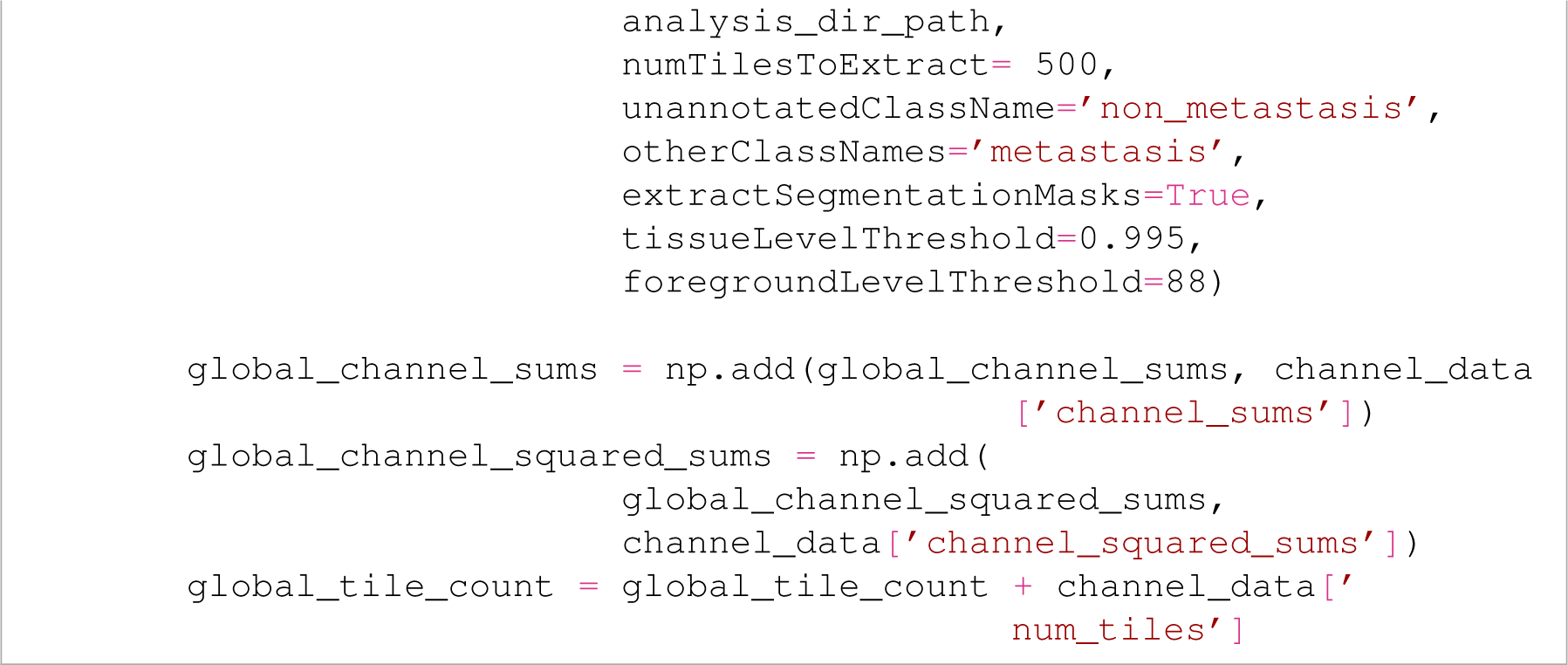

8. Compute the training and validation-wide channel-wise means by dividing the channel sums accumulated above by the total number of pixels accumulated per channel (the total number of tiles multiplied by the square of the tile size edge length in pixels). Compute the training and validation-wide channel-wise standard deviations by dividing the channel sums squared from above by the total number of pixels accumulated per channel, the subtracting from each of these channel-wise values the square of the channel means to get the channel variances. Take the square root of the channel variances to get the channel standard deviations. Save the pickled channel means and standard deviations to the `classification_results` directory. 

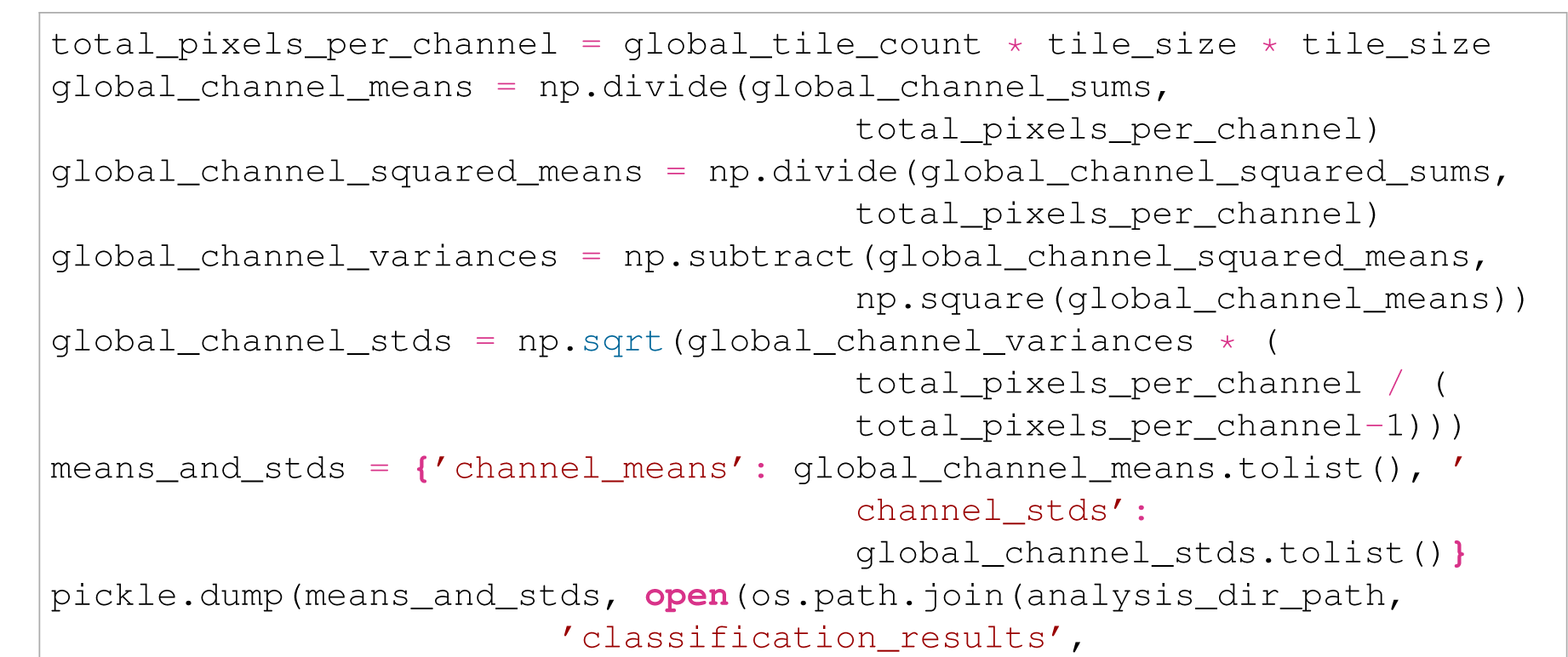
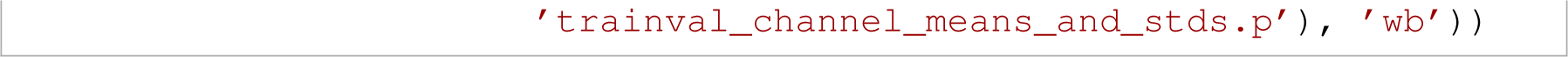
9. Count how many tiles of each class across all WSIs were extracted to ensure that the dataset is roughly class-balanced. If it is not, employ an imbalanced dataset correction technique. Here, Python’s `glob` is used to perform that task. 

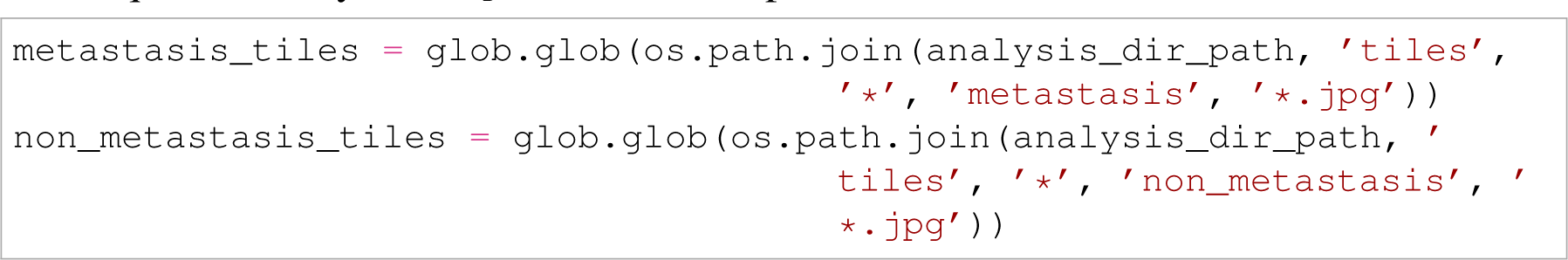

### Training a classification model

10. Define the data transforms to define the data augmentation steps for the training and validation sets before training a classification model. The channel means and standard deviations computed above can be used as a normalisation step in the augmentation data transforms. Make sure to resize to the tiles here to the necessary pixel input size of the architecture being used. Data augmentation for the validation set should be separate and should generally exclude all except the resizing and normalisation steps. 

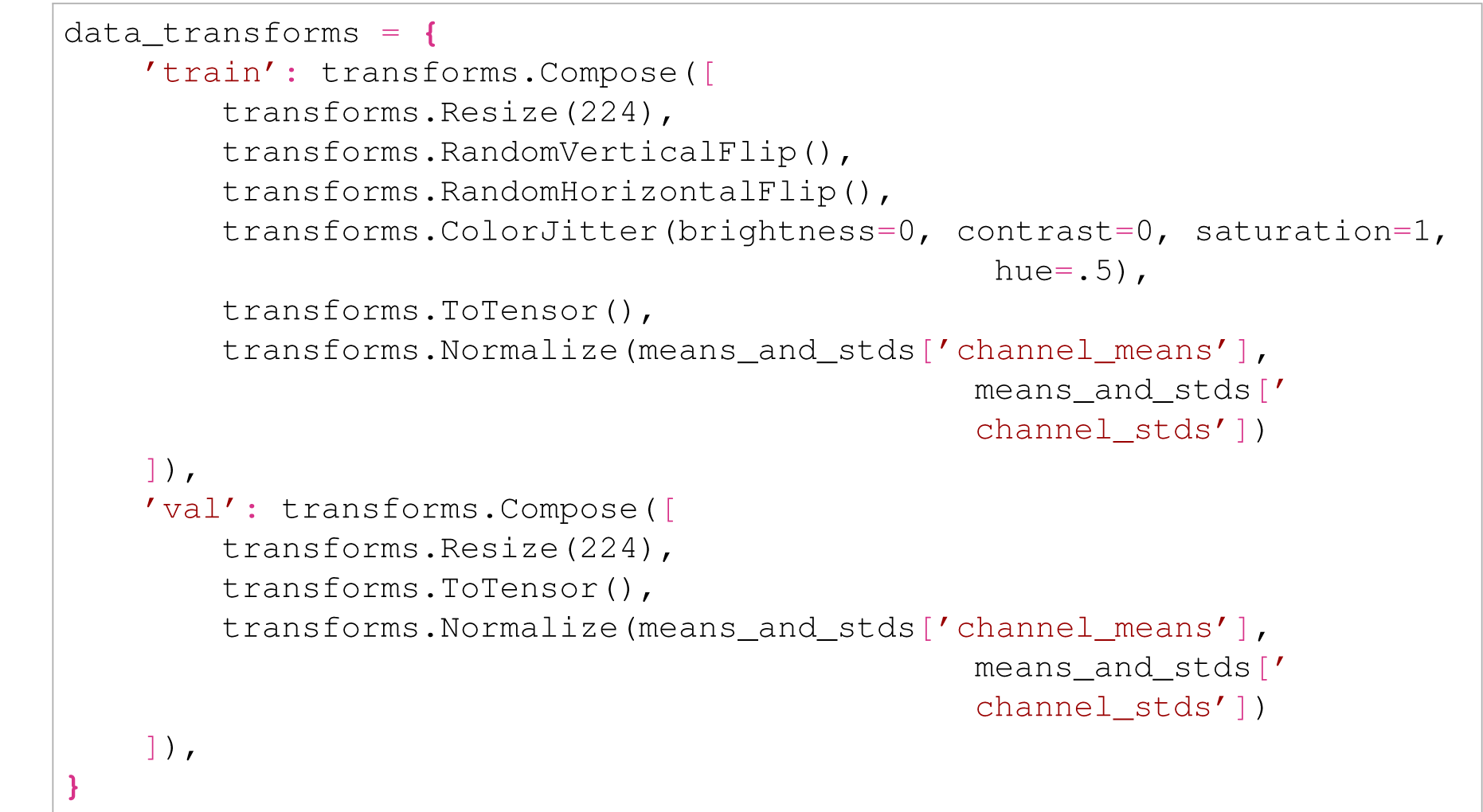
11. Define datasets and dataloaders for input into a classification model for both the training and validation sets. If using PyTorch, the `tile` directory structure outputted by the tile extraction functions above can be used to build a PyTorch dataset by inputting the path to `tile` directly to `torchvision.datasets.ImageFolder`, and then inputting the resulting dataset to `torch.utils.data.ConcatDataset` along with the corresponding data transform. Data loaders for the training and validation set can then be created from these datasets in addition to batch size and other runtime information. It is highly recommended that the data loaders be configured such that the data in the dataset are shuffled between each epoch, such as by setting `shuffle=True` in `torch.utils.data.DataLoader`. 

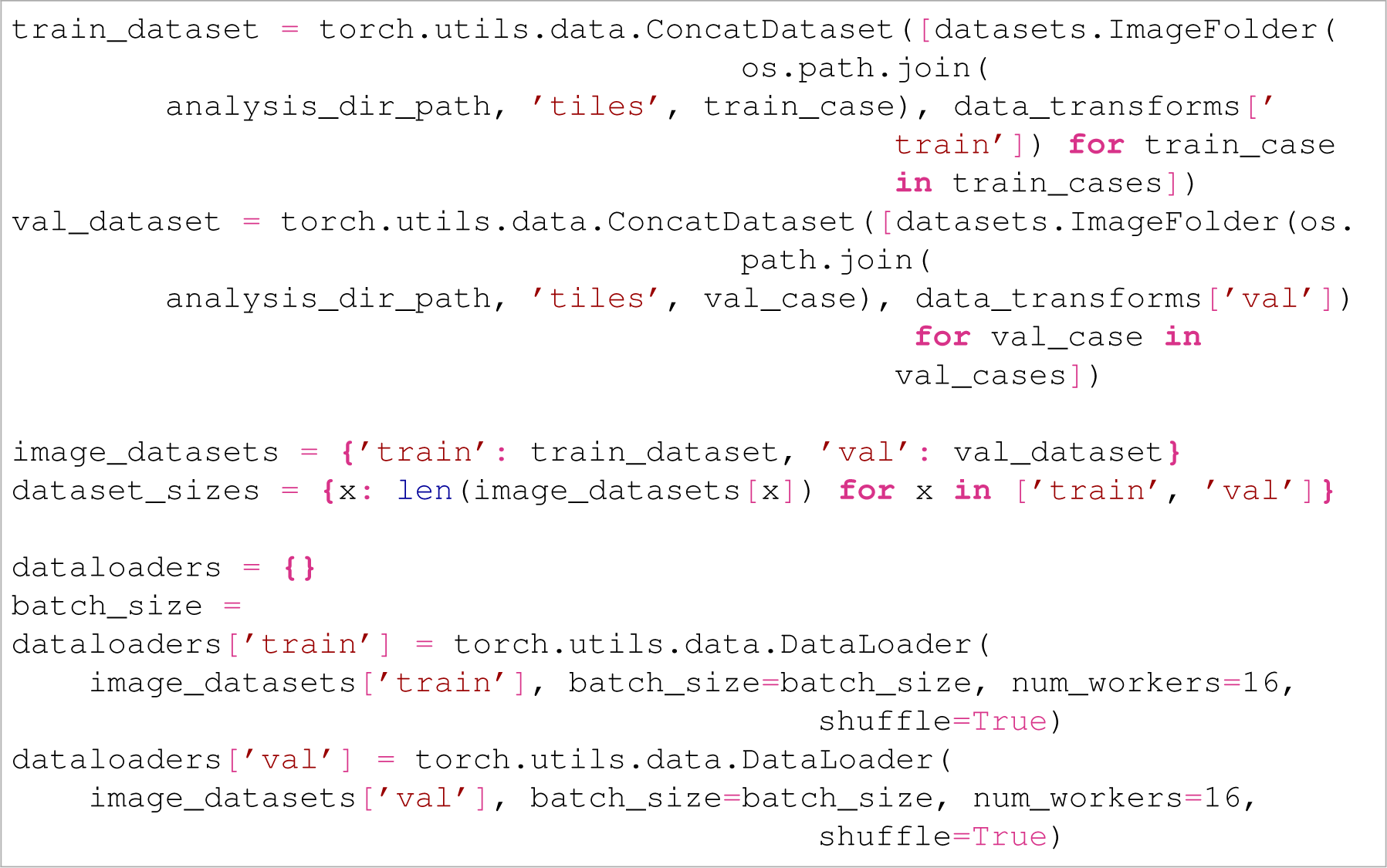
12. Visualise a batch of the training data loader to ensure that the data transforms produce tiles that are augmented in the desired ways. This code comes partially from the PyTorch documentation (*33*). 

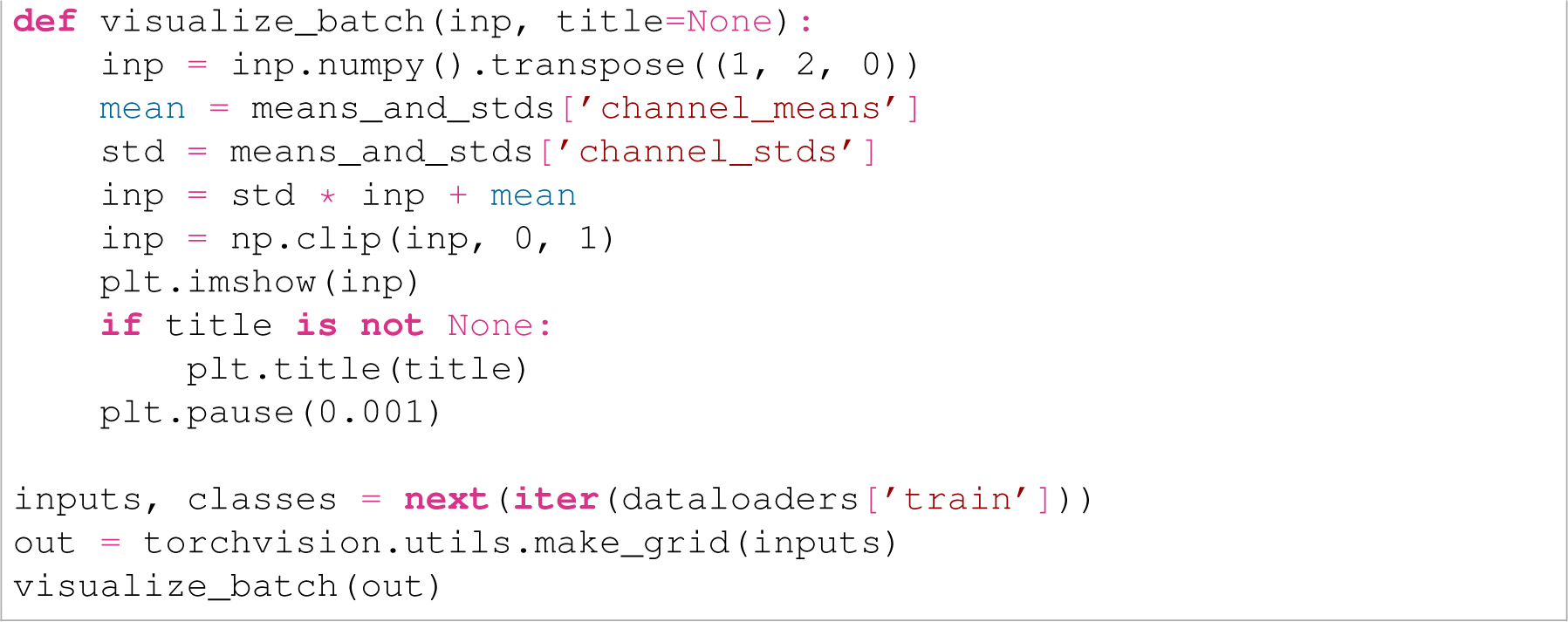
13. Define the classification model. If using a pretrained classifier, specify which pretrained weights are being used. If using a GPU, configure the model to work on the GPU (and configure parallel computation if using multiple GPUs). Define the loss function, the optimiser (including the learning rate), and the learning rate decay function (if used). 

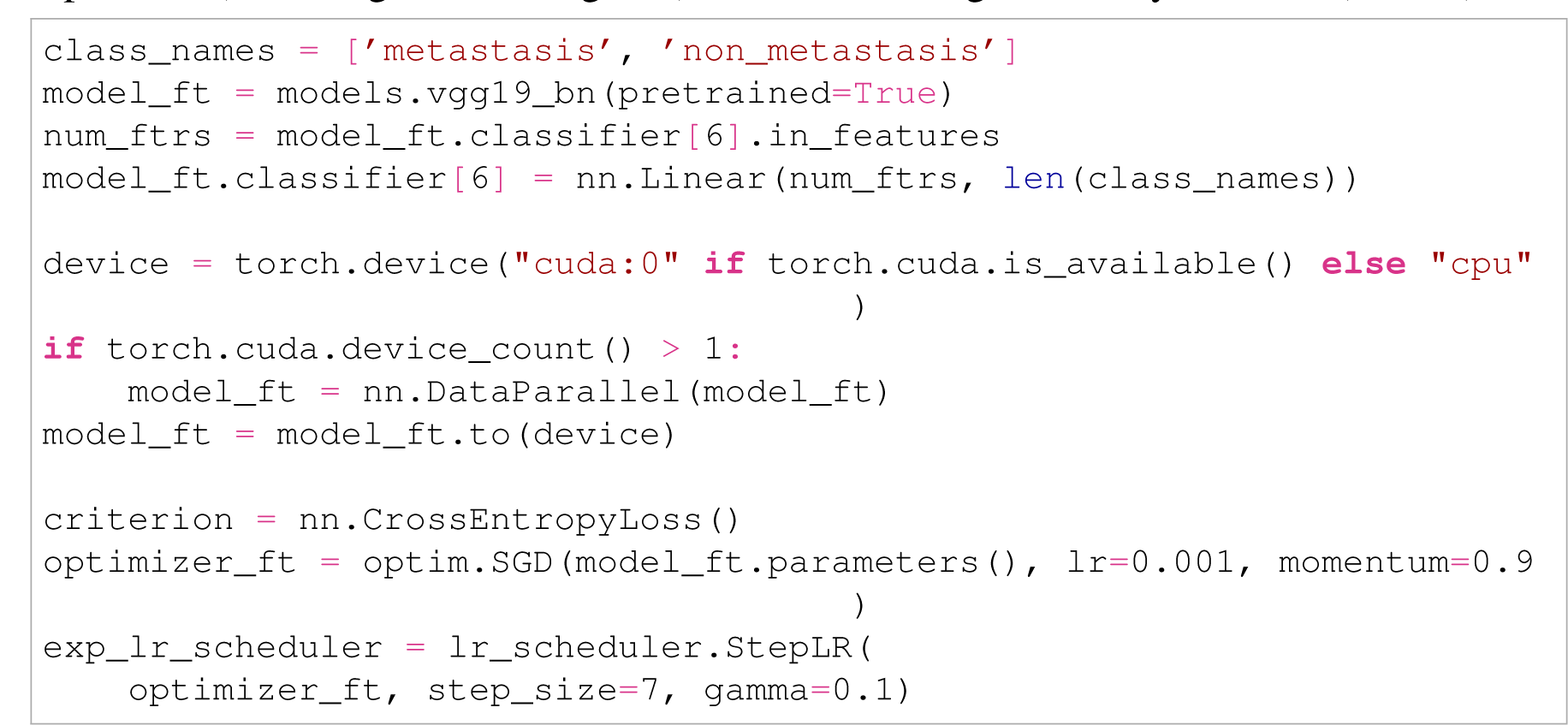
14. Construct a training function that takes as input the model, loss function, optimiser, learning rate scheduler, and number of epochs to train for and outputs the same model with the weights adjusted from training. Collect and also return the learning statistics accumulated during training, such as (at a minimum) the training accuracy, validation accuracy, training loss, and validation loss after each epoch (*33*). 

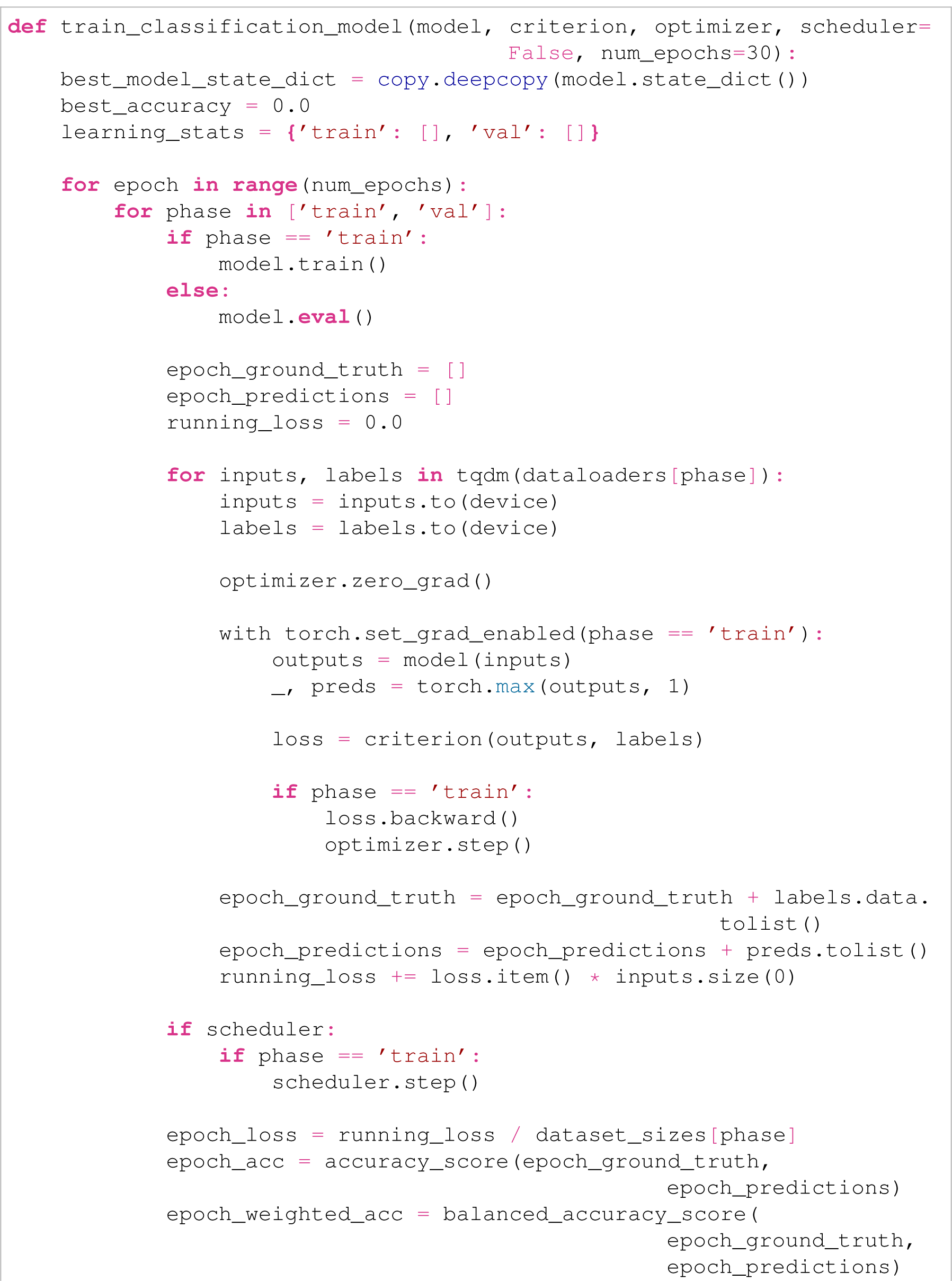
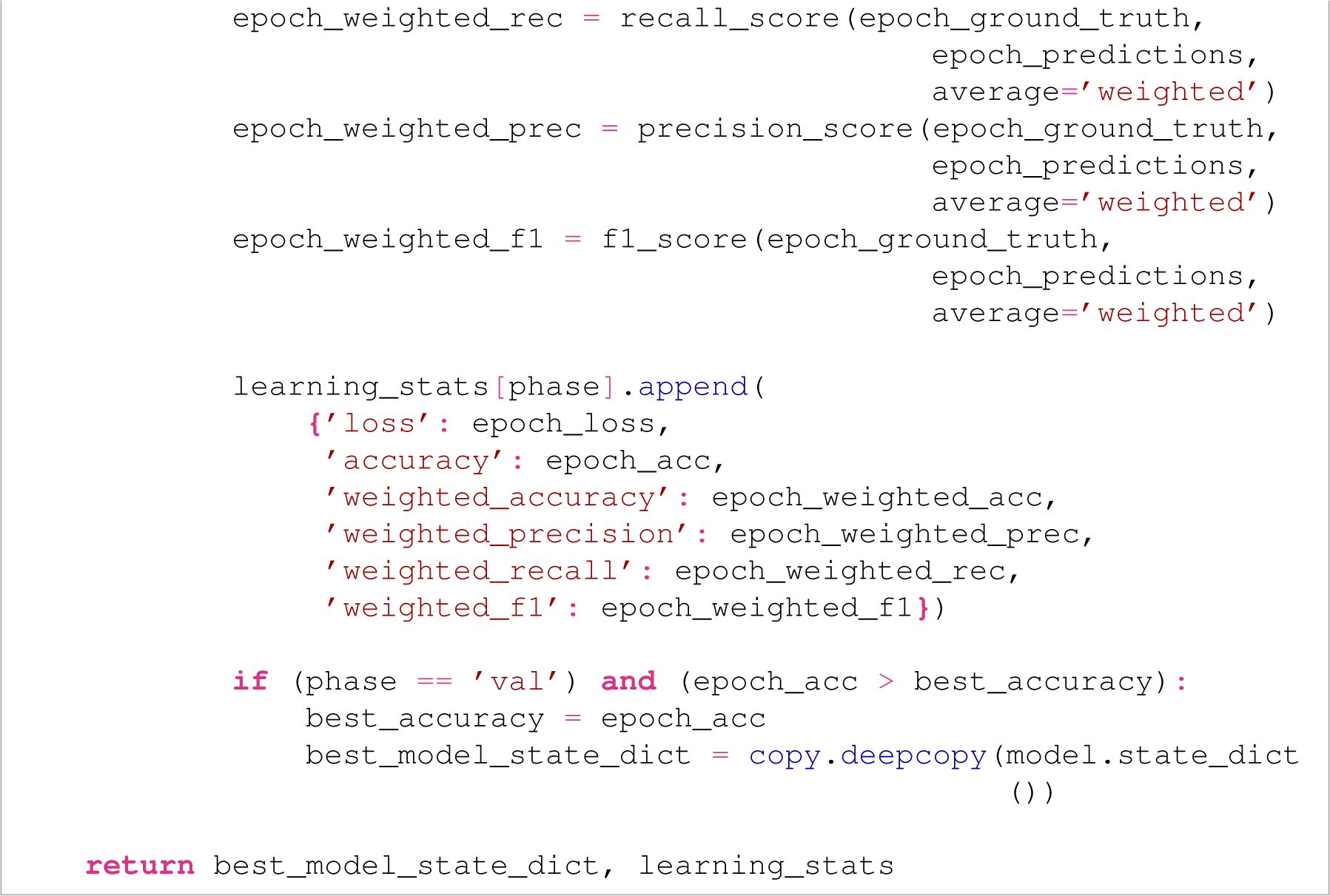
15. Run the training function. Save the learning statistics and trained model to the `classification_results` directory. If your GPU runs out of memory, reduce the batch size until it can fit an entire batch. 

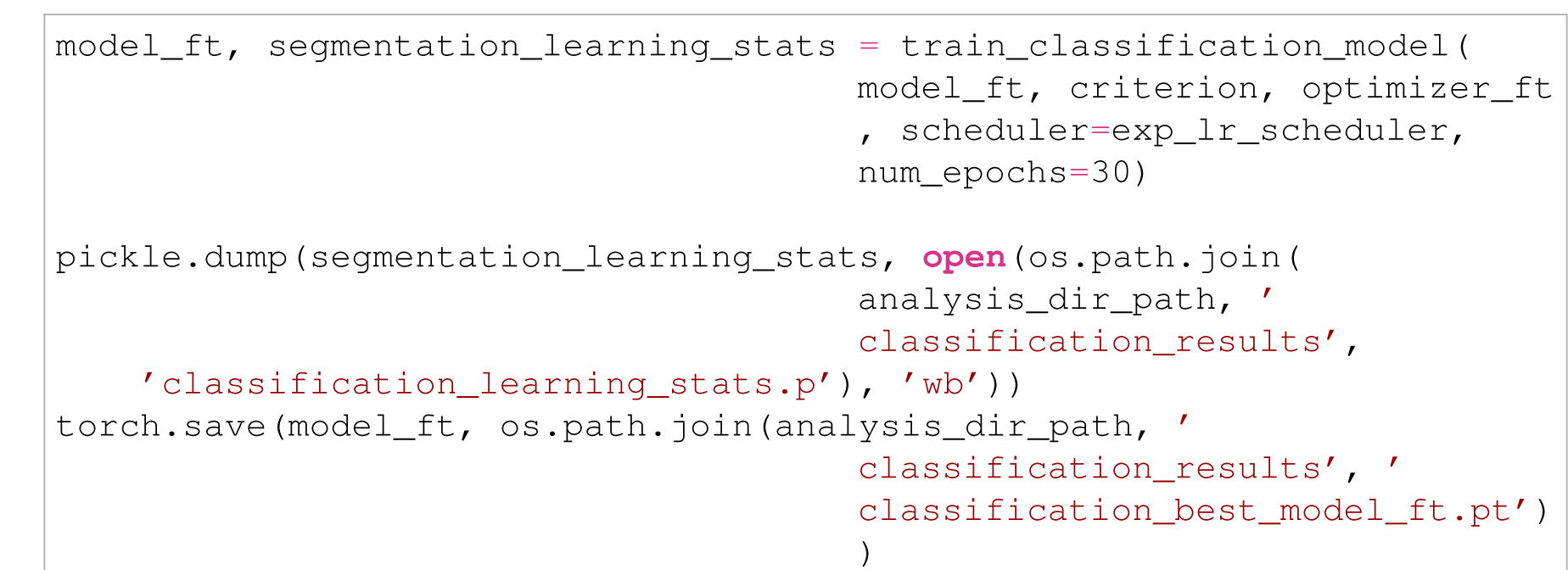
16. Produce plots showing the training and validation accuracy across training epochs, and showing the training and validation loss across training epochs.

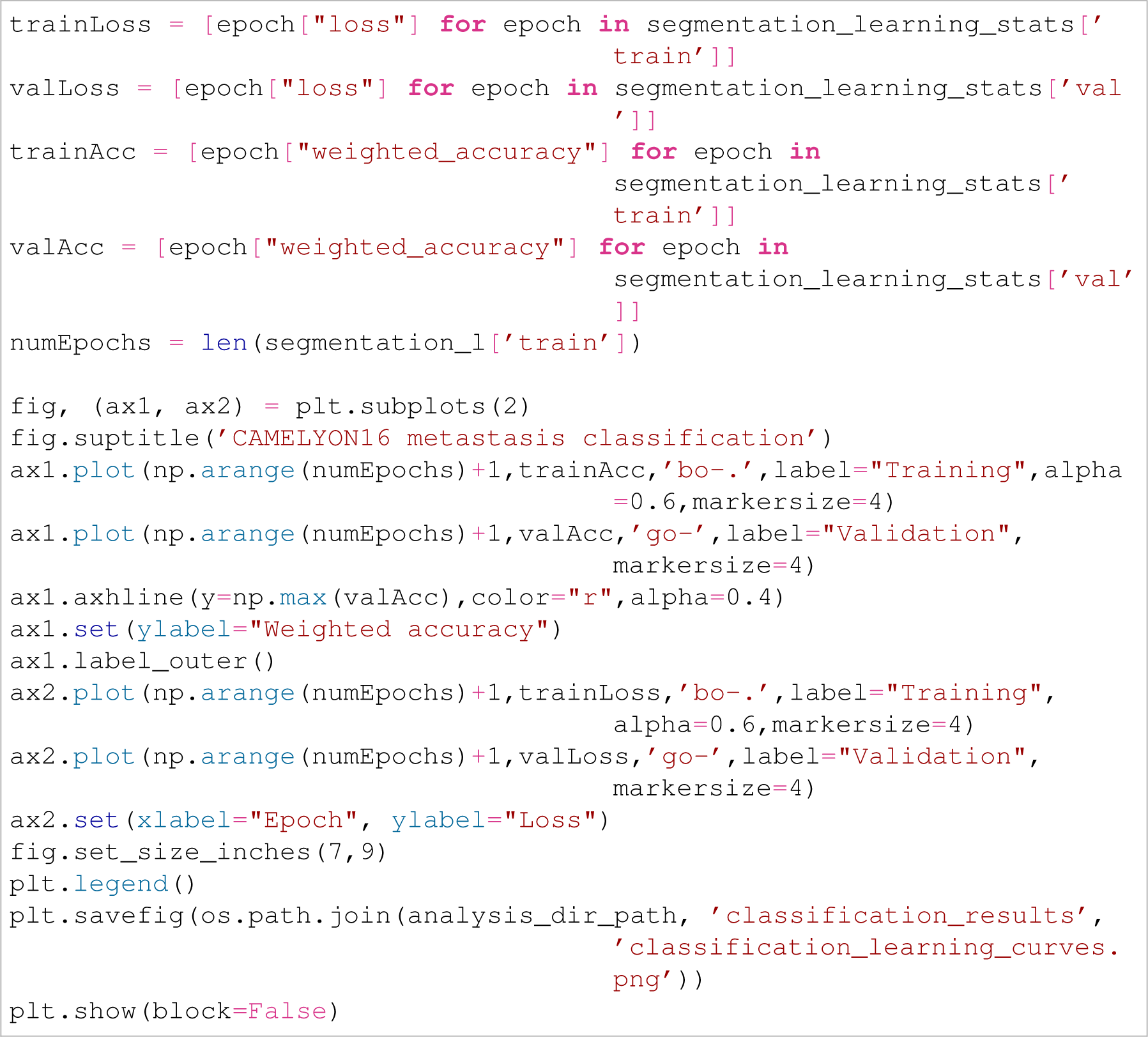

### Inferring on the trained classification model and validating model performance

17. Load the trained model and iterate over all validation and test WSIs. Load each WSI’s `Slide` object, and then infer the trained model on these WSIs by applying PathML’s `Slide.inferClassifier()` function on it, using the trained model as an argument along with the same `tissueLevelThreshold` and `foregroundLevelThreshold` values used when extracting tiles. This will ensure that `Slide.inferClassifier()` only infers on tissue tiles in these WSIs rather than wasting time inferring on artefact and background regions. Also include the same data transforms used on the validation dataset for the `dataTransforms` argument. After inferring on a `Slide`, call `Slide.save()`, using the same path to the `path_slides` directory to save these classification inference results in the same `Slide` objects. 

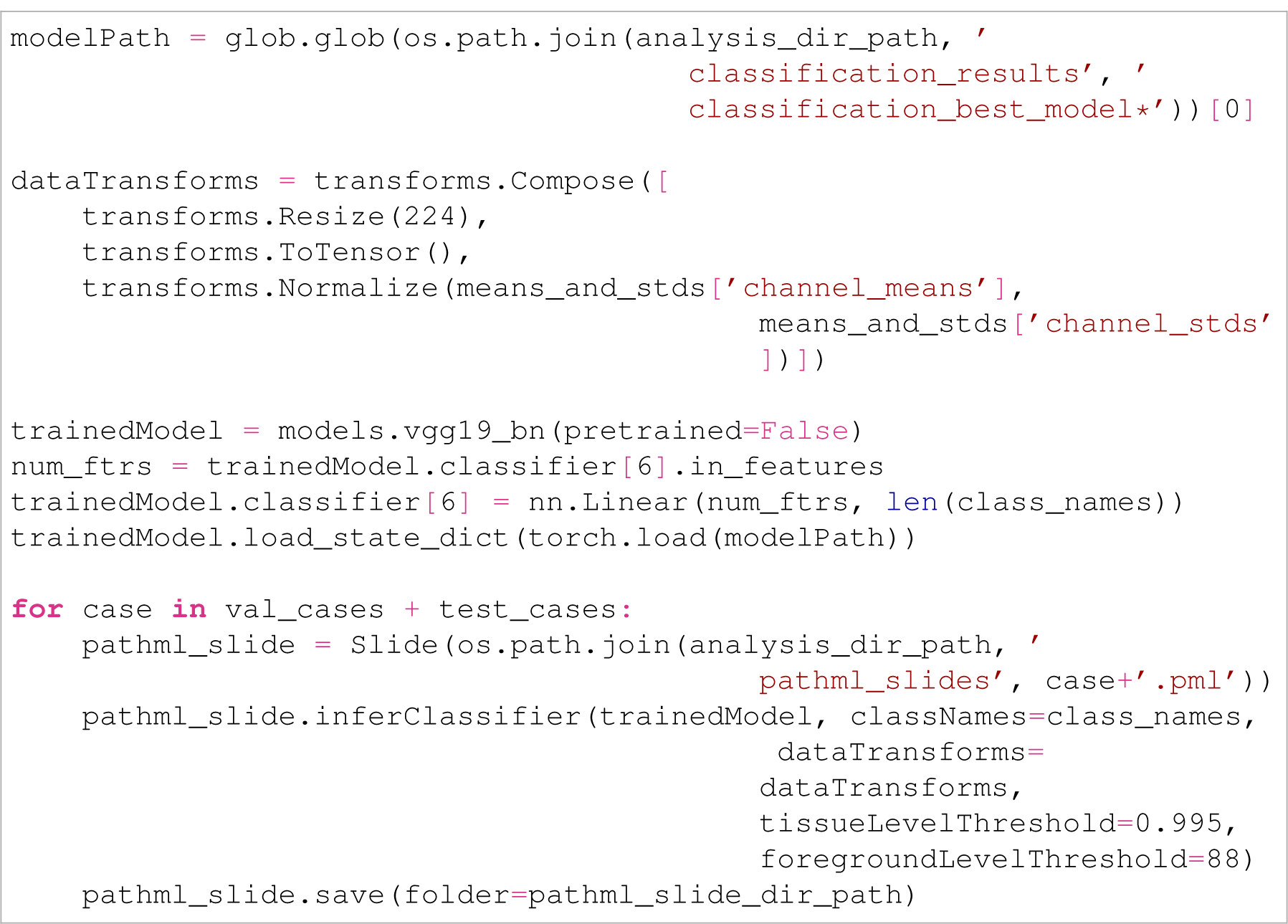
18. Load all validation set `Slide` objects and run `Slide.visualizeClassifierInference()` on them, using the name of the class of interest as argument, to generate a figure showing the classification inference results overlying that WSI in map form to ensure that the model has learned to identify that class properly. A `folder` argument can also be provided with a path to a directory to save this classification inference map to an image file. Here, we save the metastasis inference maps of all six validation WSIs to the `classification_results` directory: 

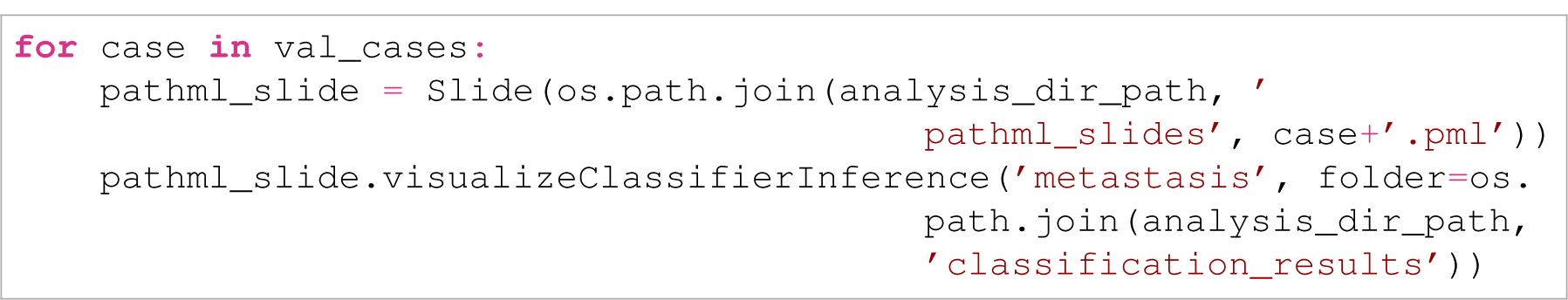
19. Define an interval of inference probability thresholds at or above which a tile will be considered a member of the class of interest. Iterate over all validation cases, loading the `Slide` object of each, then call `Slide.classifierMetricAtThreshold()` on the `Slide` object to return for that WSI the average accuracy of the classifier’s inference prediction (according to whether the classifier’s probability output is at or above the inputted probability threshold) at guessing the ground truth label of that tile for the class of interest. The name of the class of interest is used as the first argument, and the probability threshold or list of probability thresholds to check is the second argument. A tile is considered ground truth positive for the class of interest if its overlap with the ground truth annotations is above the `tileAnnotationOverlapThreshold` argument’s value. Setting the `assignZeroToTilesWithoutAnnotationOverlap` ensures that even if a `Slide` doesn’t have ground truth annotations in it from a `Slide.addAnnotations()` call, all tiles will be assumed to be ground truth negative for the class of interest. The `metric` argument is set to `accuracy` so that an accuracy operation is performed.

If a list of probability thresholds is inputted, a list of accuracies corresponding to each of those thresholds is returned. Element-wise average the accuracy list for each validation slide, then determine which probability threshold in this element-wise averaged list yields the highest accuracy for predicting the class of interest on the validation set. Note that checking more probability thresholds will give a more precise best threshold. As seen below, for this tutorial, we check probability thresholds at higher frequency the closer we get to probability 1; this is because we found in testing that for this problem, the best probability threshold for classification lied very near to 1, and so tried more thresholds closer to this number. This best validation probability threshold can then be applied to measure performance on the test set. 

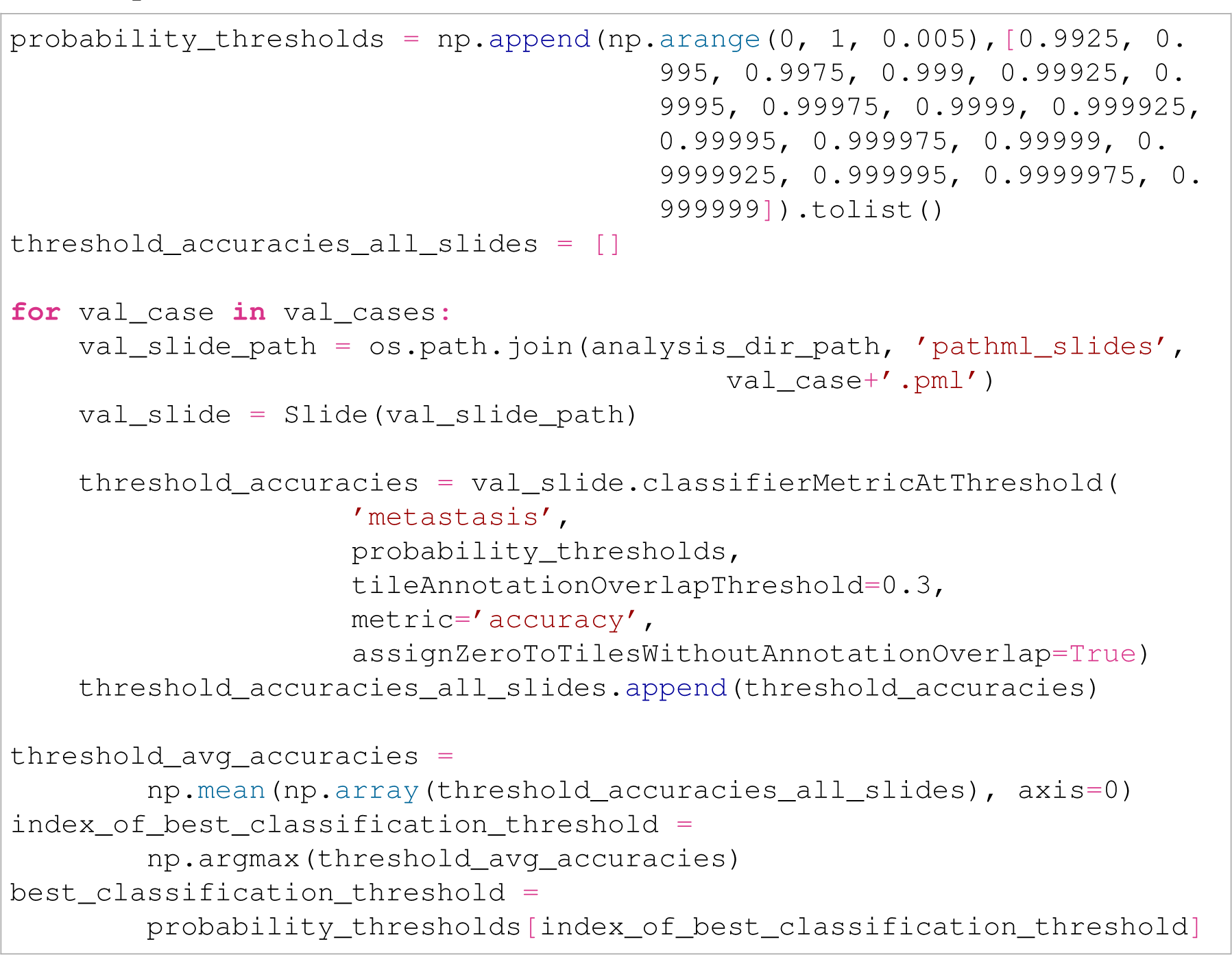

20. Plot the element-wise average prediction accuracies on the validation set against the probability thresholds used to generate them to visualise how changing the probability threshold changes validation accuracy. 

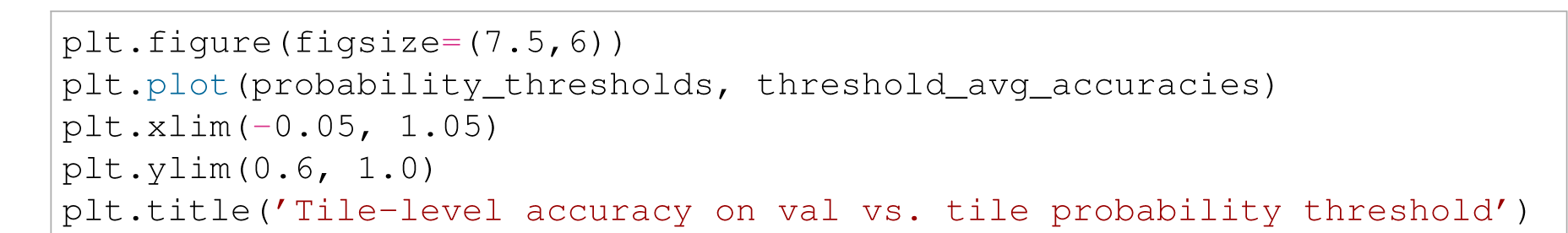
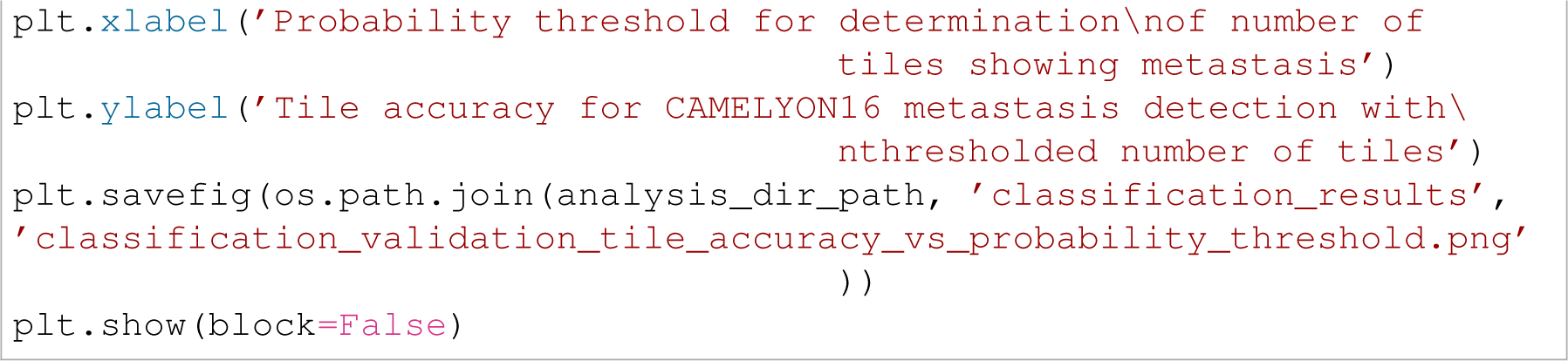
21. To compute the test set classification accuracy assuming that the test set has ground truth annotations (giving tile-level labels), iterate over the test set cases, loading the `Slide` object for each. Compute the accuracy of the classifier on the test set at the best probability threshold found from the validation set above by calling `Slide.classifierMetricAtThreshold()` on each slide, this time using only the best probability threshold as an argument, and keeping all other arguments the same as used on the validation set. Take the mean of the accuracies output by the `Slide.classifierMetricAtThreshold()` call of each test set `Slide` to evaluate the classifier’s holistic performance on the test set. 

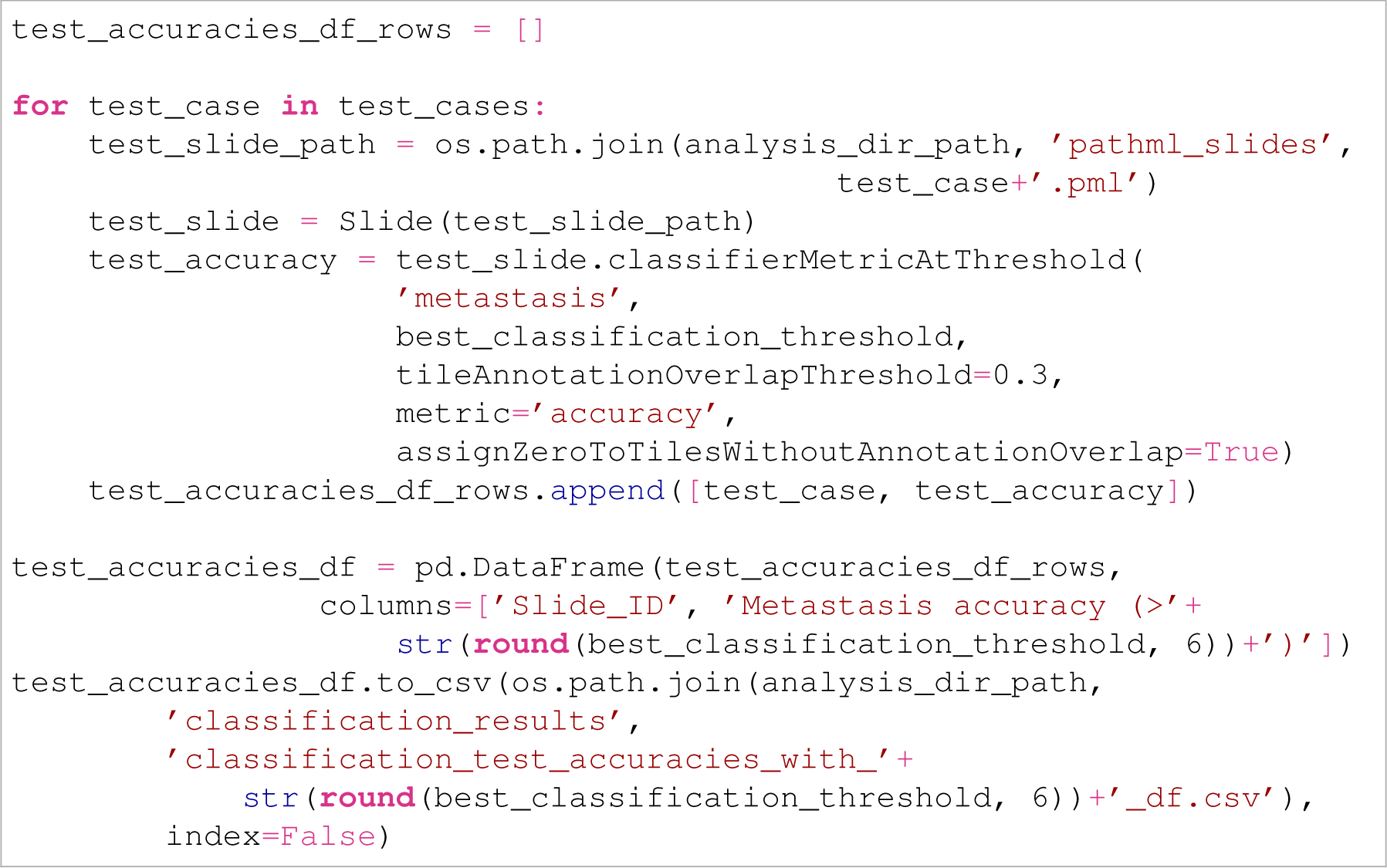
22. If there are no ground truth annotations on the test set, and only slide-wide ground truth labels, test set performance can be measured by counting the number of tiles the classifier found to be positive for the class of interest (using the best probability threshold found above to determine tiles the model considers positive) for all test set slides, and then comparing these counts to the slide-wide ground truth label for that class in an AUC analysis.

To count the number of tiles assigned a probability for the class of interest at or above the best validation probability, iterate over the test set cases, load each case’s `Slide` object, and call `Slide.numTilesAboveClassPredictionThreshold()` on it, using the class of interest as the first argument, and the best validation threshold as the second argument. Collect the positive-predicted tile counts for each test set `Slide`.

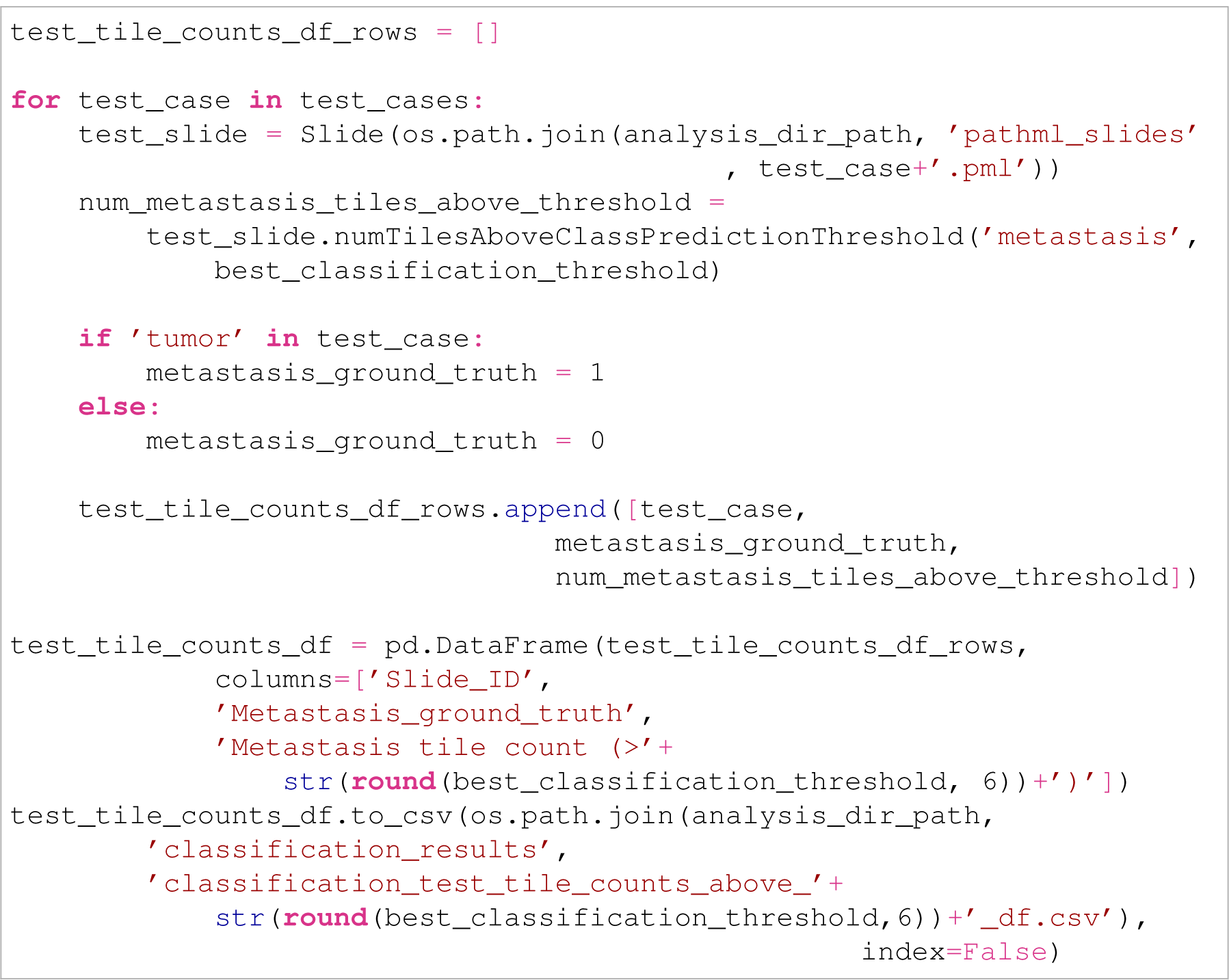

23. Compute the test set AUC, comparing the number of tiles that the classification model predicted to be from the class of interest with the slide-level ground truth. Plot the AUCROC curve.

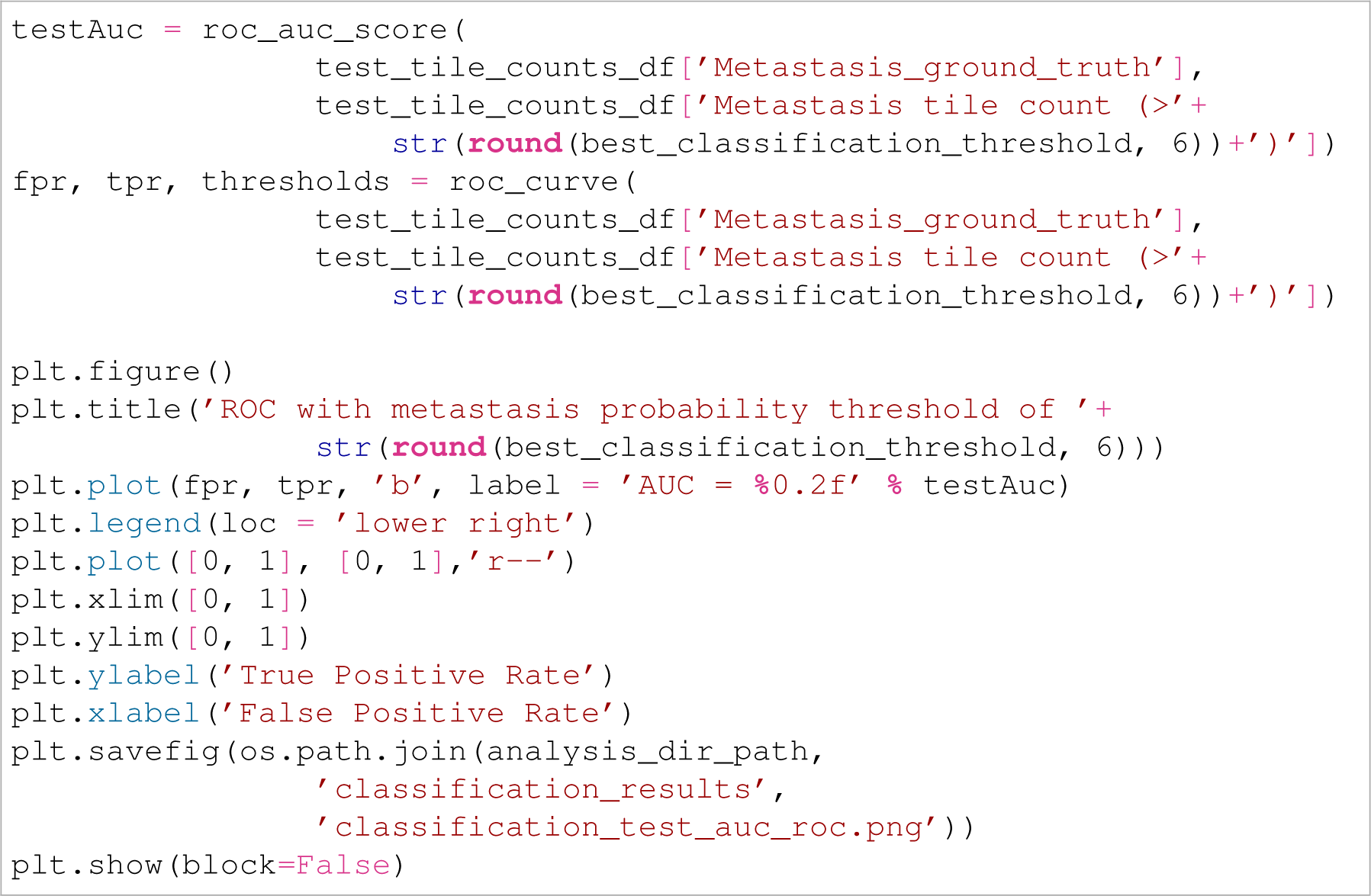

### Training a segmentation model

24. To train a segmentation model using using PathML fist append the appropriate segmentation model library to the path and import the necessary functions from it. This tutorial presents segmentation using a U-Net architecture (*40*), and uses the open source implementation of U-Net for PyTorch (including chunks of the code below) found at the following link: https://github.com/milesial/Pytorch-UNet. 

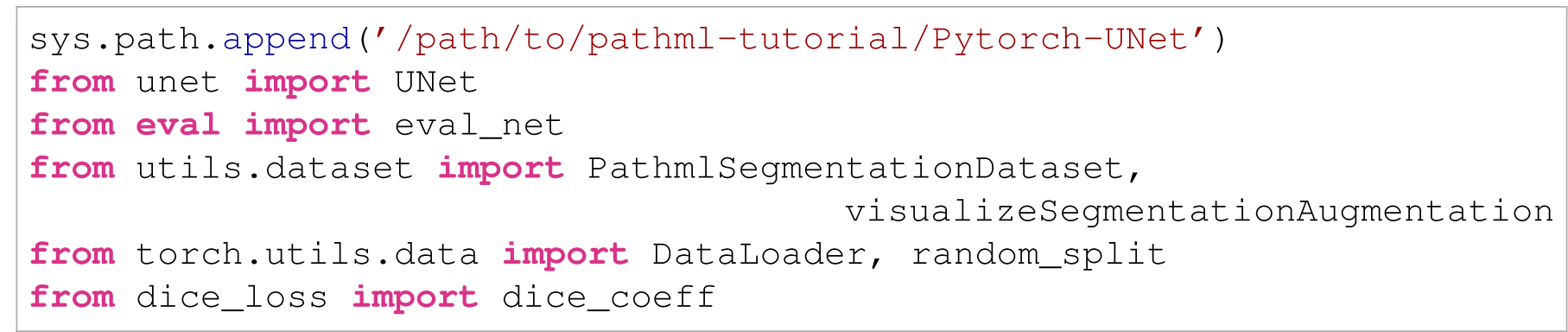
25. When training a segmentation model with PathML, it is first necessary to extract tiles and their corresponding binary masks into the directories `tiles` and `masks` using `Slide.extractAnnotationTiles()` and/or `Slide.extractRandomUnannotatedTiles()`. Since this was already performed above, begin by initialising a segmentation model architecture, moving it to the GPU, defining a loss function, optimiser, and learning rate scheduler (if desired), as well as training and validation data transforms for data augmentation. Use the Albumentations library to define the augmentations so that they can be performed in parallel between a tile image and its corresponding mask (*58*). Since `PathmlSegmentationDataset`, which will be used below, internally divides pixel values by 255, it isn’t necessary to include `ToTensorV2()` to do this at the end of the Albumentations augmentation composition. 

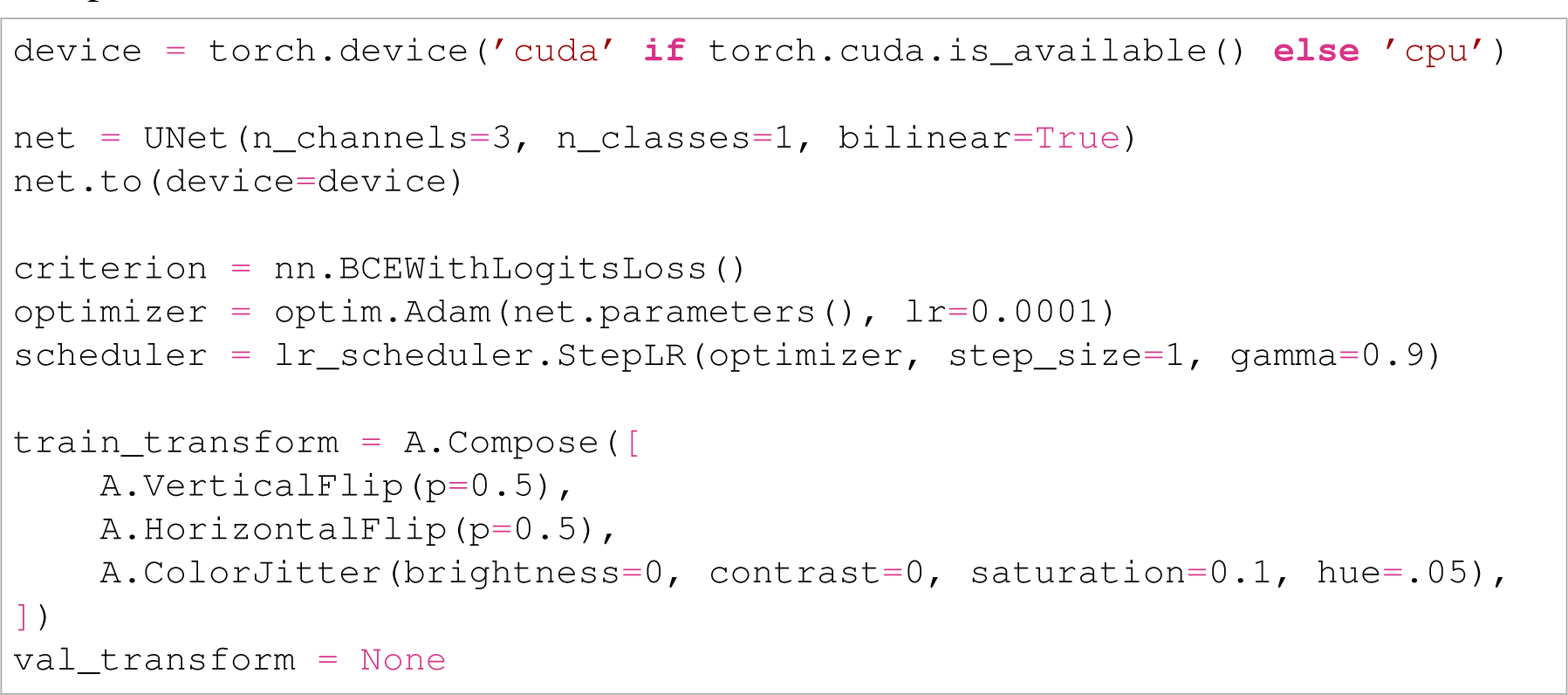
26. Visualise the augmentation on some example tiles and corresponding masks to confirm that the augmentations show the desired transformations, and that spatial transformations occur in parallel between the tile image and mask. The function `visualizeSegmentationAugmentation()` performs this. The code below can be re-run to show augmented examples of randomly selected tiles from the validation WSI `tumor_009`. Note that when visualising augmentations, it is important to avoid normalising the channel values to be between 0 and 1 so that they remain between 0 and 255 for the plotting function to show the colour appropriately. 

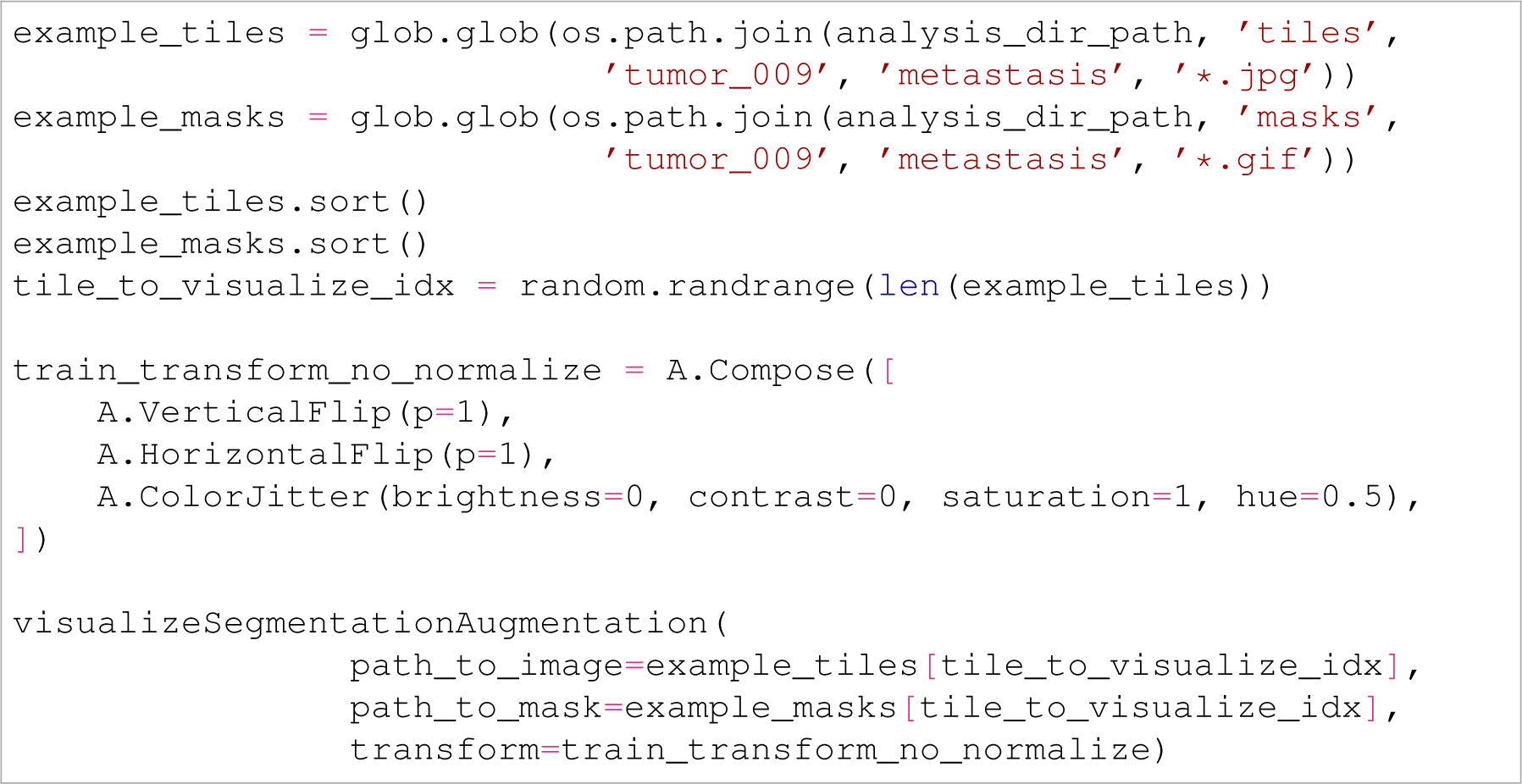
27. Define a function to train a segmentation model which takes as input a segmentation model architecture, the paths to our `tiles` and `masks` directories, as well as our loss function, optimiser, learning rate scheduler (if used), batch size, epoch count, and our data transform compositions for both training and validation. The function below constructs datasets using `PathmlSegmentationDataset`, a PyTorch-compliant dataset that works with PathML’s tile and mask output directory structures. The training function completes the desired number of epochs of training the model, checking validation Dice score performance at each epoch. It is trained from scratch (no transfer learning) and returns the learning statistics accumulated during training as well as the trained model. 

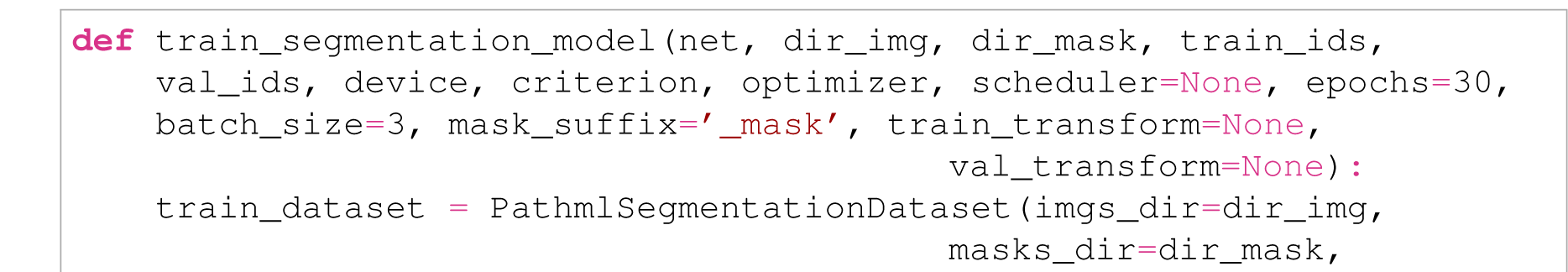
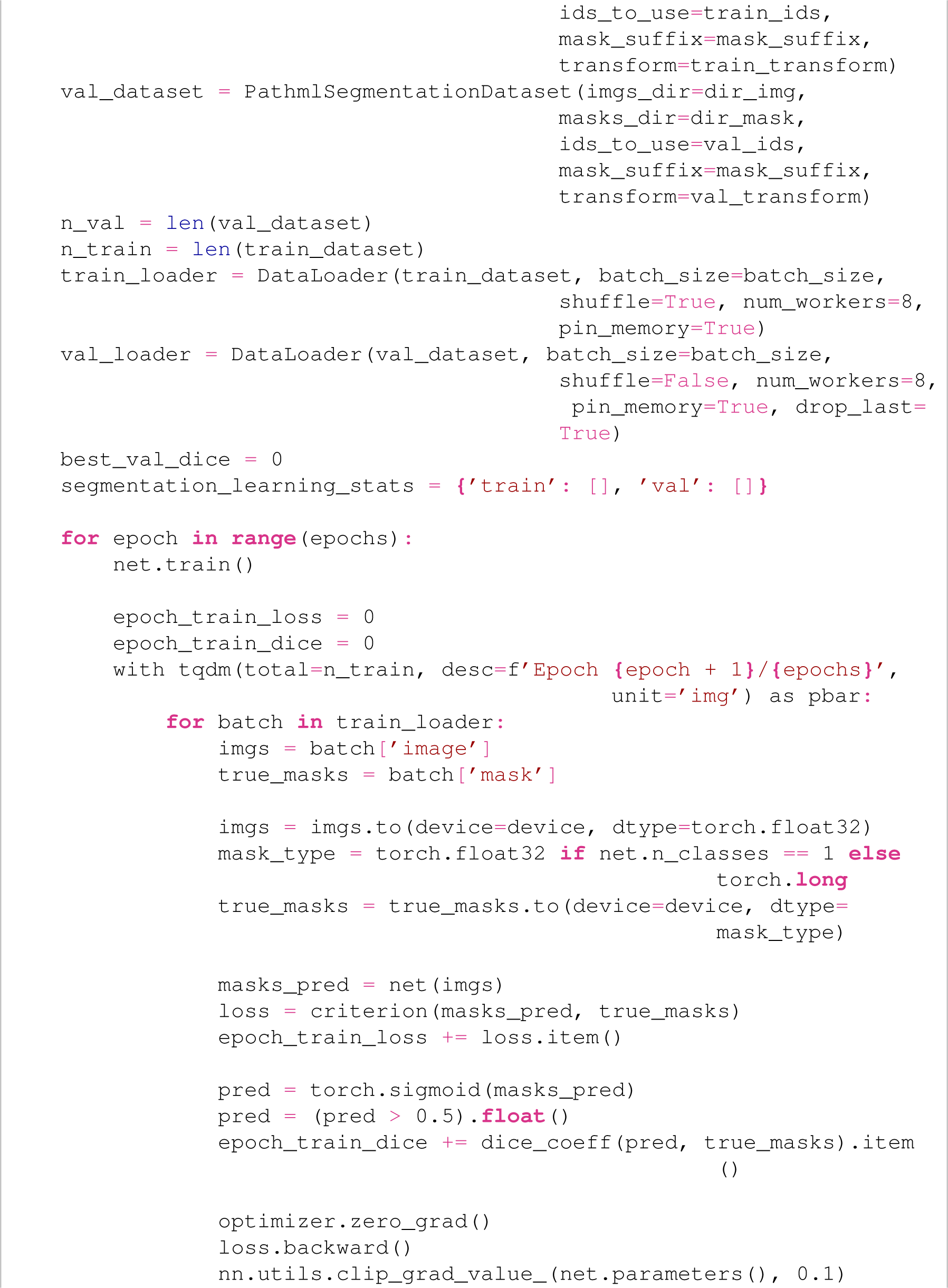
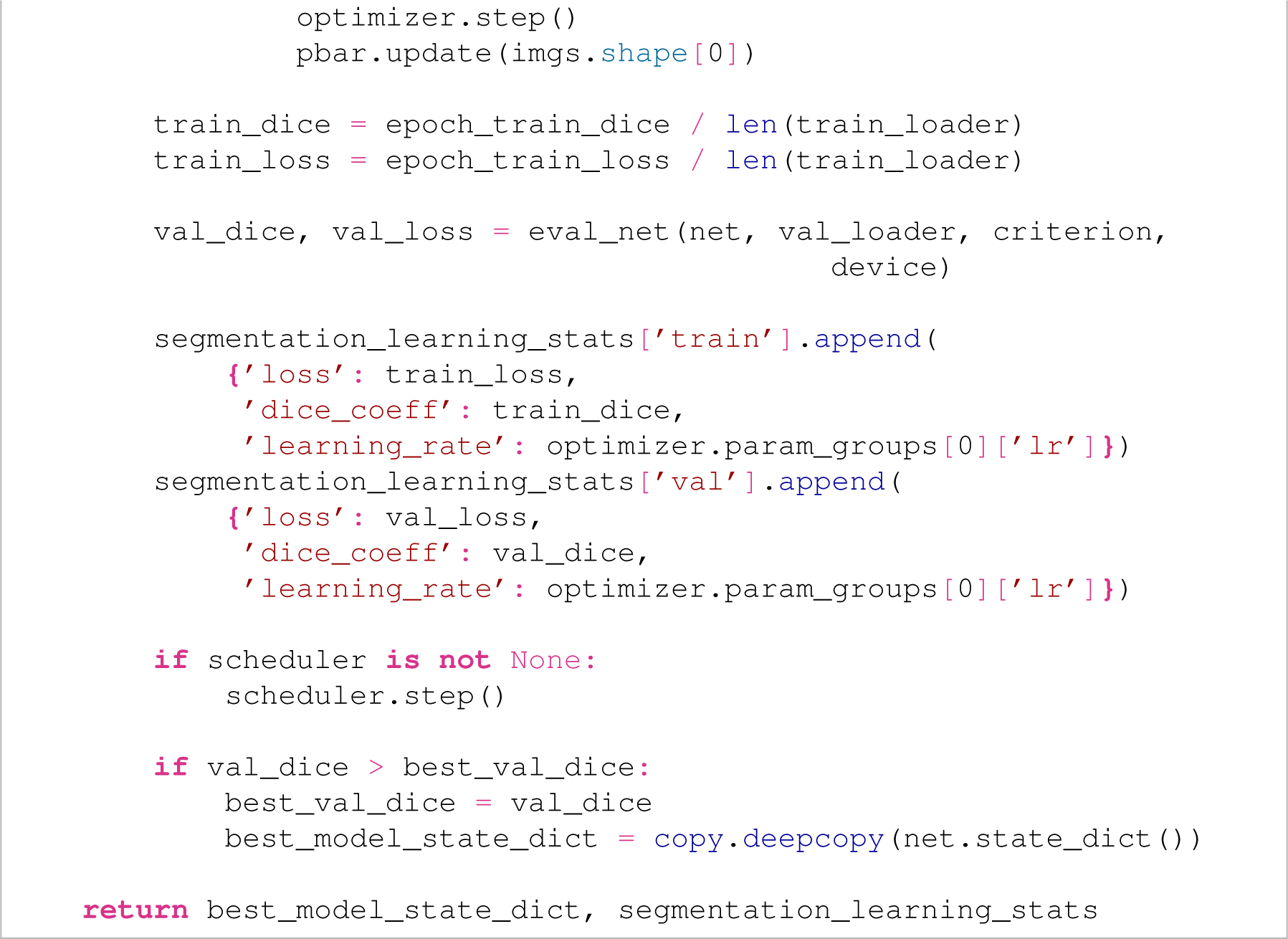
28. Run the segmentation training function using the desired hyperparameters defined above as well as the directory locations for the tiles, masks, and segmentation results (`tiles`, `masks`, and `segmentation_results`), and save the resulting trained model and accumulated learning statistics. As always, if your GPU runs out of memory, reduce the batch size until it can fit an entire batch. 

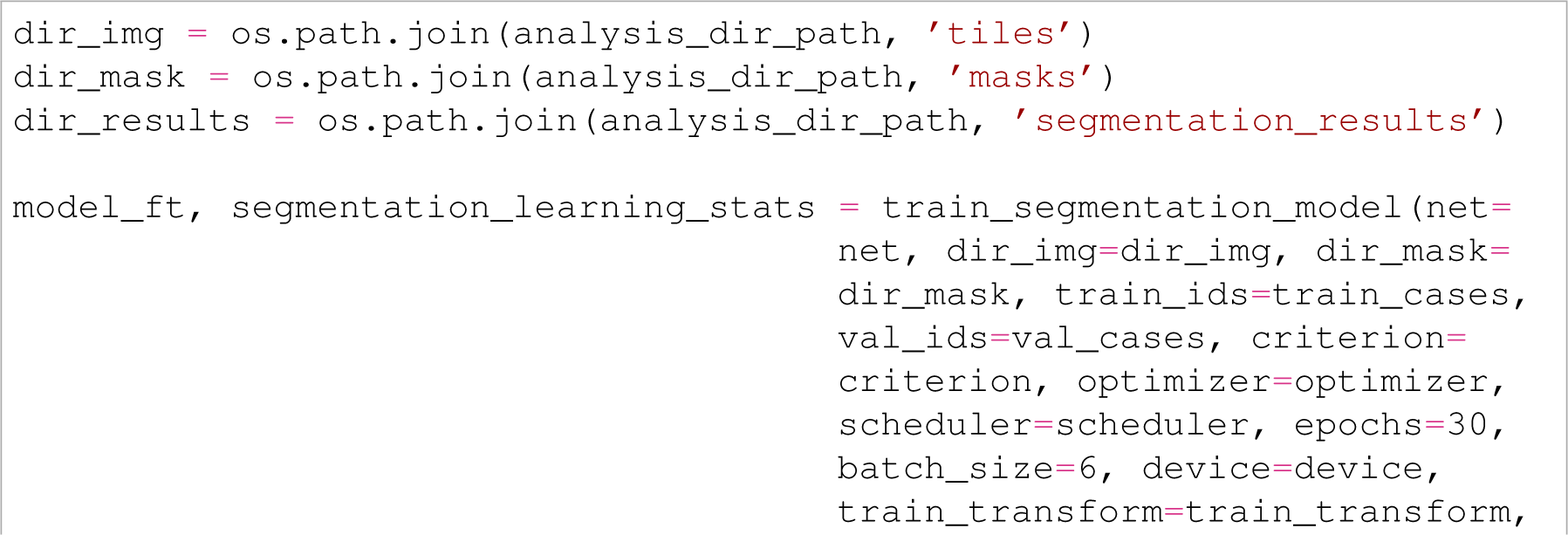
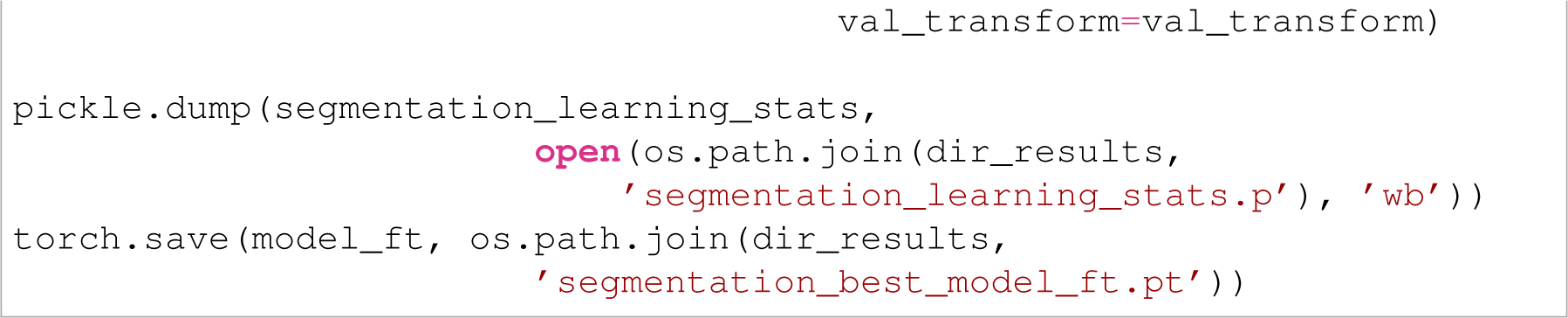
29. Plot the key learning statics, including at a minimum training and validation Dice coefficients and training and validation losses, from training to evaluate how the model performed and check for over- or under-fitting. 

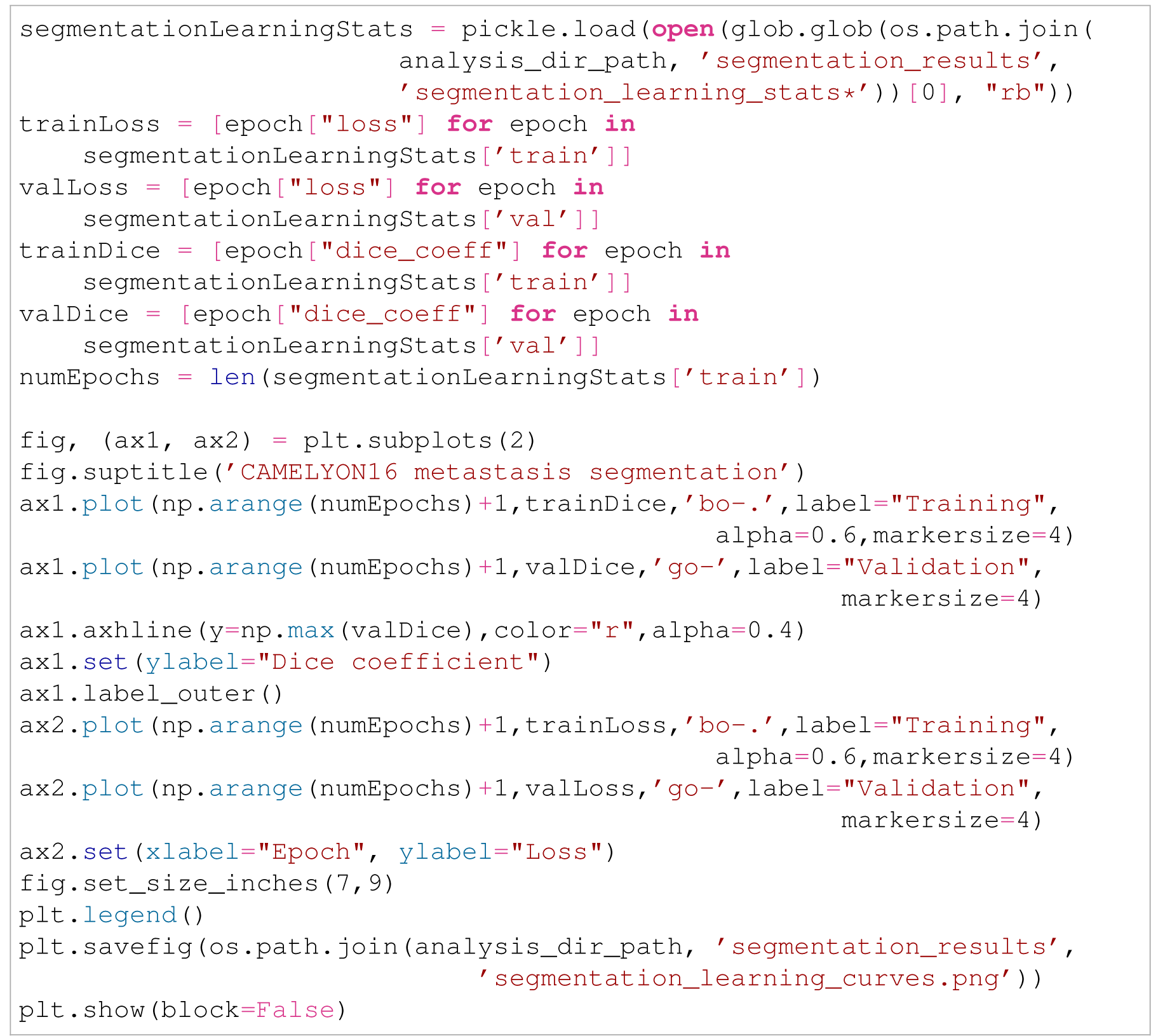

### Inferring on the trained segmentation model and validating model performance

30. To evaluate the model’s test set performance, begin by loading the trained model back into a model architecture of the same kind and then iterate over all training and validation `Slide` objects (loading them first). On each such `Slide`, run `Slide.inferSegmenter()` (the parallel function to `Slide.inferClassifier()`) to infer the trained model on the `Slide`. This will add a numpy matrix of model predictions the dimension of the input tiles to each suitable tile in the `Slide` object’s tile dictionary. The first argument to `Slide.inferSegmenter()` is the trained model which was loaded, the `classNames` argument is a list comprising the classes in the trained model to add an inference matrix to the `Slide` object’s tile dictionary for, the the `foregroundLevelThreshold` and `tissueLevelThreshold` arguments are used in the same way as in `Slide.inferClassifier()` (to only perform inference on tiles that are considered tissue), and the `batchSize` argument specifies the size of the inference batches. Call `Slide.save()` to preserve these inferences in the `Slide` objects.

Be aware that adding inference matrices to the tile dictionary can make the `Slide` objects quite large (often 1–10 gigabytes), so be wary as to where the `Slide` objects are saved and whether there is memory for them once they’ve been saved after calling `Slide.inferSegmenter()`. It is highly recommended that unless extreme precision in inference predictions is required, users set the `dtype` argument of `Slide.inferClassifier()` to ”int” (this is also the default). This will store inference predictions in `numpy.uint8` format (scaled to be integers in the range 0–255) rather than a much larger float format. Downstream functions in PathML will automatically detect the presence of these 0–255 classification predictions (as opposed to 0–1 float predictions) and treat them appropriately.

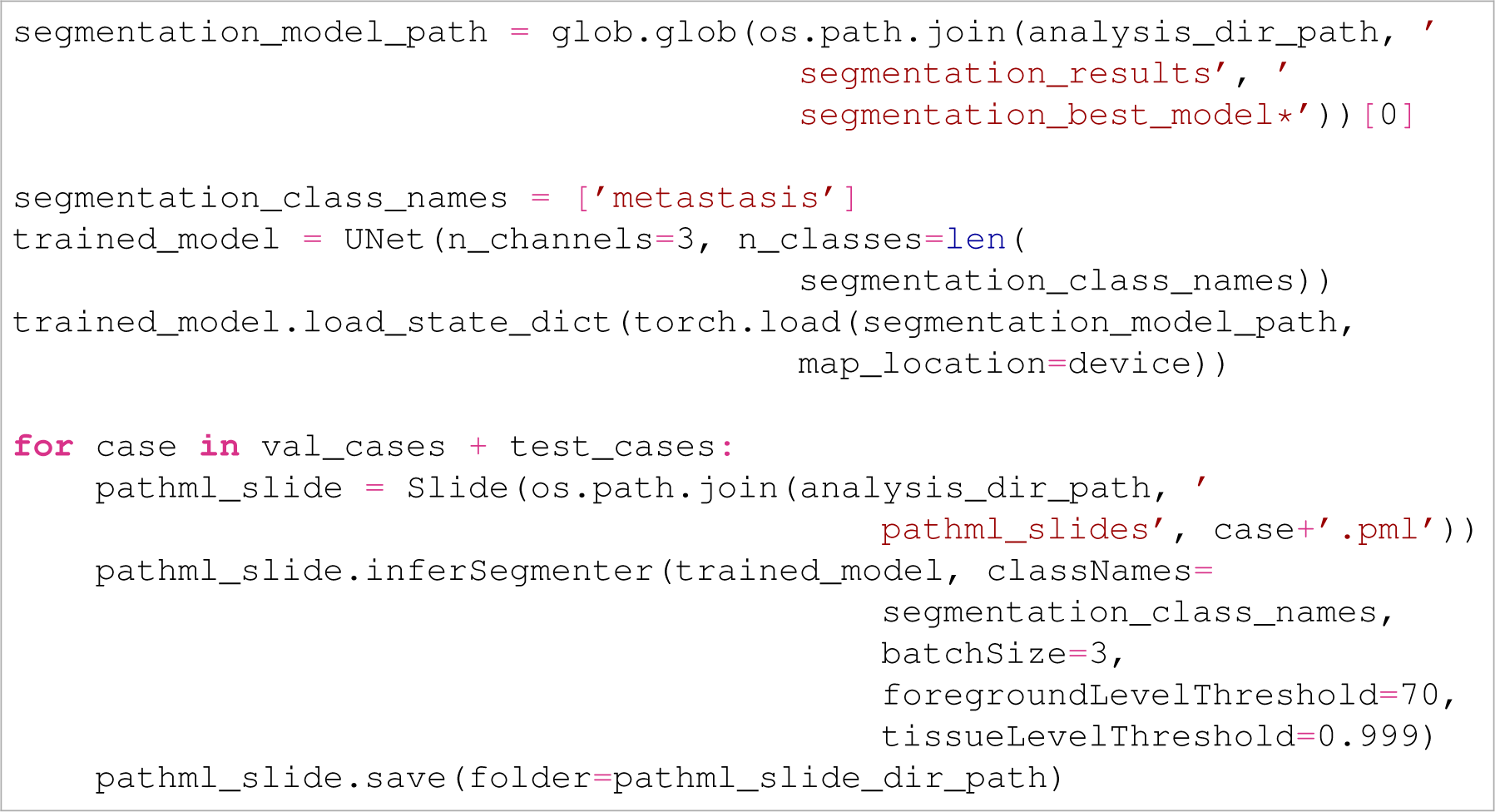

31. Visualise the pixel-wise inference map at the inferred-on tiles of each validation `Slide` using `Slide.visualizeSegmenterInference()`, the segmentation parallel of `Slide.visualizeClassifierInference()` that makes a figure showing the inference map atop the WSI. Set the first argument of the function to be the name of the class to check the inference map for. Use this to evaluate whether the trained model has learned the right thing and correctly identifies regions of the class of interest. 

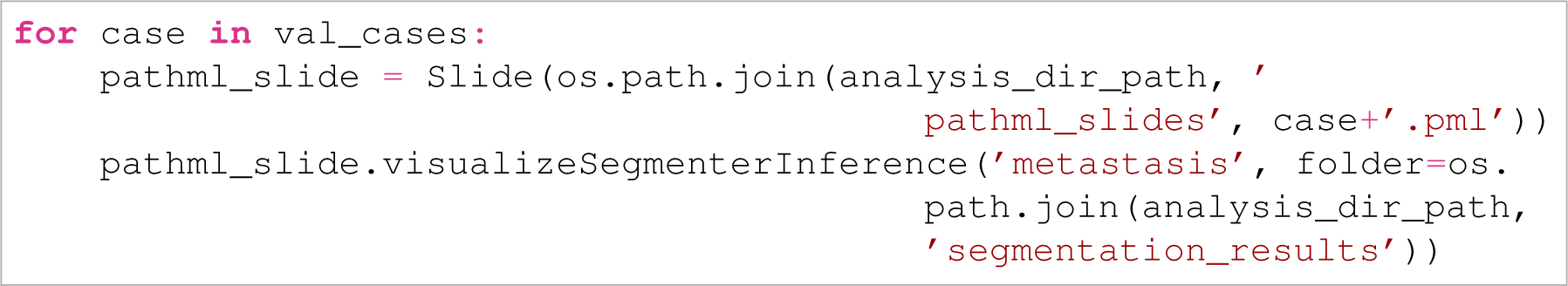
32. As with the classification task, to evaluate test set performance, it is first necessary to find a probability threshold to use to binarise the output probabilities (the predictions) of the trained model using the validation set. Just as `Slide.classifierMetricAtThreshold()` was used above to check accuracy across many potential probability thresholds to select the best one, use `Slide.segmenterMetricAtThreshold()` to select the best probability threshold from a list to produce the best Dice coefficient, a measure of accuracy for segmentation outputs. Iterate over all validation `Slide` objects, running `Slide.segmenterMetricAtThreshold()` on each. The first argument is the class of interest, the second is a list of probability thresholds to try, the set the `metric` argument is `dice_coeff`. This will return a list parallel to the probability thresholds list of average Dice coefficients comparing the inference prediction matrices of all tiles added to the `Slide` by `Slide.inferClassifier()` against the ground truth matrices added from `Slide.addAnnotations()`, where pixels in the prediction matrix at or above the given probability threshold are considered to have been marked as positive for the given class by the trained model.

Be aware that computing the Dice coefficient for thousands of tiles is a time-intensive step. Reducing the number of probability thresholds that are checked will substantially reduce the time required. It takes about one hour on average to check the 41 thresholds below on one `Slide`. As before, we have elected to check probability thresholds at greater frequency the closer we get to probability 1, as we found that for this problem, the best probability threshold for segmentation lies close to 1, as we would like the most accurate best threshold we can achieve. 

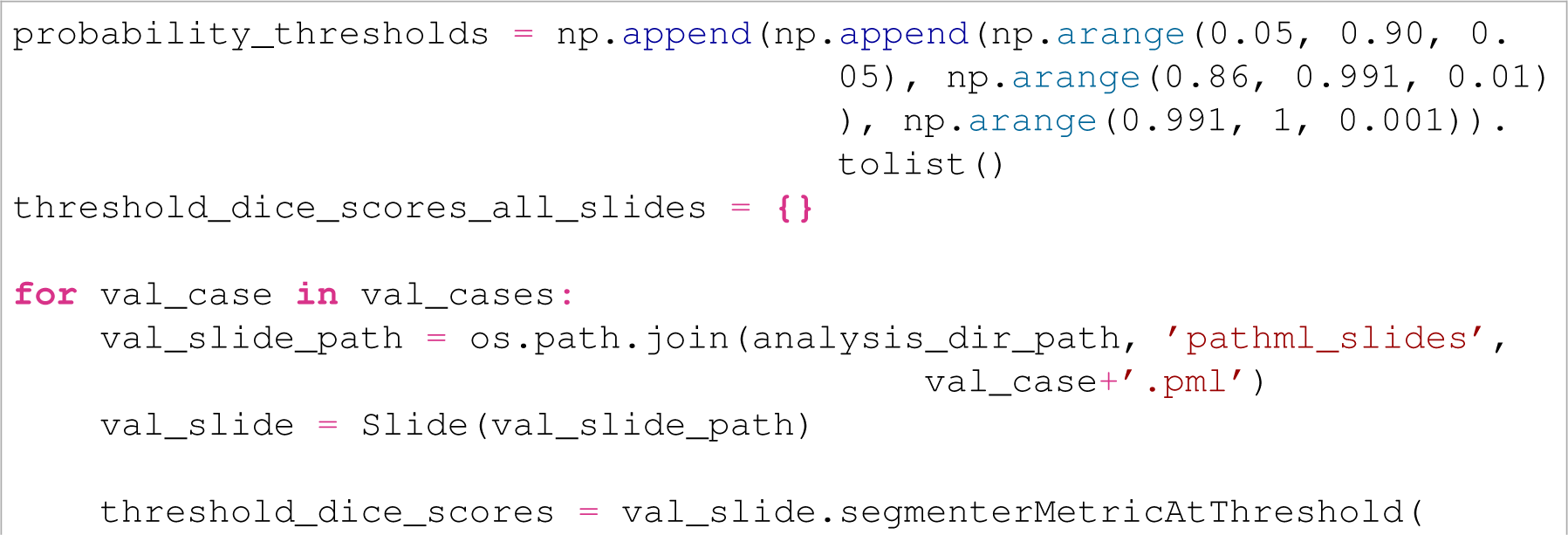
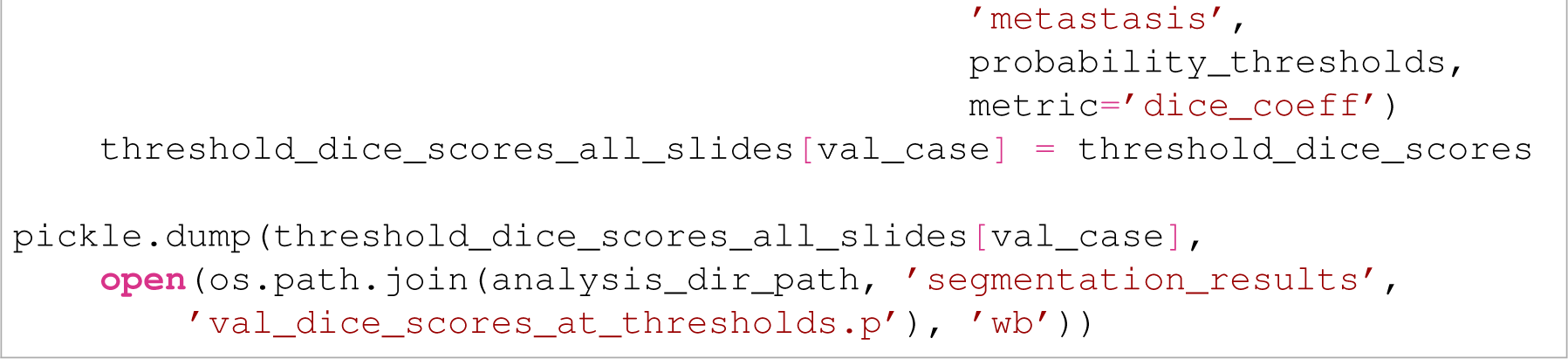

33. Take the element-wise mean of the Dice coefficient lists output by `Slide.segmenterMetricAtThreshold()` across all validation slides, then find the probability threshold which produced the maximum Dice coefficient in this mean list. This is the best validation probability threshold that will be used to check the trained model’s performance on the test set. 

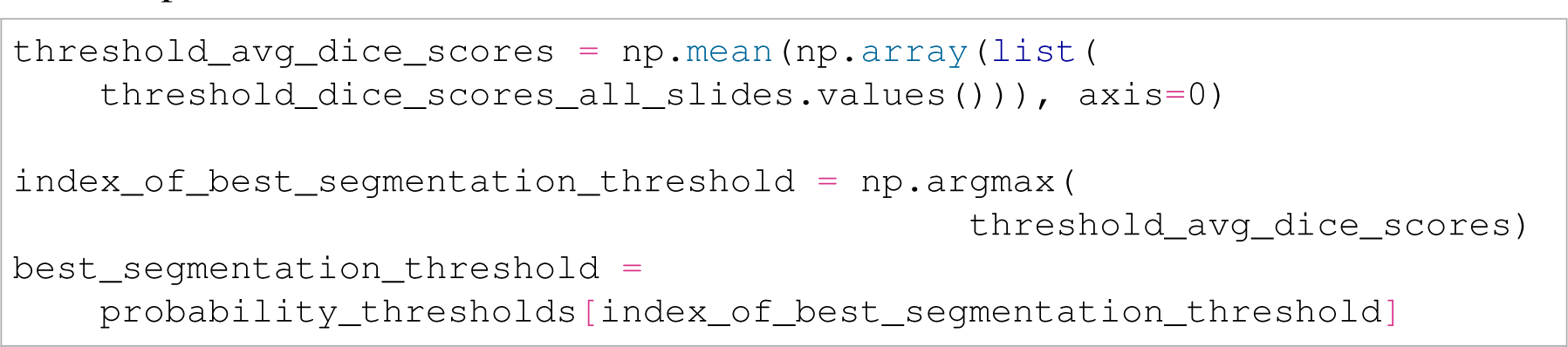
34. Plot the element-wise average prediction Dice coefficients on the validation set against the probability thresholds used to generate them to visualise how changing the probability threshold changes validation Dice coefficients. 

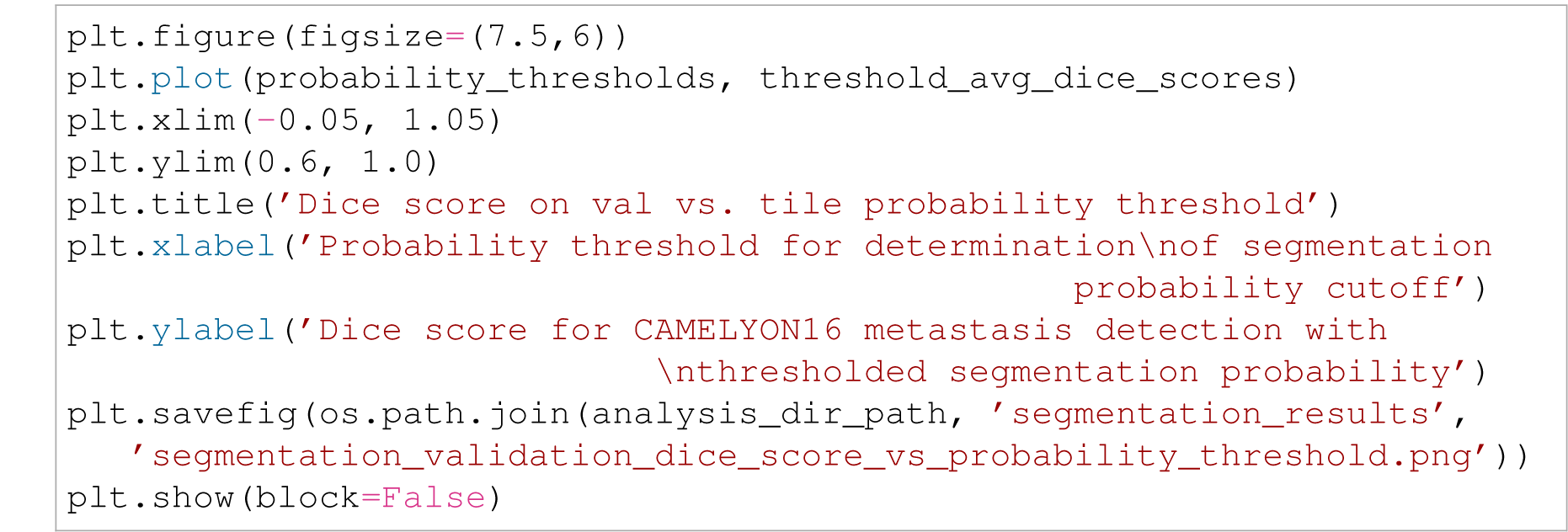
35. To compute the test set Dice coefficient, iterate over the test set cases, loading the `Slide` object for each. Compute the Dice score of the trained segmentation model on the test set at the best probability threshold found from the validation set above by calling `Slide.segmenterMetricAtThreshold()` on each slide, this time using only the best probability threshold as an argument, and keeping all other arguments the same as used on the validation set. Take the mean of the Dice scores output by the `Slide.classifierMetricAtThreshold()` call of each test set `Slide` to evaluate the classifier’s holistic performance on the test set. 

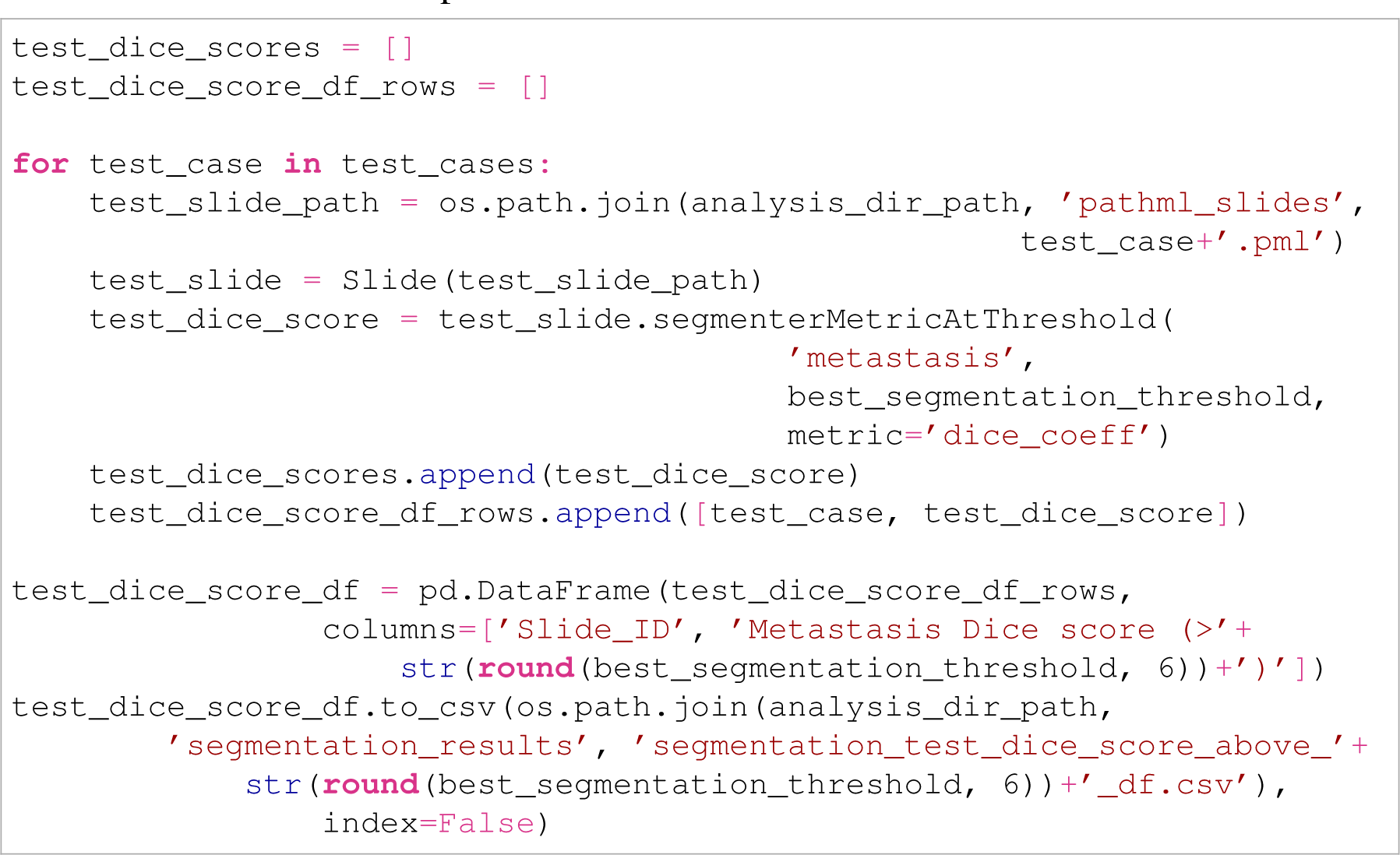
4. 36. Export the full inference map for the class of interest for each test set `Slide` by iterating over all test set `Slide` objects and calling `Slide.getNonOverlappingSegmentationInferenceArray()` on them, using as argument the name of the class to extract the inference arrays for. This will, for each `Slide`, return a 2D numpy matrix with the pixel dimensions of the WSI used to make the `Slide` object, where each pixel contains the 0 to 1 probability prediction of the trained model on that pixel if it was inferred on during `Slide.inferSegmenter()`.

If tile overlap is used, then the average prediction for a pixel across all pixel predictions that overlap it will be provided. If a binary rather than a float prediction mask is desired, include `probabilityThreshold` as an argument and set its value to the probability threshold at or above which a pixel should be considered positive for the given class. If it is preferred that pixels that were not inferred on be given values of zero rather than `numpy.nan` values, set the argument `fillUninferredPixelsWithZeros` to `True`.

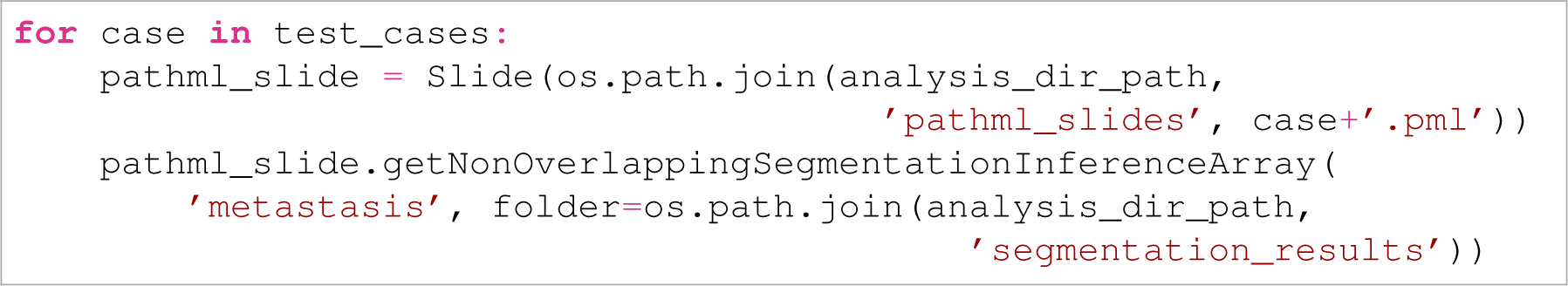

## 3 Troubleshooting

There are several mistakes and error messages that can arise when using PathML. **Table 2** presents the most common mistakes users might run into, including the **Procedure** step it is likely to occur in, the error message output by PathML, the possible reason for the mistake, and the possible solution to it.

**Table 2:** Common PathML error messages with explanations and possible solutions. X, *Y*, and *Z* represent numbers or words that will vary depending on the exact error made by the user.

| Step | Message | Possible reason | Solution |
| --- | --- | --- | --- |
| 5 | 'This image is not compatible. Please refer to the documentation for proper installation of openslide and libvips' | The WSI input into the PathML initialiser was not a supported file format | Convert the WSI to a file format supported by lipvips. |
| 5 | 'Tissue detection has already been performed. Use overwriteExistingTissueDetection if you wish to write over it' | Slide.detectTissue() has already been called on the Slide object | Set the overwriteExistingTissueDetection argument to True, or else don't perform tissue detection again |
| 5 | 'Annotation with centroid (X, Y) produces a Shapely Z instead of a polygon; check to see if it self-intersects.' | The annotation around the specified centroid pixel coordinates of the WSI does not produce a polygon when geometrically parsed | Check that annotation on the WSI in a WSI viewer looking for self-overlapping regions and correct it to be a polygon |
| 7 | 'Warning: X suitable Y tiles found but requested Z tiles to extract. Extracting all suitable tiles...' | The numTilesToExtractPerClass argument of a tile extraction function exceeds the number of suitable tiles of class Y | Reduce the numTilesToExtractPerClass argument for class Y, or else let PathML will extract all available Y tiles by default |
| 17, 30 | 'Model has X classes but only Y class names were provided in the classes argument' | The number of classes output by the model inputted to Slide.inferClassifier() or Slide.inferSegmenter() does not equal the number of classes present in the classNames argument | Verify that classNames includes all the classes that were trained on, and correct this argument as necessary |
| 17, 30 | 'No predictions found in slide. Use inferClassifier() / inferSegmenter() to generate them.' | Using a PathML function reliant on inference results without having added inference results to the Slide object | Run Slide.inferClassifier or Slide.inferSegmenter() on the Slide object |

## 4 Timing

The time required to run a pipeline with PathML can vary incredibly depending on many factors, including the number of WSIs used, the number of annotations in each annotation file, the number of tiles requested, and the computational power available, and the power of the GPU (if using the inference functions). For reference, here are the run times of the more timeconsumptive steps of the **Procedure** using a 4-core Intel Xeon E5-2623 v3 CPU and an Nvidia GeForce RTX 2080 Ti GPU (running on Ubuntu 18.04):

**Procedure steps 1–6**: Detecting tissue, detecting foreground, and adding annotations to `Slide` objects takes 36 minutes for the 18 CAMELYON16 WSIs.
**Procedure steps 7–9**: Extracting 500 tiles per WSI takes 7 minutes.
**Procedure steps 10–16**: Training a VGG19 classification model with batch normalisation ( (*59*)) for 30 epochs on 3,000 training set tiles and 3,000 set validation tiles (checking validation performance after every epoch) takes 34 minutes.
**Procedure steps 17–23**: Inferring the trained VGG19 classification model on 6,000 validation and test set tiles takes 13 minutes. Visualising the six validation inference maps takes 8 minutes. Finding the best probability threshold using the validation set takes 1 minute.
**Procedure steps 24–29**: Training a U-Net segmentation model on the same 3,000 training set tiles and 3,000 validation set tiles (again checking validation performance at every epoch) for 30 epochs takes 3 hours and 6 minutes.
**Procedure steps 30–31**: Inferring 6,000 validation and test set tiles on the trained U-Net architecture takes 51 minutes. Visualising the inference maps of these same 6 validation WSIs takes 7 minutes.
**Procedure steps 32–35**: Computing the Dice coefficients of the 6 validation set `Slide` objects across 41 probability thresholds takes 6 hours and 23 minutes, and then applying the best threshold to get the Dice coefficients of the 6 test set `Slide` objects takes 21 minutes.
**Procedure step 36**: Stitching together and exporting the complete pixel-wise segmentation inference masks for 6 test set `Slide` objects takes 39 minutes.

## 5 Anticipated results

### 5.1 Assessing foreground and deep tissue detections

If using the pipeline presented in the **Procedure**, Step 6 shows how to filter out non-tissue tiles from `Slide` objects with `Slide.detectTissue()` and/or `Slide.detectForeground()`. To evaluate which parameters to use when filtering (e.g. what to set the `tissueLevelThreshold` and `foregroundLevelThreshold` arguments to), `Slide.visualizeTissueDetection()` and `Slide.visualizeForegroundDetection()` are made-for-purpose. Standard practice is to decide on a range of filtering thresholds to test, perform a grid search, calling these visualisation functions at each combination of filtering parameters, comparing to a thumbnail image of the WSI provided by `Slide.visualizeThumbnail()`. See **fig. 4** for examples of the results of various filtering methods. **Figure 4B** shows the filtering resulting from Otsu’s method. It provides a stringent filter that doesn’t include any non-tissue regions in the foreground areas, but it also excludes lighter-coloured tissue in the centre of the top tissue region. **Figure 4C** shows the triangle algorithm, which is conversely too permissive, including dark artefact regions around the edges of the slide as foreground. **Figure 4D** shows the results of PathML’s built in deep tissue detector, which unlike the simple foreground filtering methods includes an artefact class. In the **Procedure step 6** example presented in **fig. 4** (one of the validation WSIs), the deep tissue detector is by far the most performant, neither under nor overcalling tissue while identifying artefact regions as such. The filtering parameter or combination of parameters that provide the best results upon examination can then be used for all the `Slide` objects.

### 5.2 Assessing extracted tiles, masks, and data augmentation choices

Steps 7–8 of the **Procedure** shows how to use PathML to extract tiles and segmentation masks from `Slide` objects with `Slide.extractAnnotatedTiles()` and `Slide.extractRandomUnannotatedTiles()`. The first step of assessment after running these functions is to make sure the channel-wise mean and standard deviation results computed from the data returned by these functions (see Step 8) are in the expected ranges. For reference, the channel means of ImageNet are 0.485, 0.456, 0.406 for the red, green, and blue channels respectively, and ImageNet’s standard deviations are likewise 0.229, 0.224, and 0.225 (*42*). As might be expected, H&E-stained WSIs tend to to have higher red and blue channel values compared to the average of ImageNet, which is a set of macroscopic images including animals, buildings, natural landscapes, and more. In **table 3** it is apparent that this is reflected in the CAMELYON16 H&E slides used in the **Procedure**. The standard deviations, also as expected, are roughly similar to those of ImageNet. Since these means and standard deviations will be used to normalise the tiles going into the model, if any values break from intuitive expectation, check the tiles visually to ensure that a representative sampling of tissue tiles has been achieved.

**Table 3:** Channel means and standard deviations across tile dataset.

| Channel | Mean | Standard deviation |
| --- | --- | --- |
| Red | 0.711 | 0.219 |
| Green | 0.537 | 0.256 |
| Blue | 0.674 | 0.195 |

**Procedure steps 10–12** cover defining data augmentations for classification. To assess whether a set of augmentations are satisfactory, it is necessary to visualise examples of tiles that have gone through the augmentation to ensure that they are as expected. As CAMELYON16 slides were scanned at different institutions (*57*), there is a range of stain hue and brightness that needs to be augmented for. **Figure 5** shows a batch of tiles that have undergone the colour jittering and random horizontal and vertical flip augmentations of Step 10. It is apparent that strong colour jitter, particularly hue and contrast, have been used, and the tiles otherwise look as expected.

**Procedure steps 25–26** define augmentation parameters for segmentation. When reviewing segmentation augmentation, it is essentially to visualise the segmentation mask along with the tile to ensure that the spatial transformations and cropping were applied to both the tile and the mask uniformly, and that the colour transformations were applied only to the tile image. **Figure 6** shows that the **Procedure step 12** example was augmented correctly.

Whether augmenting for a classification or segmentation task, if the transformations once visualised appear too light, too extreme, or unexpected, it is essential that the user adjust the augmentation parameters until the result is correct.

### 5.3 Assessing training statistics

**Procedure steps 14–16** demonstrate training a classification model from the tiles extracted by PathML, and **Procedure steps 27–29** show the same steps for a segmentation model. To assess the efficacy of training, it is critical to plot some training statistics to confirm that both training and validation loss generally decreased across epochs and that training and validation performance (accuracy for classification, Dice coefficient for segmentation) increased as epochs progressed. **Figure 7** shows that for the 6 training and 6 validation WSIs used in the **Procedure**, both of these trends hold true for both the training and validation sets.

**Figure 7:**
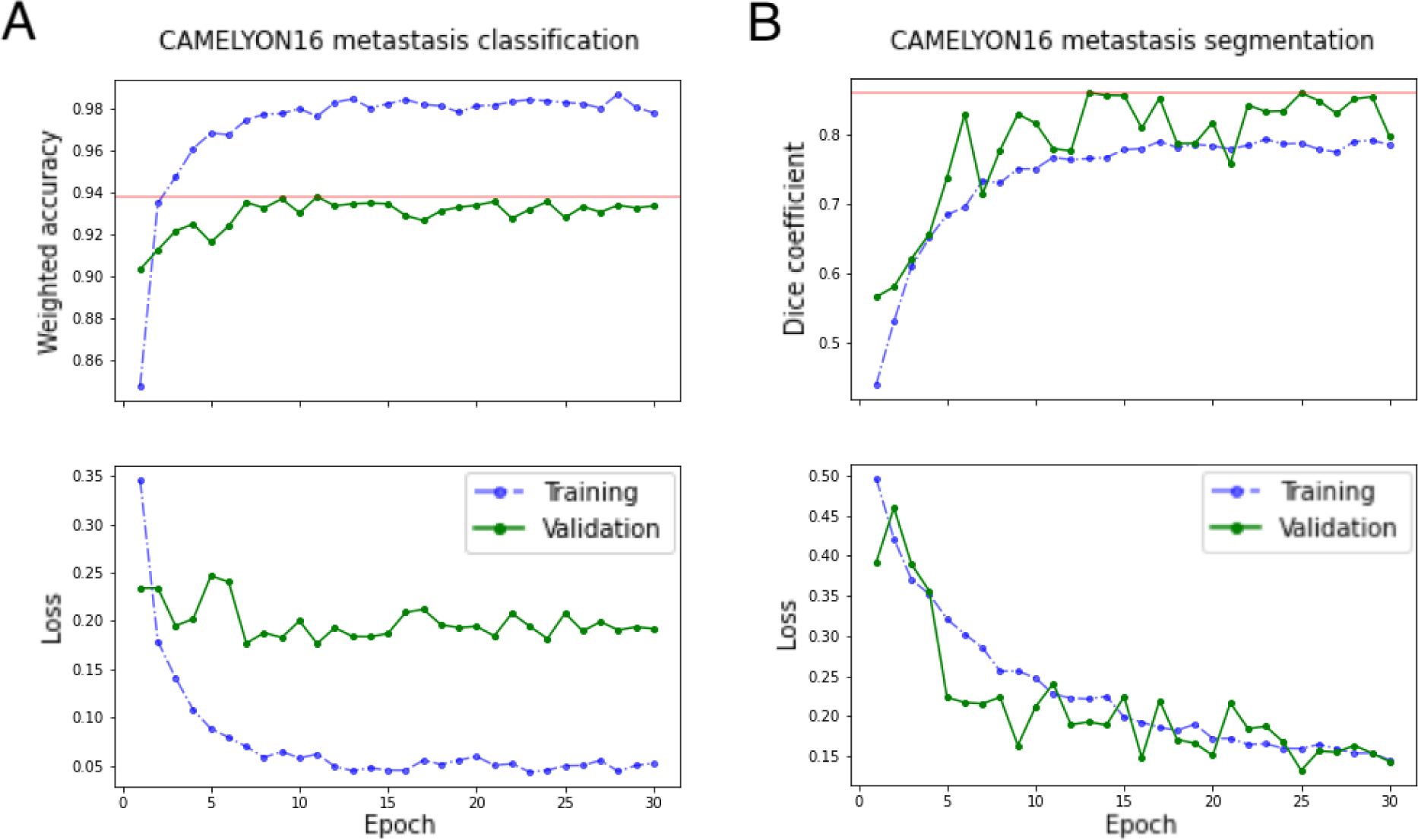
Training and validation learning statistics. **(A)** Training and validation accuracy and loss curves from a 30 epoch classification training run. **(B)** Training and validation Dice coefficient and loss curves from a 30 epoch segmentation training run.

If training performance increases while validation performance stagnates or decreases (or likewise if training loss decreases while validation loss remains flat or grows), this is an indicator that the model has overfit to the training set (**table 1**). To rectify this, a number of techniques can be employed, including increasing the size of the training dataset, using more aggressive data augmentation, trying a shallower architecture, using early stopping (if the model begins overfitting after a certain number of epochs) or other regularisation techniques (*12*). On the other hand, if even training performance remains flat or decreases across epochs (or training loss remains flat or increases), this indicates model underfitting. Underfitting could have a number of causes, but some common troubleshooting techniques include trying different hyperparameters, especially a wide range of learning rates, as well as reducing data augmentation, and increasing batch size to the largest permissible on the user’s GPU.

### 5.4 Assessing inference results

**Procedure steps 17 and 30** show the use of PathML’s `Slide.inferClassifier()` and `Slide.inferSegmenter()` functions for inferring the trained classification and segmentation models on the validation and test sets. An important step after inference is visually confirming, even if the performance statistic is adequate, what the model learned. `Slide.inferClassifier()` and `Slide.inferSegmenter()` plot the inference results of all tiles in the same spatial orientation as they occur on the WSI to form inference maps. **Figure 8** shows the inference maps of both the classification and segmentation models of the **Procedure** for the metastasis class of the three metastasis-positive validation WSIs. To assess them, evaluate whether the high metastasis probability regions align with the ground truth metastasis regions. Also check the metastasis-negative slides to ensure that few to no tiles have high metastasis probability.

**Figure 8:**
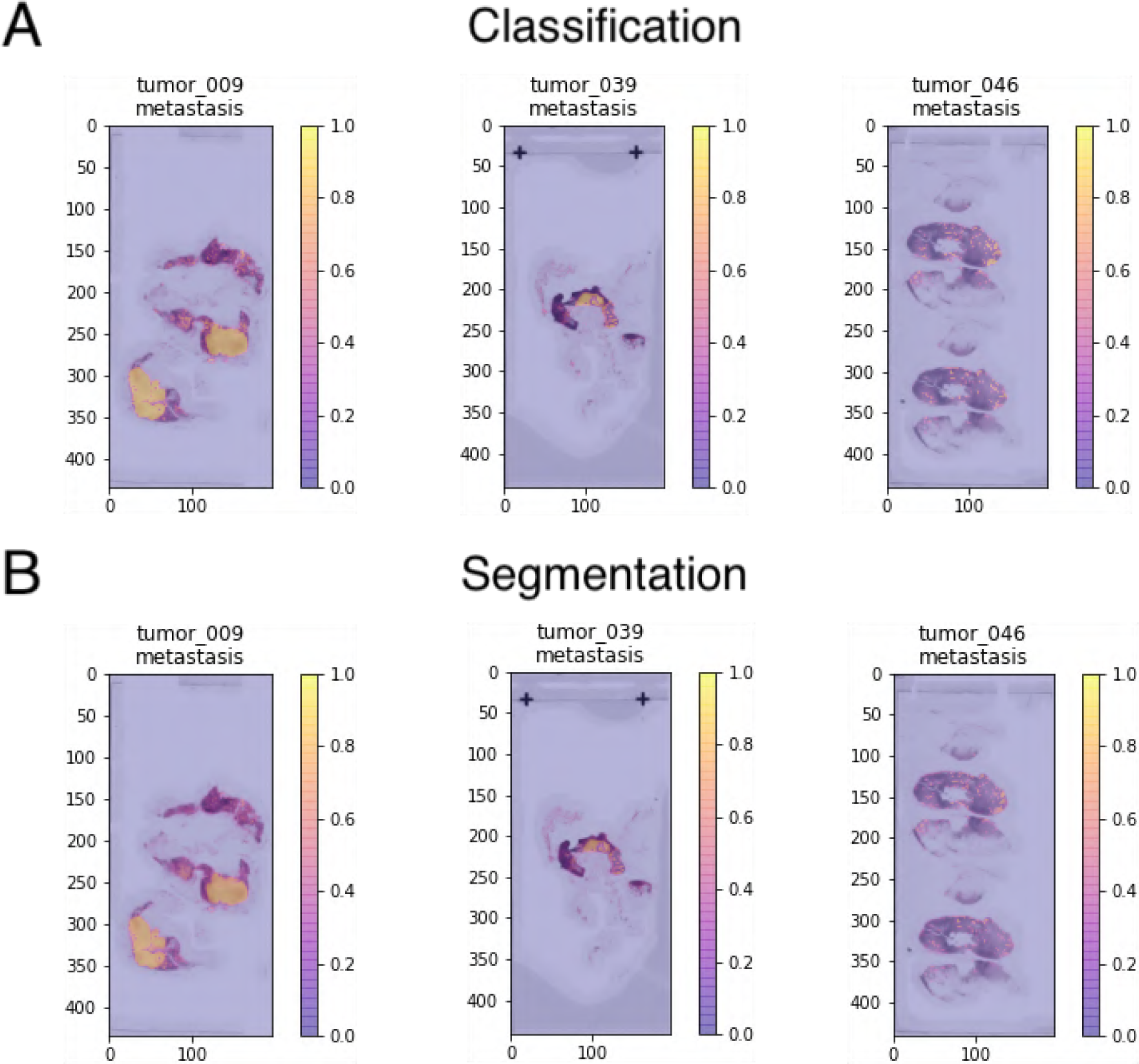
Visualising the inference of trained models on validation set. **(A)** A plot of the inference of the trained classification model on three validation slides, showing the metastasis class results. Plots were generated with Slide.visualizeClassifierInference(). **(B)** A plot of the inference of the trained segmentation model on three validation slides, showing the metastasis class results. Plots were generated with Slide.visualizeClassifierInference().

If any regions are wrongly called a certain class, it is recommended that the user zoom in on these regions in a WSI viewer such as QuPath or ASAP to determine if there is a pattern to the miscalls (i.e. a certain tissue appearance or artefact is consistently wrongly called) (*22, 23*). Users can then annotate and introduce examples like these wrongly called regions into the training set and retrain to teach the model the correct labels for these regions.

### 5.5 Assessing the best probability threshold to apply to the test set

After inference, it is often important to select an operating threshold from the validation set to apply to the test set. Plotting and selecting this threshold is performed in **Procedure steps 19–20** for the classification model and **Procedure steps 32–34** for the segmentation model using `Slide.classifierMetricAtThreshold()` and `Slide.segmenterMetricAtThreshold()`, respectively. These functions return the performance of the model on a desired metric across a range of probability thresholds. **Figure 9** shows plots of these performance metrics, accuracy for the classification model and the Dice coefficient for the segmentation model, across the range of tested inference probability thresholds on the validation set. Assessing plots of this nature can be tricky, as there is no standard pattern by which performance should behave as the probability threshold is altered. A smooth curve is expected. As seen in **fig. 9**, for both classification and segmentation of metastases in the CAMELYON16 subset used in the **Procedure**, very high probability thresholds yield the best performance on the validation set.

**Figure 9:**
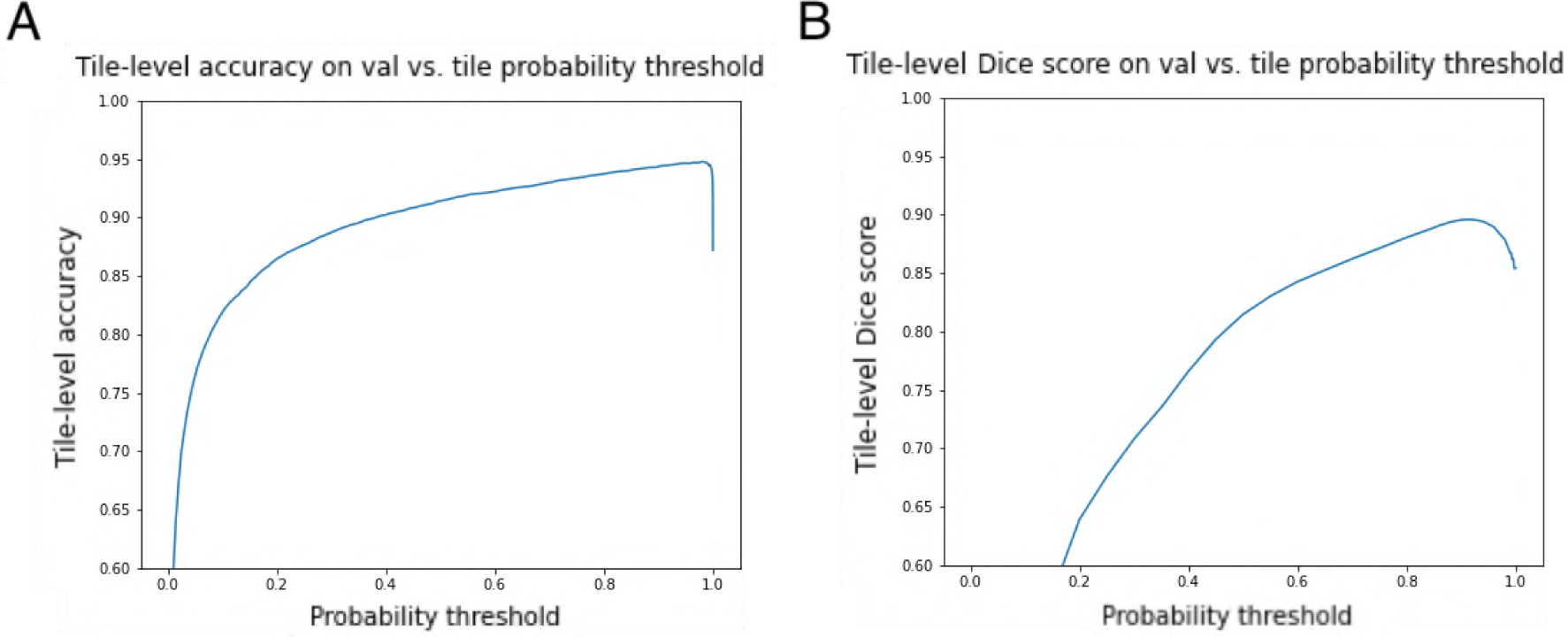
Selecting the probability threshold with the highest validation set performance. **(A)** Average tile-level accuracy of trained classification model on validation set across many inference probability thresholds to binarise tile-level predictions. **(B)** Average tile-level Dice coefficient of trained classification model on validation set across many inference probability thresholds to binarise pixel-level predictions.

### 5.6 Assessing performance on the test set

Once a best probability threshold has been determined, that best threshold can be applied to the test set to determine the model’s overall performance, as see in **Procedure steps 21–23** (classification) and **Procedure step 35** (segmentation). In classification tasks, there aren’t always tile-level labels in the test set; sometimes only slide-level labels are available. In these situations, an AUC can be computed comparing the number of positive tiles according to the best threshold in each test set slide (computable with `Slide.numTilesAboveClassPredictionThreshold()` as seen in **Procedure step 22**) against the slide-level ground truth labels. For classification tasks with tile-level ground truth labels on the test set, `Slide.classifierMetricAtThreshold()` can be applied to give an average tile-level accuracy of the test set (**Procedure step 21**). **Table 4** shows the average accuracy of the best-thresholded model on the six slides in the CAMELYON16 test set used in the **Procedure** along with the metastasis-positive tile count of each. The average accuracy across the test set is 0.97, which is very good, and the AUC comparing the positive tile counts to the slide level ground truths is 0.89 (see **fig. 10** for the AUC-ROC curve).

**Figure 10:**
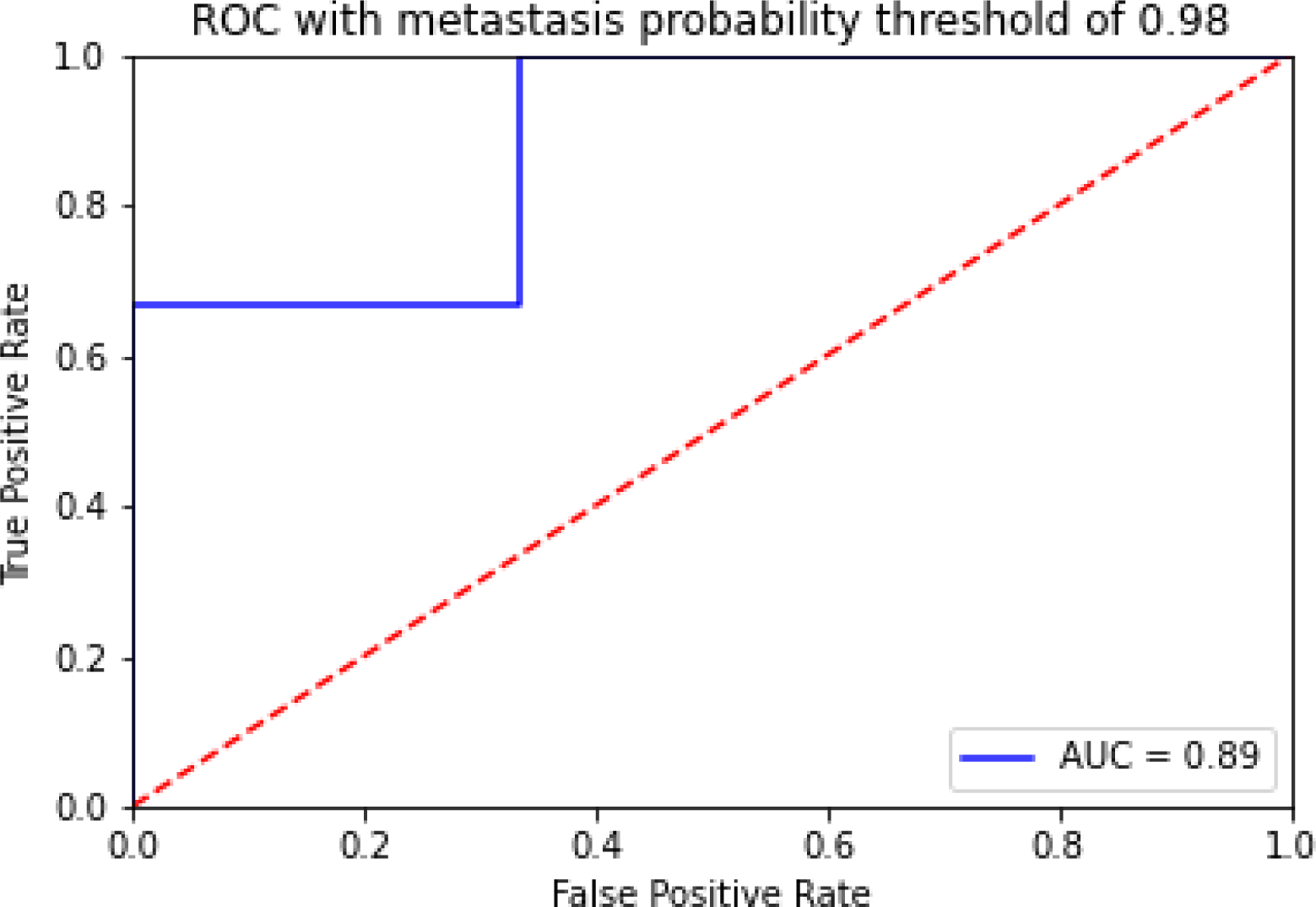
AUC-ROC curve comparing the number of metastasis-positive tiles in each slide with slide-level ground truth labels. The curve is very simplistic because for the sake of the Procedure tutorial, only 6 test set slides were used, and the metastasis tile count of the 3 metastasis-positive slides all exceed the metastasis tile counts of the 3 metastasis negative slides. In practice, test sets used for AUC analyses should be larger to give a more robust result.

**Table 4:** Test set classification accuracy on the metastasis class. Tiles with metastasis probability greater than 0.98 are considered positive. The number of tiles that cross that threshold are also shown. Results generated with Slide.classifierMetricatThreshold().

| Test set slide | Avg. metastasis accuracy | Num. metastasis tiles |
| --- | --- | --- |
| normal_028 | 0.998 | 3 |
| normal_074 | 0.983 | 97 |
| normal_141 | 0.999 | 3 |
| tumor_036 | 0.977 | 51 |
| tumor_058 | 0.940 | 589 |
| tumor_076 | 0.951 | 8863 |

Note that for the purposes of the procedure being computationally inexpensive, only six slides were used for the test set, which makes this AUC have little statistical power. Also, keep in mind that the number of tiles that cross the metastasis threshold depends heavily on the amount of tissue present in the slide; for example, `normal_074`, a metastasis-negative slide. still has 52 tiles that crossed that are considered metastatic at our operating threshold. This is because it is a massive slide with a lot of tissue in it, so the number of false positive tiles will be expected to be greater. Conversely, `tumor_036` only shows 51 metastasis tiles due to the dearth of tissue in this WSI, making a smaller number of metastasis tiles more significant. Users may want to normalise positive tile counts by the total number of suitable tissue tiles (the length of the list `Slide.suitableTileAddresses()` can be used to find this number) in a slide for this reason.

**Table 5** shows the Dice coefficient results of applying the best segmentation probability found from the validation set to the test set using `Slide.segmenterMetricAtThreshold()`. None of the test set slides have an average Dice coefficient below 0.82, and the mean Dice coefficient across all test set slides is 0.89.

**Table 5:** Test set segmentation Dice coefficients on the metastasis class. Pixels with metastasis probability greater than 0.91 are considered positive. Results generated with Slide.segmenterMetricatThreshold().

| Test set slide | Avg. metastasis Dice coefficient |
| --- | --- |
| normal_028 | 0.986 |
| normal_074 | 0.922 |
| normal_141 | 0.823 |
| tumor_036 | 0.902 |
| tumor_058 | 0.866 |
| tumor_076 | 0.859 |

Assessing what constitutes adequate test set performance depends heavily on the context of the dataset and the metric used to measure performance. For example, for a very difficult training task, a test set accuracy of 0.68 might be considered acceptable. On a more straightforward task, that performance might be woefully inadequate. In general, though, Dice coefficient is a more stringent metric than accuracy, so lower values are acceptable.

## 6 Code availability

The source code of PathML is freely available at a public repository: https://github.c `om/markowetzlab/pathml`. The source code of the tutorial presented in the **Procedure** section is also freely available at the following public repository: https://github.com/m `arkowetzlab/pathml-tutorial`. That repository also contains the code used to train PathML’s deep tissue detector in its `deep_tissue_detector` subdirectory.

## 7 Data availability

The WSIs used in the tutorial presented in the **Procedure** section, along with the annotation files demarcating regions of metastasis in each WSI, are publicly available at the following link: https://drive.google.com/drive/folders/0BzsdkU4jWx9Ba2x1NTZhdz `Q5Zjg?resourcekey=0-g2TRih6YKi5P2O1SiBB1LA`. Note that the `annotations` directory in the `pathml-tutorial` repository contains corrected versions of some of the annotation files present in the CAMELYON16 Google Drive link above (some of the original annotation files contained small self-overlapping regions), so the .xml files in `annotations` are recommended in lieu of the originals. A copy of the results from a full **Procedure**run can be found at the `pathml-tutorial` repository.

The WSIs used to train PathML’s deep tissue detector are governed by data usage policies specified by the data controller (University of Cambridge, Cancer Research UK). We are committed to complying with Cancer Research UK’s Data Sharing and Preservation Policy. Whole-slide images used in this study will be available for non-commercial research purposes upon approval by a Data Access Committee according to institutional requirements. Applications for data access should be directed to. The annotation .xml files corresponding to these WSIs are publicly available in the `deep_tissue_detector` subdirectory of `pathml-tutorial`.

## Data Availability

CODE AVAILABILITY:
The source code of PathML is freely available at the following public repository: https://github.com/markowetzlab/pathml. The source code of the tutorial presented in the Procedure section of the manuscript is also freely available at the following public repository: https://github.com/markowetzlab/pathml-tutorial. That repository also contains the code used to train PathML's deep tissue detector in its "deep_tissue_detector" subdirectory. DATA AVAILABILITY: The whole-slide images (WSIs) used in the tutorial presented in the Procedure section, along with the annotation files demarcating regions of metastasis in each WSI, are publicly available at the following link: https://drive.google.com/drive/folders/0BzsdkU4jWx9Ba2x1NTZhdzQ5Zjg?resourcekey=0-g2TRih6YKi5P2O1SiBB1LA. Note that the "annotations" directory in the pathml-tutorial repository (https://github.com/markowetzlab/pathml-tutorial) contains corrected versions of some of the annotation files present in the CAMELYON16 Google Drive link above (some of the original annotation files contained small self-overlapping regions), so the .xml files in "annotations" are recommended in lieu of the originals. A copy of the results from a full Procedure run can be found at the pathml-tutorial repository. A complete run of the tutorial presented in the Procedure section of the manuscript include all outputs can be found at the following link: https://doi.org/10.5281/zenodo.5006409. The WSIs used to train PathML's deep tissue detector are governed by data usage policies specified by the data controller (University of Cambridge, Cancer Research UK). We are committed to complying with Cancer Research UK's Data Sharing and Preservation Policy. Whole-slide images used in this study will be available for non-commercial research purposes upon approval by a Data Access Committee according to institutional requirements. Applications for data access should be directed to. The annotation .xml files corresponding to these WSIs are publicly available in the "deep_tissue_detector" subdirectory of the pathml-tutorial repository.

https://github.com/markowetzlab/pathml-tutorial

https://drive.google.com/drive/folders/0BzsdkU4jWx9Ba2x1NTZhdzQ5Zjg?resourcekey=0-g2TRih6YKi5P2O1SiBB1LA

https://doi.org/10.5281/zenodo.5006409

## Acknowledgments

We would like to thank Tristan Whitmarsh for his assistance with choosing suitable U-Net hyperparameters for the **Procedure**. We would also like to thank Sarah Killcoyne for her early testing of PathML, and Paula Martin-Gonzalez for validating the **Procedure**.

## 8 Author contributions

A.B. implemented the current version of PathML and wrote the **Procedure** section of the manuscript as well as an early draft of the **Introduction**. M.G. developed the original version of PathML. W.O. wrote the manuscript, created figures, and contributed ideas to the features and code of PathML. All work was performed under the supervision of F.M. The final manuscript was read and approved by all authors.

